# Epidemiological trends of osteoarthritis at the global, regional, and national levels from 1990 to 2021 and projections to 2050

**DOI:** 10.1101/2024.06.30.24309697

**Authors:** Lichun Qiao, Miaoqian Li, Feidan Deng, Xinyue Wen, Huan Deng, Zhaowei Xue, Ping Wan, Rongqi Xiang, Yanjun Xie, Huifang He, Xiangyu Fan, Yufei Song, Jun Wang, Jing Han

## Abstract

**Background:** Osteoarthritis is a major cause of disability worldwide, with its prevalence expected to increase due to ageing populations and rising obesity. Understanding the epidemiological trends in osteoarthritis is critical for public health planning and intervention strategies.

**Methods:** This study analyzed global, regional, and national data on osteoarthritis incidence, prevalence, and Disability-adjusted life years (DALYs) utilizing the Global Burden of Disease Study 2021. Statistical modelling was used to assess trends over the past 32 years and projections were made for 2050 based on demographic changes and historical data.

**Results:** In 2021, 607 (95% UI: 538-671) million people worldwide had osteoarthritis, with 46.6 (95% UI: 41.1-51.6) million new cases and 21.3 (95% UI: 10.2-42.9) million DALYs. Age-standardized incidence, prevalence and DALYs rates increased to 535.00 (95% UI: 472.38-591.97), 6967.29 (95% UI: 6180.70-7686.06), and 244.50 (95% UI: 117.06-493.11) per 100,000 population, with knee osteoarthritis accounting for more than 56%. Age-standardized rates of osteoarthritis were higher in females than in males. East Asia, South Asia, and Western Europe were the regions and China, India, and the United States were the countries with the highest burdens. In addition, high body-mass index (BMI) led to 4.43 (95% UI: -0.42-12.34) million DALYs, with an increase of 205.10%. Bayesian age-period cohort projections showed that the burden of osteoarthritis would continue to rise from 2021 to 2050.

**Conclusions:** The findings indicated that the burden of osteoarthritis is on a rising trend with an ageing population and increasing global obesity rates, with females and middle-aged and older age groups being the current populations of concern. Comprehensive public health policies and strategies are urgently needed to address its impact. Increased awareness, early detection and effective management are essential to reduce the burden of osteoarthritis in the coming decades, especially among vulnerable groups.

## 1 Introduction

Osteoarthritis is the most common musculoskeletal disorder worldwide, characterized by cartilage degeneration, changes in subchondral bone, synovial inflammation, and joint inflammation, resulting in symptoms such as pain, stiffness, disability, and function impairment [1-4]. The knees, hips, hands, feet, and spine are usually the typical sites of osteoarthritis, with a high prevalence of polyarticular involvement [4, 5]. Osteoarthritis is a significant contributor to disability and impaired quality of life in the elderly population, sometimes necessitating joint replacement surgery in advanced stages. One study reported a prevalence of 595 million individuals affected by osteoarthritis globally in 2020, accounting for 7.6% of the total population, with osteoarthritis being identified as the seventh leading cause of years lived with disability for those aged 70 years and older [6]. The epidemiological patterns of osteoarthritis have been the subject of extensive research in recent decades. In the wake of an ageing population and rising global obesity rates, there is growing attention directed towards comprehending its worldwide burden and implications for public health.

Risk factors for osteoarthritis primarily encompass human factors such as age, gender, obesity, genetics, and diet, as well as joint-level factors such as injury, misalignment, and abnormal loading, which interact with each other in a complex manner to influence the development of osteoarthritis [7]. Among these factors, age, gender, and obesity are recognized as independent risk factors for osteoarthritis. The prevalence of overweight obesity has emerged as a significant public health concern on a global scale. According to data from the World Health Organization, the incidence of obesity has more than doubled in adults and tripled in adolescents worldwide since 1990, and by 2022, approximately 2.5 billion adults will be classified as overweight. The Global Burden of Disease (GBD) Study indicated that high body-mass index (BMI) contributed to 20.4% (95% UI -1.7 to 36.6) of osteoarthritis cases in 2020 [6]. In light of risk factors such as the ageing population, sedentary lifestyles, and escalating rates of obesity, the incidence of osteoarthritis is rapidly increasing [7, 8], not only causing severe disease and economic burden but also putting pressure on healthcare systems [9, 10].

This study analyzed the epidemiological trends of osteoarthritis from 1990 to 2021 at global, regional, and national levels and the impact of factors such as sex, age, site of disease onset and high body mass index (BMI). At the same time, changes in the burden of osteoarthritis were projected for the next 29 years. The timely and comprehensive analyses of long-term epidemiological trends in osteoarthritis and projections of future burden trends in this study would help to inform future prevention and control measures and action strategies. It also highlights the urgent need for the prevention and management of osteoarthritis on a current and future global scale.

## 2 Methods

### 2.1. Study data source

The GBD 2021, led by the Institute for Health Metrics and Evaluation (IHME), provides the most comprehensive and up-to-date data assessment of the descriptive epidemiology of diseases in 21 regions and 204 countries and territories from 1990 to 2021. Data for this study were sourced from the Global Health Data Exchange Tool (http://ghdx.healthdata.org/gbd-results-tool), including the number and rate of incidence, prevalence, and disability-adjusted life years (DALYs) as well as age-standardized incidence rate (ASIR), age-standardized prevalence rate (ASPR), and age-standardized DALYs rate (ASDR) of osteoarthritis at the global, socio-demographic index (SDI) quintile, regional, and national levels. The burden data attributable to high BMI were also included. Moreover, countries were divided by SDI into five parts according to total fertility rate, per capita income, and average years of education, including low, low-middle, middle, high-middle, and high) to assess the relationship between osteoarthritis and social development status.

### 2.2. Definition

In the GBD 2021 study, osteoarthritis of the hip, knee, hand, and other joints was included, and osteoarthritis affecting the cervical spine, lumbar spine, or both sites was not included in the other osteoarthritis categories. The GBD defines the reference case for hip and knee osteoarthritis as symptomatic osteoarthritis, confirmed radiographically as the Kellgren-Lawrence grade 2-4 [11, 12]. Grade 2 indicates the presence of a definite osteophyte, grade 3 indicates multiple osteophytes and joint space narrowing, and grade 4 includes bone deformity along with the criteria of grade 3. Symptomatic osteoarthritis requires reporting pain for at least one month within the past 12 months [6]. The reference case for hand osteoarthritis is any single hand joint or the presence of several joints presenting with symptoms and radiologically confirmed as osteoarthritis [6]. In addition, given the lack of survey data, U.S. insurance claims data from 2000 to 2016 constitute the only source of other osteoarthritis data [6]. High BMI is a GBD risk factor for knee and hip osteoarthritis, which is defined as BMI greater than or equal to25 kg/m^2^ among people aged over 20 years, and a BMI of 20-25 kg/m^2^ is considered to be the theoretical minimum level of risk exposure [6, 13]. Previous studies have described detailed methods for comparative risk assessment for BMI estimation and high BMI [14].

### 2.3 Statistical Analysis

To compare the osteoarthritis burden in 2021 with that in 1990, changes in incident cases, prevalent cases, and DALYs were calculated. Estimated annual percentage changes (EAPCs) were calculated to reflect the temporal trends in age-standardized rates (ASRs) of osteoarthritis over the past 32 years. The ASRs are regarded as increasing if both the EAPC and the lower limit of the 95% uncertain interval (UI) are positive. In contrast, if the EAPC and the upper limit of the 95% UI are negative, the ASRs are considered to be decreasing. Also, the Pearson correlation coefficient was used to evaluate the relationship between SDI and ASRs of osteoarthritis at the global, regional, and national levels. In addition, the Bayesian age-period cohort (BAPC) analysis was used to predict the disease burden of osteoarthritis from 2021 to 2050. *P-*value < 0.05 was considered statistically significant.

## 3 Results

### 3.1 Burden at the global level

In 2021, there were approximately 46.6 (95% UI: 41.1-51.6) million incident cases of osteoarthritis, which represented an increase of 123.11% (95% UI: 120.62%-125.21%) compared to 1990 **(Table S1)**. The ASIR of osteoarthritis increased from 489.78/100,000 (95% UI: 433.10/100,000-541.51/100,000) in 1990 and 535.00/100,000 (95% UI: 472.38/100,000-591.97/100,000) in 2021, with an EAPC of 0.33 (95% CI: 0.31-0.35), which indicated that ASIR experienced a slight increase from 1990 to 2021 **(Table S2)**. Specifically by sex, the incidence cases for females were 27.70 (95% UI: 24.48-30.57) million in 2021, about 1.46 times higher than for males **(Table S1)**. In addition, the ASIR for females in 2021 was 621.32/100,000 (95% UI: 548.49/100,000-686.94/100,000) with an EAPC of 0.36 (95% CI: 0.32-0.39), and for males was 445.74/100,000 (95% UI: 393.68/100,000-493.98/100,000) with an EAPC of 0.28 (95% CI: 0.25-0.30) **(Table S2)**.

The prevalent cases of osteoarthritis increased from 256.08 (95% UI: 227.12-283.442) million in 1990 to 607 (95% UI: 538-671) million in 2021, representing an estimated increase of 137.03% (95% UI: 135.49%-138.71%) **(Table S1)**. From 1990 to 2021, the ASPR increased relatively slightly, from 6393.12/100,000 (95% UI: 5683.20/100,000-7059.53/100,000) to 6967.29/100,000 (95% UI: 6180.70/100,000-7686.06/100,000), and the EAPC was 0.34 (95% CI: 0.31-0.37) **(Table S2)**. Although the prevalent cases for females were higher than for males, the increasing change in males was higher than in females [139.55% (95% UI: 137.55%-141.80%) vs. 135.43% (95% UI: 134.00%-136.97%)] in 2021 **(Table S1)**. The ASPR for females was also higher than males, with the former approximately 1.39 times higher than the latter both in 1990 and 2021 **(Table S2)**.

Over the past 32 years, the DALYs number has increased by 138.87% (95% UI: 136.83%-140.75%), reaching 21.30 (95% UI: 10.19-42.94) million in 2021 **(Table S1)**. As for ASDR, it increased from 222.80/100,000 (95% UI: 106.65/100,000-450.29/100,000) in 1990 to 244.50/100,000 (95% UI: 117.06/100,000-493.11/100,000) in 2021, with an EAPC of 0.37 (95% CI: 0.33-0.40) **(Table S2)**. Furthermore, the DALYs number and rate were higher for females than for males, with the former being 1.57 and 1.42 times higher than the latter in 2021, respectively **(Table S1-S2)**. Notably, from 1990 to 2021, the ASDR for females increased from 257.65/100,000 (95% UI: 123.39/100,000-520.40/100,000) to 284.14/100,000 (95% UI: 136.29/100,000-573.11/100,000) and the EAPC was 0.42 (95% CI: 0.37-0.47) **(Table S2)**.

For the different types of osteoarthritis, the incident cases and prevalent cases of osteoarthritis were ranked in descending order as knee, hand, other, and hip osteoarthritis, whereas the DALYs number was ranked in descending order as knee, hand, hip and other osteoarthritis **(Table S1)**. The incident case, prevalent case, and DALYs number for knee osteoarthritis were 30.856 (95% UI: 26.53-35.19) million, 374.74 (95% UI: 321.86-428.35) million, 12.02 (95% UI: 5.86-23.27) million in 2021. Globally, the ASRs were all ranked in descending order as knee, hand, other, and hip osteoarthritis **(Table S2)**. The ASIR, ASPR, and ASDR for knee osteoarthritis were 353.67/100,000 (95% UI: 304.56/100,000-402.50/100,000), 4294.27/100,000 (95% UI: 3695.04/100,000-4910.76/100,000), and 137.59/100,000 (95% UI: 67.08/100,000-266.87/100,000).

### 3.2 Burden at the regional level

At the regional level, there was a varying degree of increase in incident cases, prevalent cases, DALYs number, and ASRs of osteoarthritis across the 21 regions over the past 32 years **(Table S1-S2 and Figure S1)**. The EAPCs for ASRs were all greater than 0 **(Figure S2)**. In 2021, the top three regions with the incident cases, prevalent cases and DALYs numbers were East Asia, South Asia, and Western Europe, while the top three regions with the ASIR, ASPR and ASDR were high-income Asia Pacific, high-income North America, and Australasia **(Table S3)**. Among these regions, East Asia had the highest cases and number of 12.05 (95% UI: 10.56-13.56) million, 158.29 (95% UI: 139.47-176.82) million, and 5.52 (95% UI: 2.63-11.06) million, which increased by 149.17% (95% UI: 141.74%-155.84%), 185.17% (95% UI: 178.43%-191.41%), and 189.79% (95% UI: 182.36%-197.27%) in 2021, respectively. Furthermore, High-income Asia Pacific had the highest ASIR, ASPR, and ASDR of 682.07/100,000 (95% UI: 606.06/100,000-752.84/100,000), 8608.63/100,000 (95% UI: 7674.07/100,000-9485.19/100,000), and 314.98/100,000 (95% UI: 150.55/100,000-636.77/100,000).

Across sexes, females had higher incident cases, prevalent cases, DALYs number, and ASRs than males in 1990 and 2019 **(Table S1-S2 and Figure 1)**. The top three regions with the incident cases, prevalent cases and DALYs numbers for males and females were also East Asia, South Asia, and Western Europe, among which East Asia has the highest cases and numbers **(Table S3)**. The top three regions with the ASRs for females were also the high-income Asia Pacific, high-income North America, and Australasia. Although the top three regions with the ASRs for males varied, high-income North America had the highest ASRs **(Table S3)**. Knee and hand osteoarthritis dominated the ASRs in most regions, followed by other types of hip osteoarthritis in 1990 and 2021 **(Figure 2)**. In most regions, knee osteoarthritis accounted for over 50%, with East Asia having the highest proportion, where the ASIR, ASPR, and ASDR of knee osteoarthritis reached 73.3%, 66.0%, and 66.3% in 2021.

**Figure 1.**
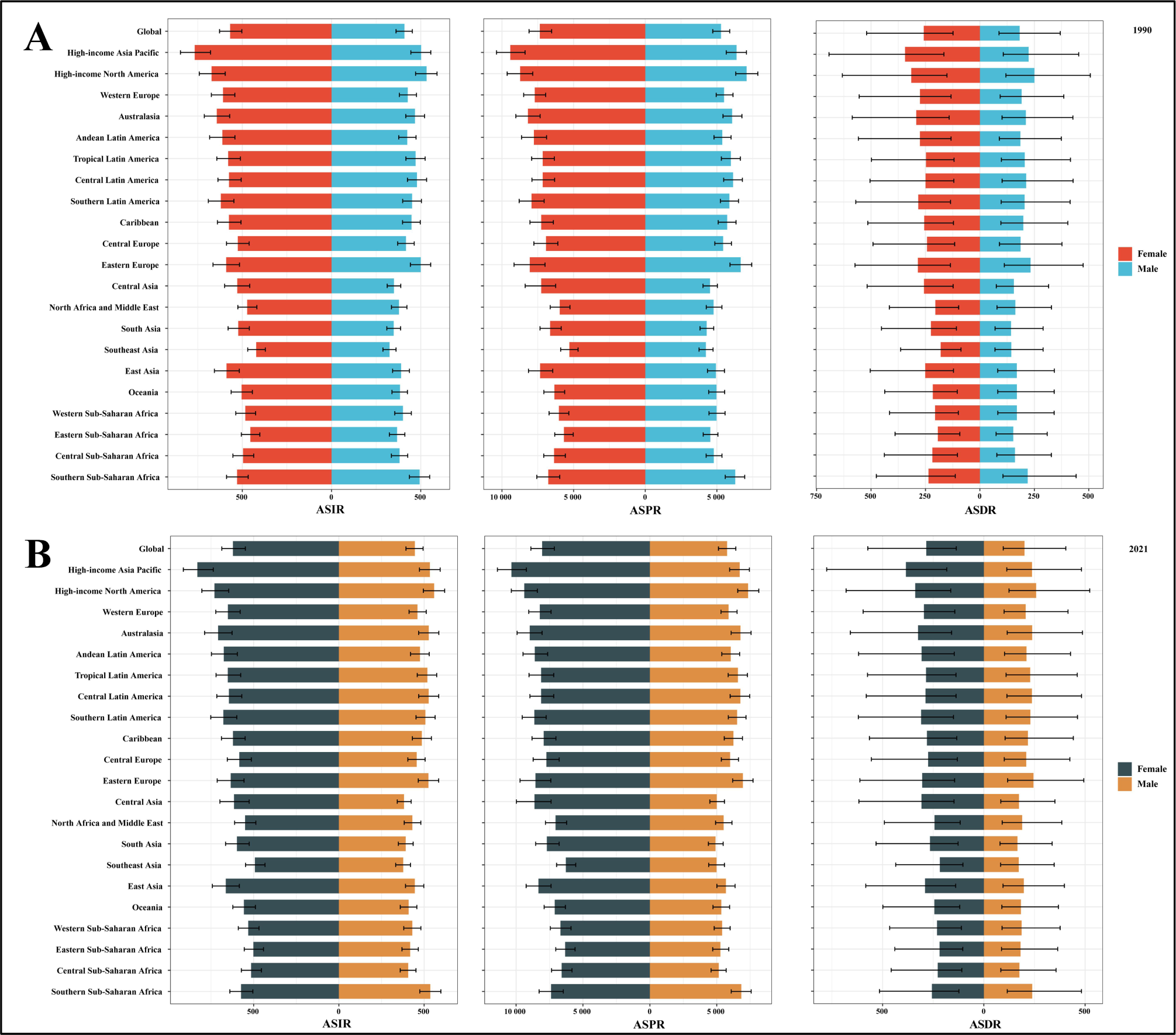
The ASRs (per 100,000 population) of osteoarthritis by sex at the global and regional levels in 1990 and 2021. **(A)** ASIR, ASPR, and ASDR in 1990. **(B)** ASIR, ASPR, and ASDR in 2021. ASRs: age-standardized rates; ASIR: age-standardized incidence rate; ASPR: age-standardized prevalence rate; ASDR: age-standardized disability-adjusted life years rate.

**Figure 2.**
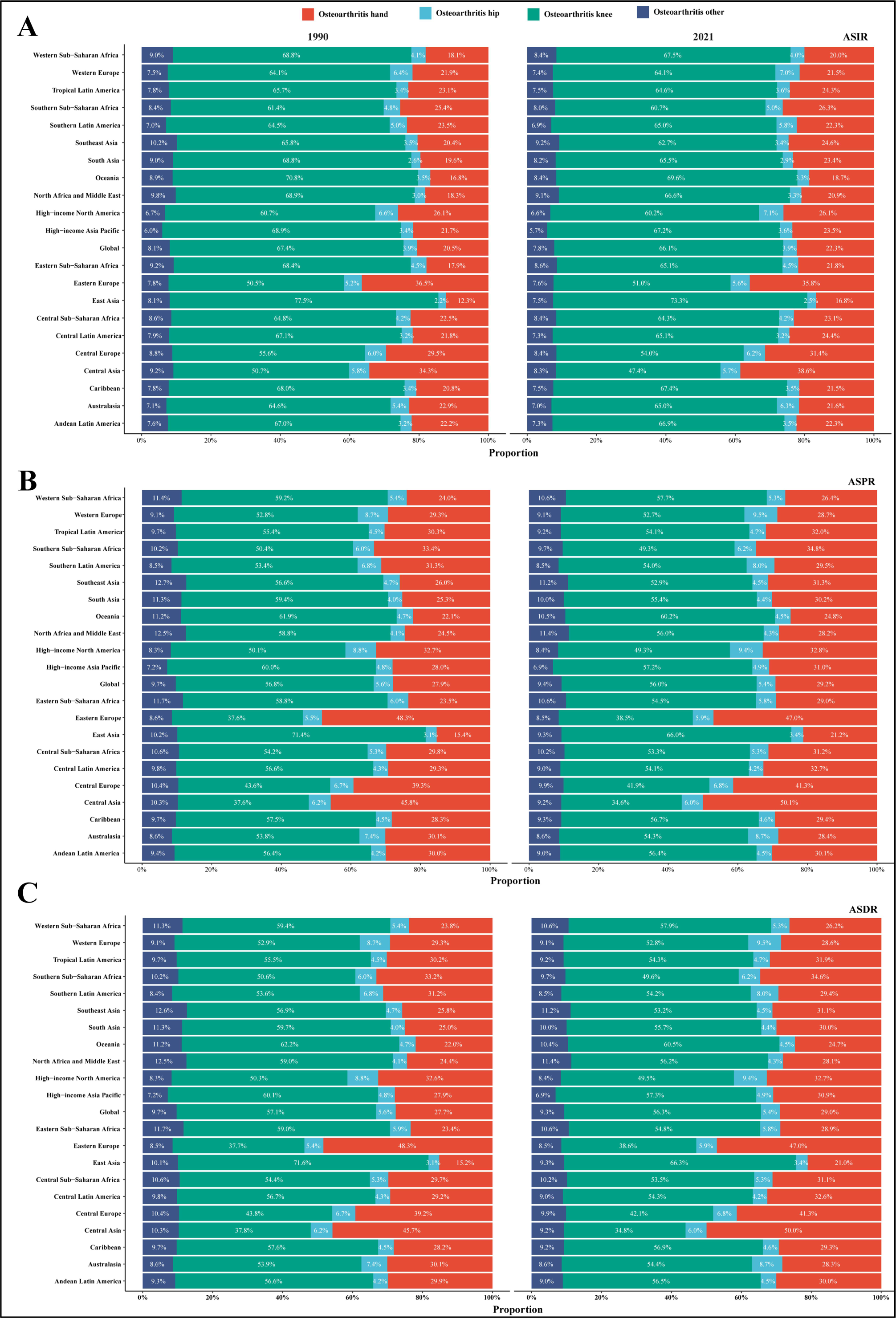
The proportion of ASRs (per 100,000 population) of knee, hand, hip, and other osteoarthritis at the global and regional levels in 1990 and 2021. ASRs: age-standardized rates; ASIR: age-standardized incidence rate; ASPR: age-standardized prevalence rate; ASDR: age-standardized disability-adjusted life years rate.

### 3.3 Burden at the national level

The incident cases, prevalent cases, DALYs number, and ASRs of osteoarthritis increased in most 204 countries and territories from 1990 to 2021 **(Table S4-S5)**. The top three nations with the incident cases, prevalent cases, and DALYs number of osteoarthritis for both sexes, males, and females were China, India, and the United States in 1990 and 2019 **(Table S6, Figure 3 and Figure S3)**. Among these countries and territories, China had the highest cases and number of 11.65 (95% UI: 10.21-13.11) million, 158.85 (95% UI: 134.66-170.84) million, and 5.33 (95% UI: 2.54-10.68) million, which increased by 150.37% (95% UI: 142.75%-155.84%), 186.49% (95% UI: 179.62%-192.93%), and 191.21% (95% UI: 183.62%-198.69%), respectively. Moreover, the increasing changes among 204 countries and territories are presented in **Figure S4**.

**Figure 3.**
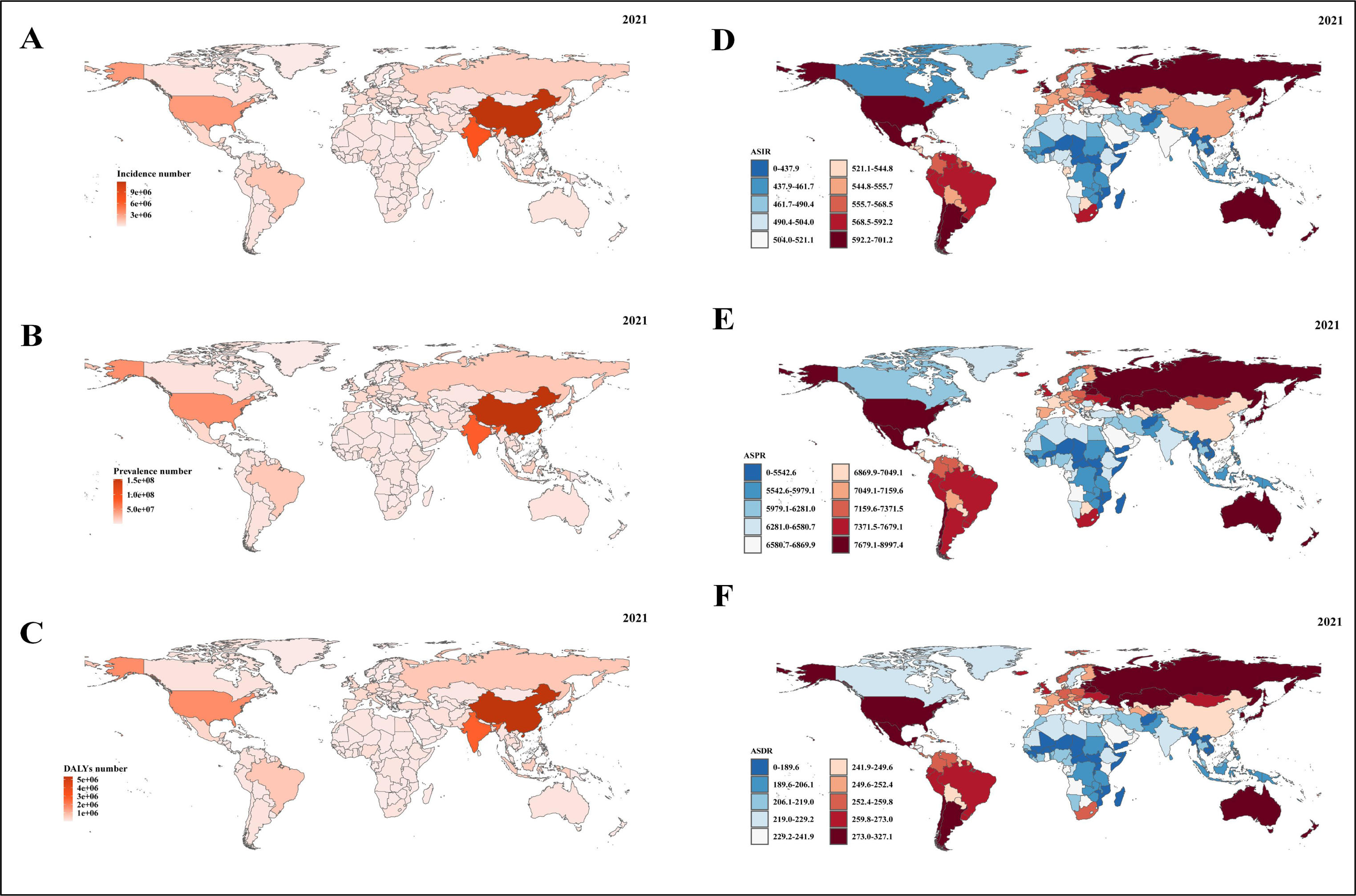
The incident cases, prevalent cases, DALYs, and ASRs (per 100,000 population) of osteoarthritis at the national level in 2021. **(A)** Incident case. **(B)** Prevalent case. **(C)** DALYs number. **(D)** ASIR. **(E)** ASPR. **(F)** ASDR. DALYs: disability-adjusted life years rate; ASRs: age-standardized rates; ASIR: age-standardized incidence rate; ASPR: age-standardized prevalence rate; ASDR: age-standardized disability-adjusted life years rate.

In 2021, the Republic of Korea, Brunei Darussalam, and Singapore exhibited the highest ASRs estimates **(Table S6 and Figure 3)**. Among these countries and territories, the Republic of Korea had the highest ASRs of 701.23/100,000 (95% UI: 625.36/100,000-776.78/100,000), 8997.39/100,000 (95% UI: 8082.99/100,000-9897.80/100,000), and 327.14/100,000 (95% UI: 157.44/100,000-662.81/100,000). The top three countries and territories with the ASRs for females were the same as the ASRs for both sexes. For males, the United States of America had the highest ASRs. The EAPCs for ASRs among 204 countries and territories are illustrated in **Figure S5**.

### 3.4 Burden at the SDI level

There was an increase in incident cases, prevalent cases, and DALYs number of osteoarthritis across SDI quintiles in 2021, with the most significant rise in the middle SDI quintile [168.44% (95% UI: 162.44%-173.96%), 195.94% (95% UI: 190.22%-201.67%), and 200.09% (95% UI: 193.73%-206.34%)] **(Table S1)**. The middle SDI quintile exhibited the highest incident cases, prevalent cases, and DALYs number in 2021, while the high SDI quintile had the highest ASRs, followed by the high-middle, middle, low-middle, and low SDI quintiles **(Table S1-S2)**. Moreover, the ASRs for both sexes, males, and females in the high and high-middle SDI quintiles exceeded the global level in 2021 **(Figure S6)**. The rankings of ASRs for the four types of osteoarthritis were consistent with global trends **(Figure S7)**.

In addition, positive associations were observed at the regional level, indicating that osteoarthritis burden for both sexes, males and females increases with higher SDI **(Figure S8-S10)**. The burden estimates in Western Sub-Saharan Africa, Southern Sub-Saharan Africa, Central Latin America, Tropical Latin America, the Caribbean, Southern Latin America, Eastern Europe, and high-income Asia Pacific exceeded expectations based on SDI from 1990 to 2021. At the national level, a positive association was identified between ASRs and SDI for 204 countries and territories in 2021, indicating an increase in the burden of osteoarthritis with higher SDI levels **(Figure S11)**. Similar positive correlations were also observed across different sexes **(Figure S12-S13)**. Notably, the United States of America, the Republic of Korea, Brunei Darussalam, and Singapore exhibited a burden that exceeded expectations, while Viet Nam, Afghanistan, Canada, and Cambodia displayed a burden lower than anticipated.

### 3.5 Burden based on age and sex

In 1990 and 2021, the incident cases and incidence rate increased and then decreased with age, and were consistently higher in females than in males. In addition, the incident cases and incidence rates were higher mainly in the age subgroups 45-49, 50-54 and 55-59 years **(Figure S14A-S14B)**. Prevalent cases and DALYs number increased and then decreased with age, while prevalence and DALYs rates continued to increase with age and were consistently higher among females than males **(Figure S14C-S14F)**. In addition, prevalent cases and DALYs number were higher mainly in the age subgroups 55-59, 60-64 and 65-69 years.

### 3.6 Burden attributable to high BMI at the global and SDI levels

**Table S7** presents the distribution and variations of DALYs number and ASDR of osteoarthritis attributable to high BMI. Globally, the DALYs number was 4.43 (95% UI: -0.42-12.34) million in 2021, representing a 205.10% (95% UI: 191.71%-227.09%) increase compared to 1990. Over the past 32 years, the ASDR has demonstrated a consistent rise from 35.97/100,000 (95% UI: -3.14/100,000-102.31/100,000) in 1990 to 50.59/100,000 (95% UI: -4.81/100,000-141.35/100,000), with an EAPC of 1.17 (95% CI: 1.15-1.20) **(Figure 4A and Table S7)**. In the GBD 2021 study, the ASDR for osteoarthritis due to high BMI rose from 8th in 1990 to 7th among all 28 diseases **(Table S15)**. The ASDR for females in 2021 was approximately 1.54 times higher than that of males. Specifically, the DALYs number attributable to knee and hip osteoarthritis due to high BMI increased globally by 207.64% (95% UI: 194.14%-229.71%) and 181.92% (95% UI: 169.20%-204.89%). The DALYs number and ASDR for knee osteoarthritis were approximately 10 and 9 times higher than those for hip osteoarthritis in 2021 **(Table S7)**. The DALYs number tended to peak in the 60-64 and 55-59 age groups in 1990 and 2021, with higher numbers in females than males **(Figure 4B-4C)**. In addition, the proportion of DALYs number attributable to total osteoarthritis, knee osteoarthritis, and hip osteoarthritis due to high BMI increased annually **(Figure 4D and Figure S16-S17)**.

**Figure 4.**
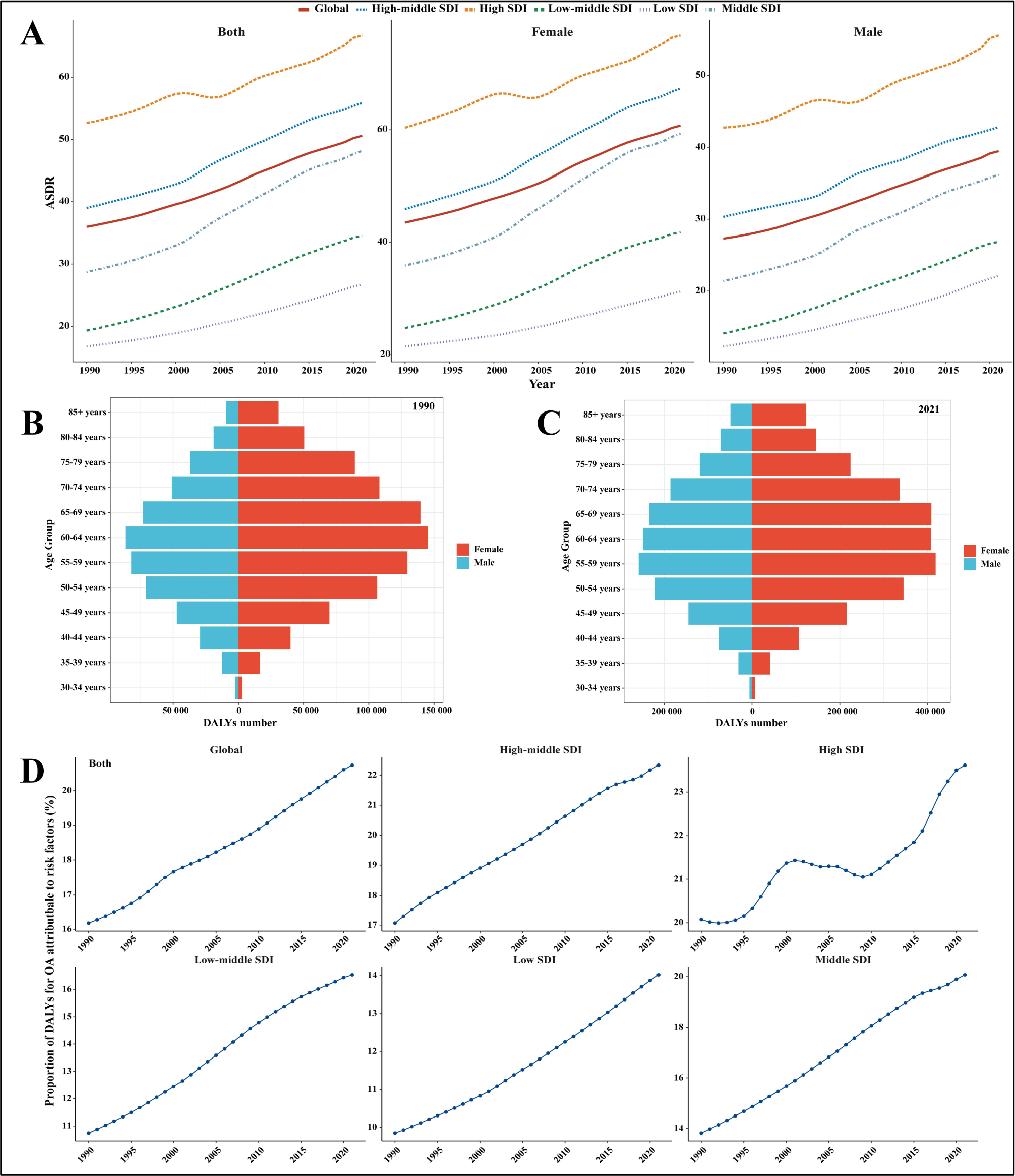
The disease burden of osteoarthritis attributable to high BMI at the global and SDI quintile levels. **(A)** ASDR (per 100,000 population) by sex from 1990 to 2021. **(B-C)** DALYs by age and sex in 1990 and 2021. **(D)** Proportion of DALYs attributable to high BMI among both sexes from 1990 to 2021. BMI: body-mass index; SDI: socio-demographic index; DALYs: disability-adjusted life years; ASDR: age-standardized disability-adjusted life years rate.

The order of increase in DALYs number was observed as follows: middle, low-middle, low, high-middle, and high SDI quintiles. The highest increase amounted to 329.11% (95% UI: 302.47%-367.69%), while the lowest was 123.35% (95% UI: 112.98%-144.93%). Moreover, the ASDR exhibited a steady upward trajectory over the past 32 years, with the highest ASDR being documented at 66.70/100,000 (95% UI: -6.82/100,000-179.94/100,000) in the high SDI quintile. The ASDRs in the high and high-middle quintiles exceeded the global level, whereas the remaining quintiles fell below it **(Figure 4A and Table S7**). The proportion of DALYs of osteoarthritis across the five SDI quintiles increased, with values of 22.623%, 22.33%, 20.07%, 16.53%, and 14.02% in 2021, descending from the high SDI quintile to low SDI quintile **(Figure 4D and Figure S16)**. The proportion of attributable DALYs of knee and hip osteoarthritis also increased **(Figure S17)**.

### 3.7 Burden attributable to high BMI at the regional and national levels

At the regional level, Southeast Asia, Southern Latin America, and East Asia were the three regions with the fastest growth in ASDR, with EAPCs of 2.65 (95% CI: 2.58-2.73), 2.65 (95% CI: 2.58-2.73), and 2.47 (95% CI: 2.33-2.60) **(Figures 5A and Table S8)**. The ASDR positively correlated with SDI across different regions. Latin America, Australasia, and Oceania were experiencing faster-than-expected growth, while East Asia, Southeast Asia, and Eastern Europe were growing at a slower-than-expected pace **(Figure 5B)**. At the national level, Bangladesh, Viet Nam, and India were the three regions with the fastest growth in ASDR, with EAPCs of 3.11 (95% CI: 2.99-3.23), 2.82 (95% CI: 2.66-2.98), and 2.70 (95% CI: 2.63-2.78) **(Figures 5C-5E and Table S8)**. There was a positive correlation between ASDR and SDI in various countries. Notably, the United States of America, Australia, and the United Kingdom were observing faster-than-expected growth, while China, India, and Japan were experiencing lower growth than expected **(Figure 5F)**.

**Figure 5.**
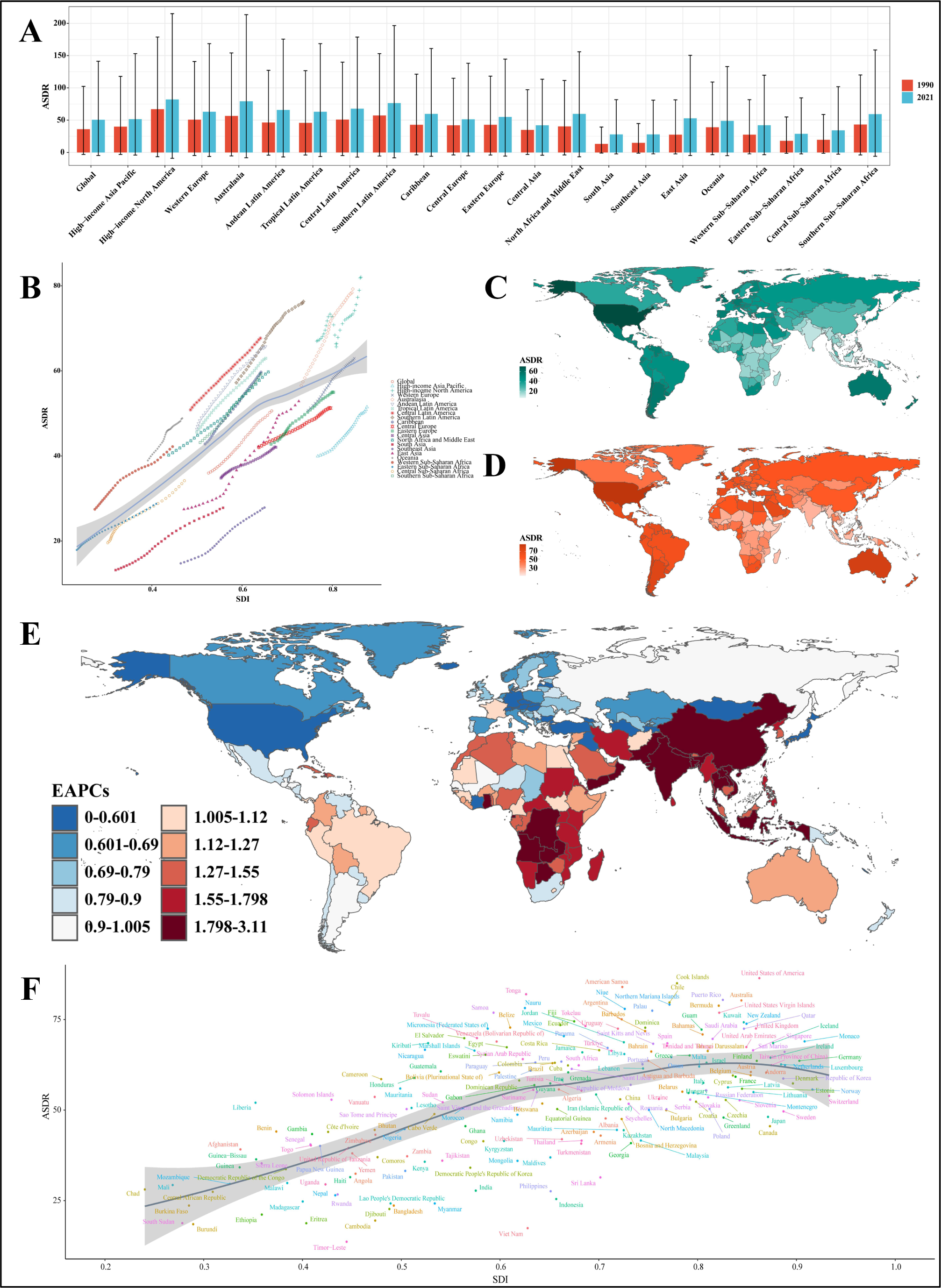
The burden of osteoarthritis attributable to high BMI at the national and national levels. **(A)** ASDR (per 100,000 population) at the national level in 1990 and 2021. **(B)** Correlation between SDI and ASDR at the global and regional levels (r=0.6815, *P*<0.001). **(C-D)** ASDR in 1990 and 2021. **(E)** EAPC. **(F)** Correlation between SDI and ASDR at the national level (r=0.5764, *P*<0.001). EAPC: estimated annual percentage change; BMI: body-mass index; SDI: socio-demographic index; DALYs: disability-adjusted life years; ASDR: age-standardized disability-adjusted life years rate.

### 3.8 Projections of burden until to 2050

To obtain an overview of the epidemiological trends of osteoarthritis over the next 29 years, we projected the ASRs of total osteoarthritis and rough rates across different age subgroups using the BAPC models. The ASRs of osteoarthritis among both sexes, males, and females would increase annually until 2050. ASIR, ASPR, and ASDR would increase from 519.49/100,000 (95% CI: 519.39/100,000-519.69/100,000), 6906.07/100,000 (95% CI: 6905.51/100,000-6906.62/100,000) and 242.52/100,000 (95% CI: 242.41/100,000-242.62/100,000) in 2021 to 589.54/100,000 (95% CI: 409.84/100,000-769.23/100,000), 7993.76/100,000 (95% CI: 6479.29/100,000-9508.24/100,000) and 281.18/100,000 (95% CI: 229.30/100,000-333.06/100,000) in 2050, respectively **(Figure S18)**. The predicted ASRs of osteoarthritis would continue to be higher for females than males **(Figure S19-S20)**. The predicted incidence, prevalence, and DALYs rates of osteoarthritis in different age subgroups would show varying degrees of trend, ultimately showing an increase in 2050 **(Figure S21-S23)**.

## 4 Discussion

At present, osteoarthritis continues to be a significant public health problem globally. As it is a highly prevalent and disabling disease, it imposes a tremendous disease burden. A comprehensive understanding of the epidemiological trends of osteoarthritis is essential for developing and implementing effective prevention and treatment strategies. This study presents the most recent findings on the disease burden of osteoarthritis from 1990 to 2021 at the global, SDI quintile, regional, and national levels based on the GBD Study 2021. The epidemiological trends indicated an increase in incident cases, prevalent cases, DALYs number, and ASRs of osteoarthritis, which increased globally and in all GBD regions and most countries and territories, consistent with previous GBD study reports [6, 15]. DALYs and ASDR of osteoarthritis related to high BMI showed similar trends. The projected trends from 2021 to 2050 suggest that the disease burden of osteoarthritis will continue to grow, posing a challenge for healthcare systems worldwide.

The diagnosis of osteoarthritis has improved significantly with the continuous improvement of medical diagnostic and treatment technology, enabling more accurate assessment of the location and type of lesion [15]. Until 2021, there were 46.6 million incident cases, 607.0 million prevalent cases, and 21.3 million DALYs globally, representing a 137.03% increase in osteoarthritis patients. In particular, the knee osteoarthritis represented the largest proportion globally. Therefore, it is imperative to prioritise preventive measures, as well as the management and treatment of osteoarthritis, especially knee osteoarthritis. Notably, in previous studies and the present study, the incidence, prevalence, and DALYs rates of osteoarthritis were found to increase with age, possibly due to degenerative changes in different aspects of the capabilities of the body with advancing age [16]. Many patients with advanced osteoarthritis even need to undergo joint replacement. In addition, the burden of osteoarthritis was significantly higher in females than in males. Studies have shown that oestrogen plays an important role in maintaining the homeostasis of joint tissues, as well as the joints themselves, and therefore, gender variability in osteoarthritis is thought to be related to oestrogen-related receptor effects and reduced oestrogen levels [17-19]. These suggested that in formulating future prevention and treatment strategies and implementing measures, emphasis should be placed on the female and middle-aged and elderly groups. The burdens of osteoarthritis were imbalanced across the five SDI quintiles, which might be explained by the inequalities in obtaining health care [20-23]. Studies have shown that socioeconomic status relates to access to health care, and people with higher socioeconomic status are more likely to have access to specialized health care [24, 25]. Although osteoarthritis in the high SDI quintile had the highest ASRs, its burden was lower than the middle SDI quintile, which might be attributed to advanced medical conditions [26, 27].

The differences in osteoarthritis burden across regions and countries may be due to genetic, metabolic, and behavioural factors [6]; in addition, they may also be related to various factors such as ageing populations, rising obesity rates and lack of access to healthcare services. In 2021, the burden of osteoarthritis was highest in East Asia, South Asia and Western Europe and lowest in the Caribbean and Oceania. The latest World Bank report states that the population of East Asia is already ageing faster than any other region in history. Furthermore, the ASRs were highest in high-income Asia Pacific, high-income North America and Australasia and lowest in Eastern Sub-Saharan Africa, Central Sub-Saharan Africa and Southeast Asia in 2021. The higher ASRs in high-income country regions may be related to better healthcare and higher income perception in these regions, allowing more osteoarthritis cases to be diagnosed and included in the statistics. In contrast, the lower ASRs may be related to factors such as a lack of local healthcare resources, inadequate medical care, and lack of awareness of osteoarthritis, which has led to under-diagnosis and under-recording of osteoarthritis cases in the region. At the national level, China, India and the United States have the highest osteoarthritis burdens. The three countries are the top three populous countries in the world, with the largest populations in both developing and developed countries. Their osteoarthritis burdens are the highest, firstly, because of the large population bases of these three countries, and secondly, ageing is a common trend in the demographic changes of the three countries. One of our key findings is that osteoarthritis is becoming more prevalent in developing countries, even though it has traditionally been more common in developed countries. This shift might be attributed to factors such as urbanization, sedentary lifestyles, and changes in dietary habits leading to obesity, all of which are known risk factors for osteoarthritis. In addition, there was a positive association between SDI and ASRs both at the regional and national levels, indicating that the higher the SDI, the higher the ASR is likely to be. Consistent with previous reports [15, 16].

This study also provides the first systematic elucidation of the global burden of disease associated with BMI in osteoarthritis from 1990 to 2021. The DALYs of osteoarthritis attributable to high BMI in 2021 was 4.43 million, accounting for 21% of the total DALYs globally, in line with a GBD 2019 study [1]. Moreover, the attributable DALYs to knee osteoarthritis increased globally by 207.64%. There is no cure for osteoarthritis despite its heavy burden, and prevention of osteoarthritis should focus on its modifiable risk factors, such as the BMI [16, 28-30]. Prospective studies have shown that being overweight or obese increases the risk of hand, hip, and knee osteoarthritis, with knee osteoarthritis at greatest risk, and that the risk increases in a dose-response gradient with increasing BMI [31, 32]. Weight loss in obese patients with knee osteoarthritis is clinically beneficial, as it can reduce pain and improve function [33]. In this study, osteoarthritis burden was more highly correlated with high BMI in countries with higher SDI than countries with lower SDI. Due to relatively poor economic development, access to adequate food remains a problem in many countries with lower SDI, and people are more likely to engage in physical activities that require high energy expenditure [1, 9]. In contrast, higher-income people consume more fat, salt, and processed foods and have higher rates of obesity [9].

In the next 30 years, the burden of disease in osteoarthritis will continue to rise; it is essential to prioritize preventive measures and early interventions for osteoarthritis. Public health initiatives promoting physical activity, healthy diet, and weight management can help reduce the risk of developing the disease. Additionally, healthcare systems should focus on improving access to affordable treatments and rehabilitation services for individuals with osteoarthritis. In conclusion, the epidemiological trends of osteoarthritis highlight the growing burden of this condition on a global scale. Addressing this challenge will require a multi-faceted approach involving public health efforts, healthcare system reforms, and strategies to reduce disparities in access to care. By taking proactive steps now, we can work towards mitigating the impact of osteoarthritis and improving the quality of life for individuals affected by this chronic condition.

## Data Availability

The data used for the analyses are publicly available from http://ghdx.healthdata.org/gbd-results-tool.

https://vizhub.healthdata.org/gbd-results/

## Abbreviations

GBD: Global Burden of Disease
BMI: High body-mass index
IHME: Health Metrics and Evaluation
DALYs: Disability-adjusted life years
ASIR: Age-standardized incidence rate
ASPR: Age-standardized prevalence rate
ASDR: age-standardized DALYs rate
SDI: Socio-demographic index
EAPCs: Estimated annual percentage changes
ASRs: Age-standardized rates
UI: Uncertain interval
BAPC: Bayesian age-period cohort
CI: Confidence interval

## Acknowledgments

We thank the staff of the Institute for Health Metrics and Evaluation and its collaborators, and Xiao Ming for his work in the GBD database. We also thank HOME for Researchers (https://www.home-for-researchers.com/) for their help with writing in English.

## Author contribution

LCQ, MQL, and JH designed the study. FDD, XYW, PW, YJX, HFH and XYF analysed the data and performed the statistical analyses. LCQ, MQL, ZWX, YFS, and RQX drafted the initial manuscript. JW, HD, and JH reviewed the drafted manuscript for critical content. All authors approved the final version of the manuscript.

## Declarations

### Ethics approval and consent to participate

Not applicable.

### Consent for publication

Not applicable.

