## Supplementary materials for "Epidemiological trends of osteoarthritis at the global, regional, and national levels from 1990 to 2021 and projections to 2050"

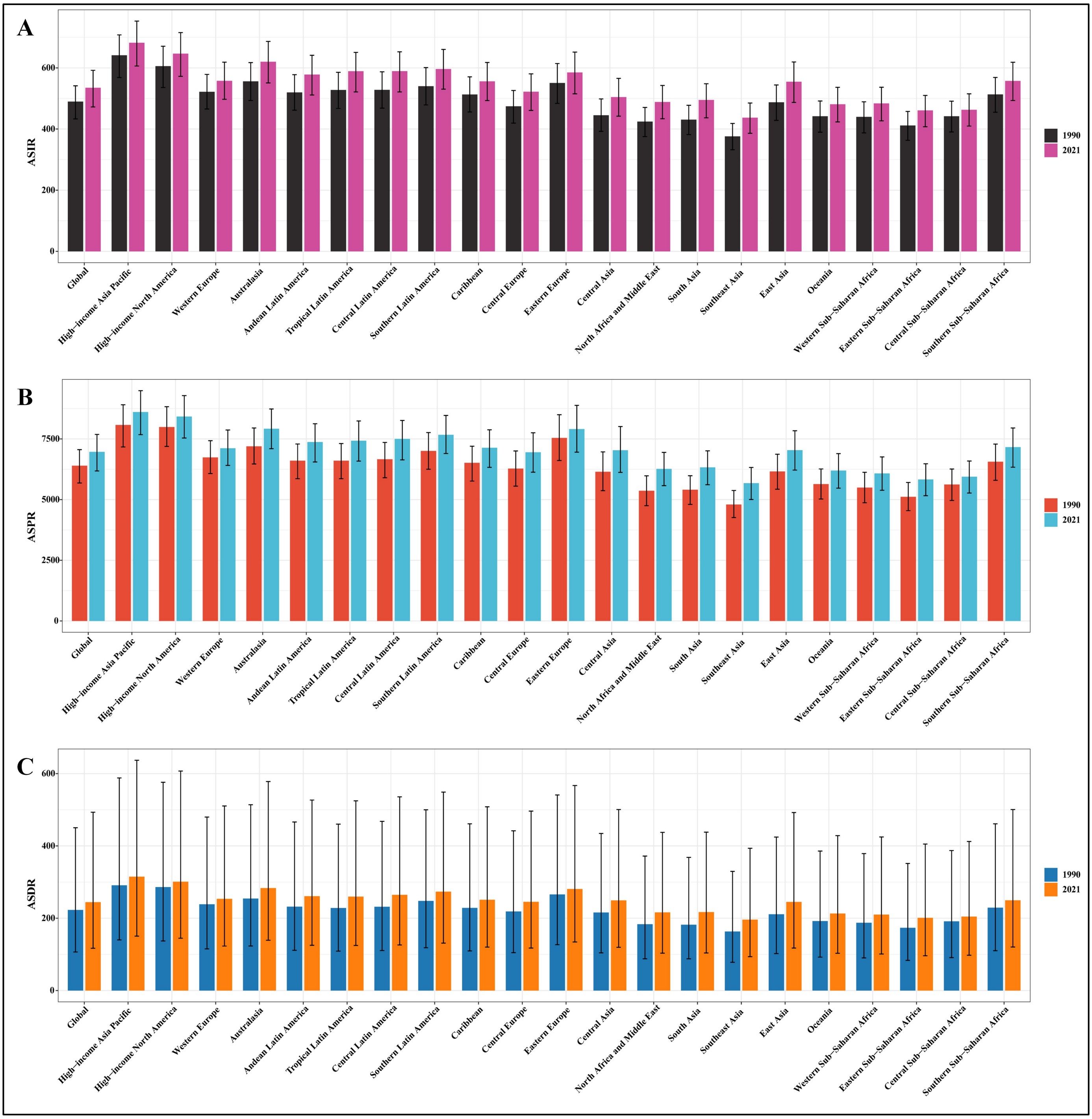


**Figure S1.** The ASRs (per 100,000 population) for osteoarthritis among both sexes at the global and regional levels in 1990 and 2021. **(A)** ASIR. **(B)** ASPR. **(C)** ASDR. ASR age-standardized rate; ASIR: age-standarzied incidence rate; ASPR: age-standarzied prevalence rate; ASDR: age-standardized disability-adjusted life years rate.

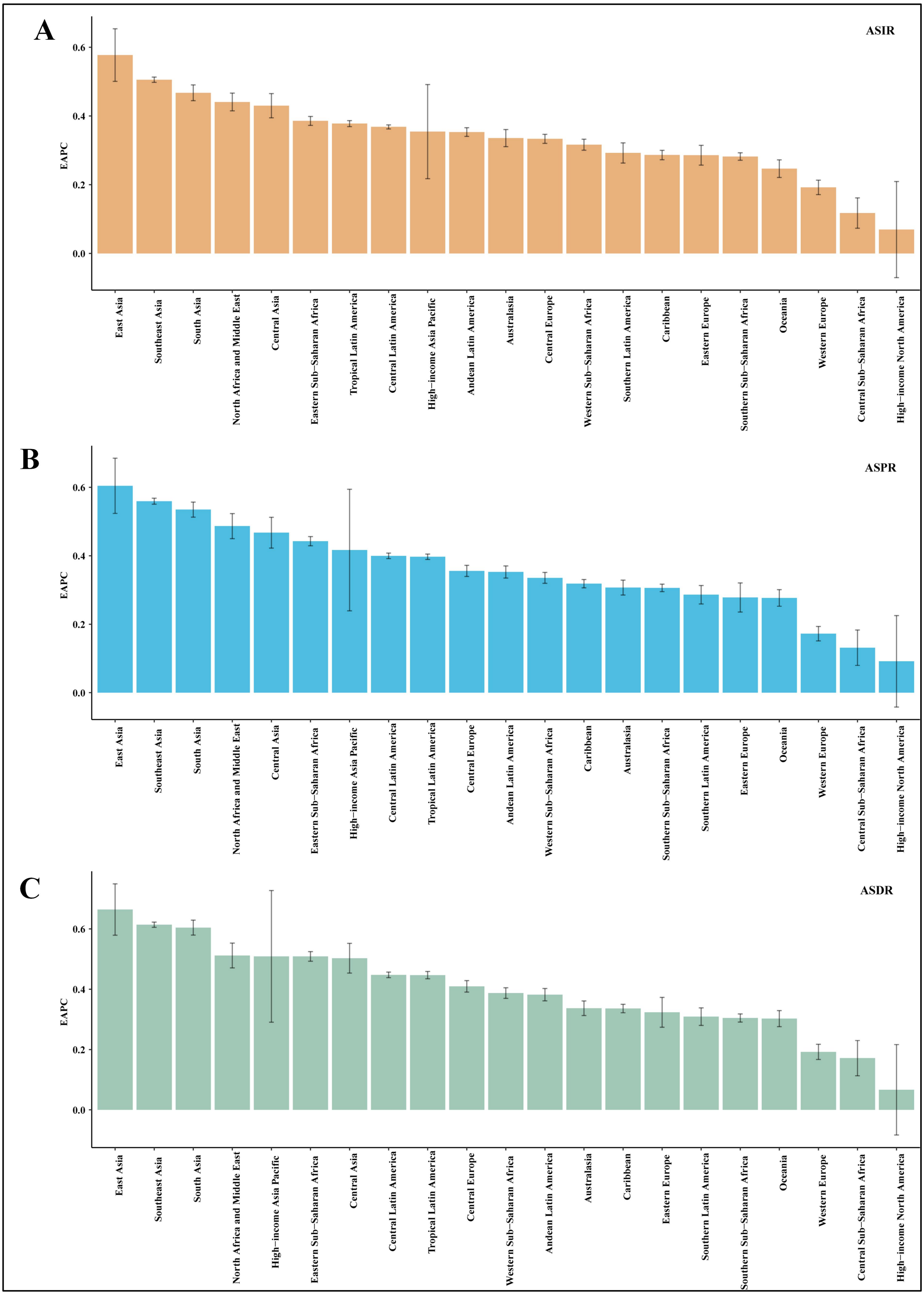


**Figure S2.** EAPCs of the ASRs for osteoarthritis in 21 regions. **(A)** EAPCs of the ASIR. **(B)** EAPCs of the ASPR. **(C)** EAPCs of the ASDR. ASR age-standardized rate; ASIR: age-standarzied incidence rate; ASPR: age-standarzied prevalence rate; ASDR: age-standardized disability-adjusted life years rate. EAPC estimated annual percentage change.

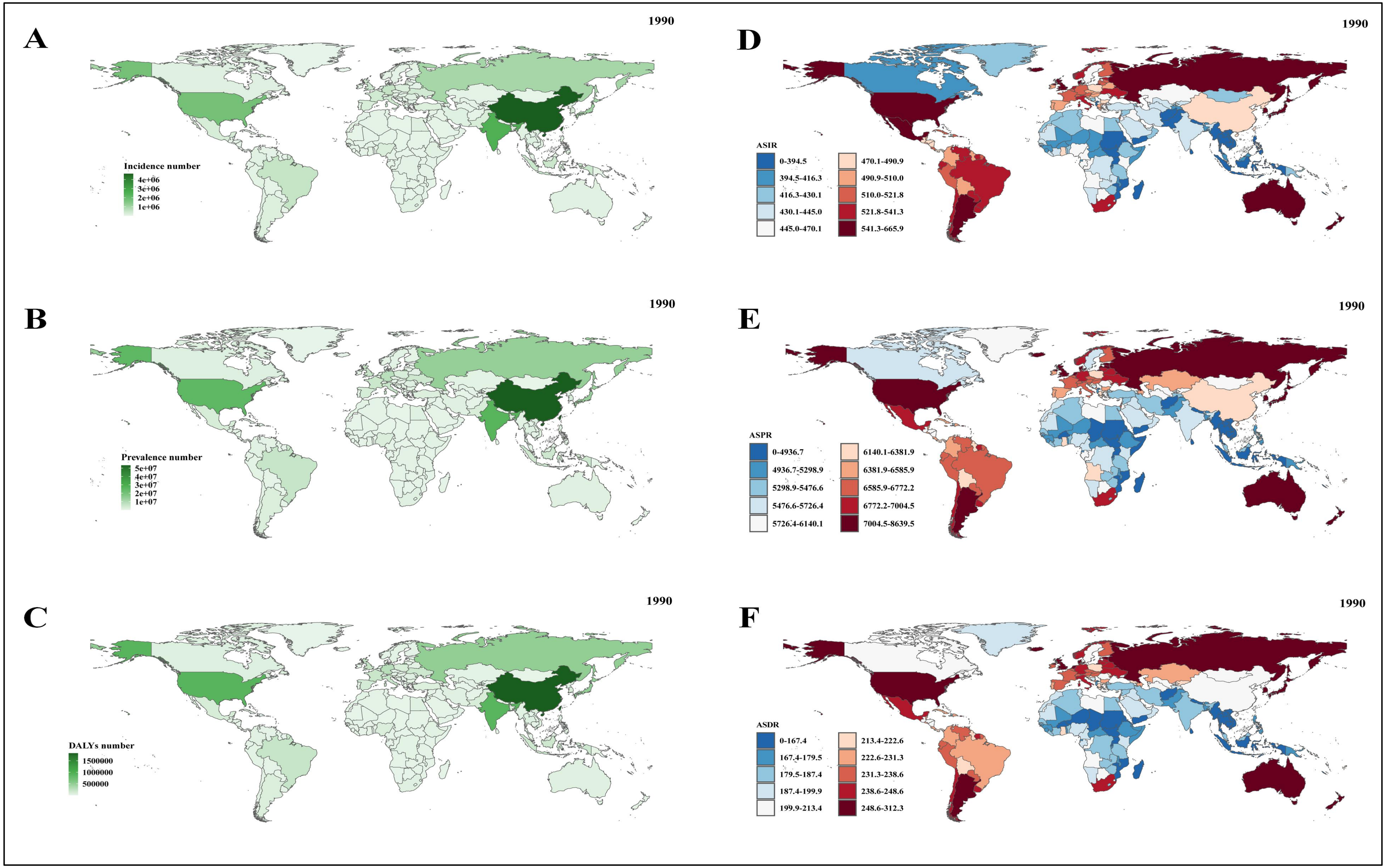


**Figure S3.** The incident case, prevalent case, DALYs, and ASRs (per 100,000 population) of osteoarthritis at the national level in 1990. **(A)** Incident case. **(B)** Prevalent case. **(C)** DALYs number. **(D)** ASIR. **(E)** ASPR. **(F)** ASDR. DALYs: disability-adjusted life years rate; ASRs: age-standardized rates; ASIR: age-standardized incidence rate; ASPR: age-standardized prevalence rate; ASDR: age-standardized disability-adjusted life years rate.

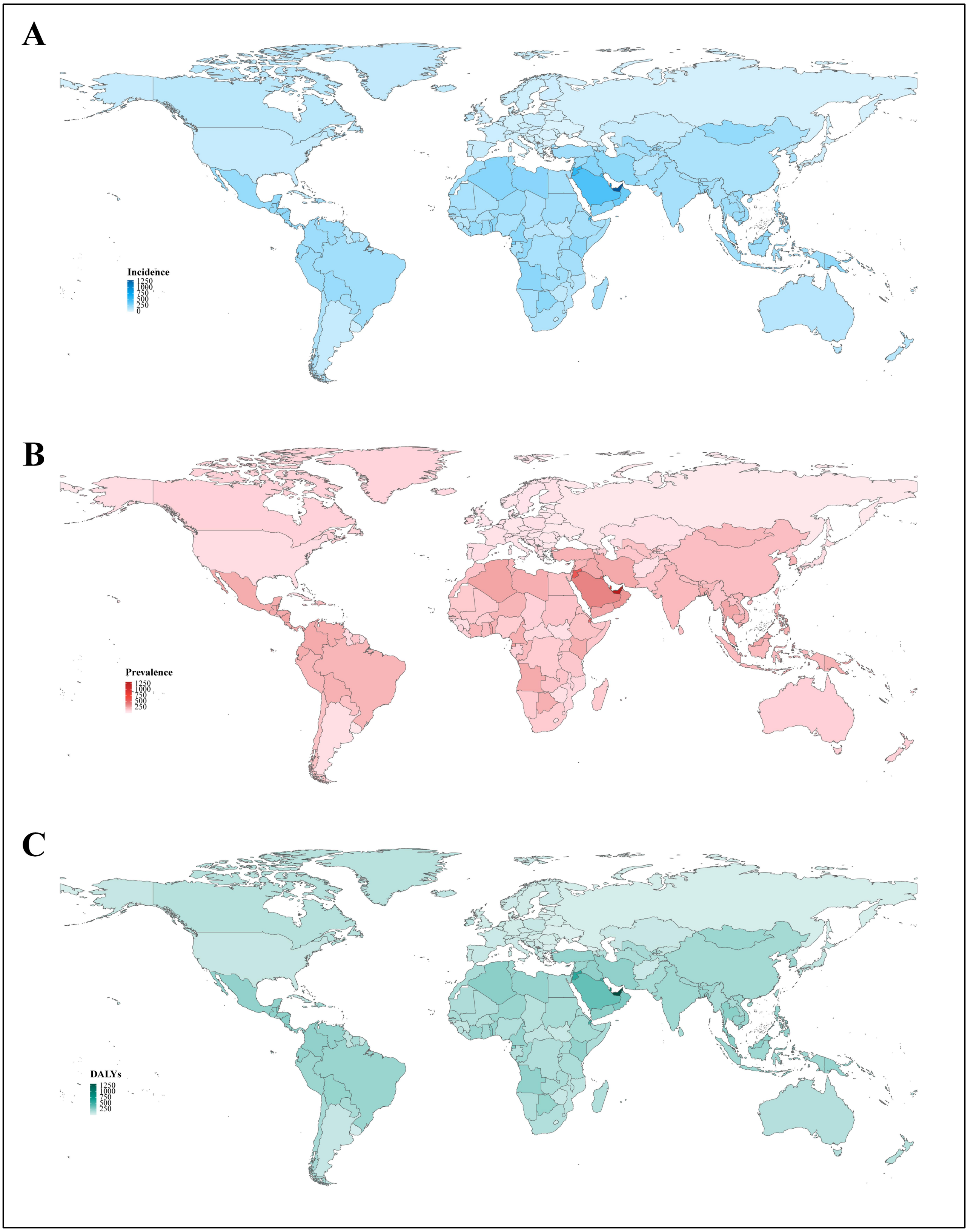


**Figure S4.** The incresing changes in incident cases, prevalent cases, and DALYs of osteoarthritis at the national level from 1990 to 2021. **(A)** Incident case. **(B)** Prevalent case. **(C)** DALYs number. DALYs: disability-adjusted life years rate.

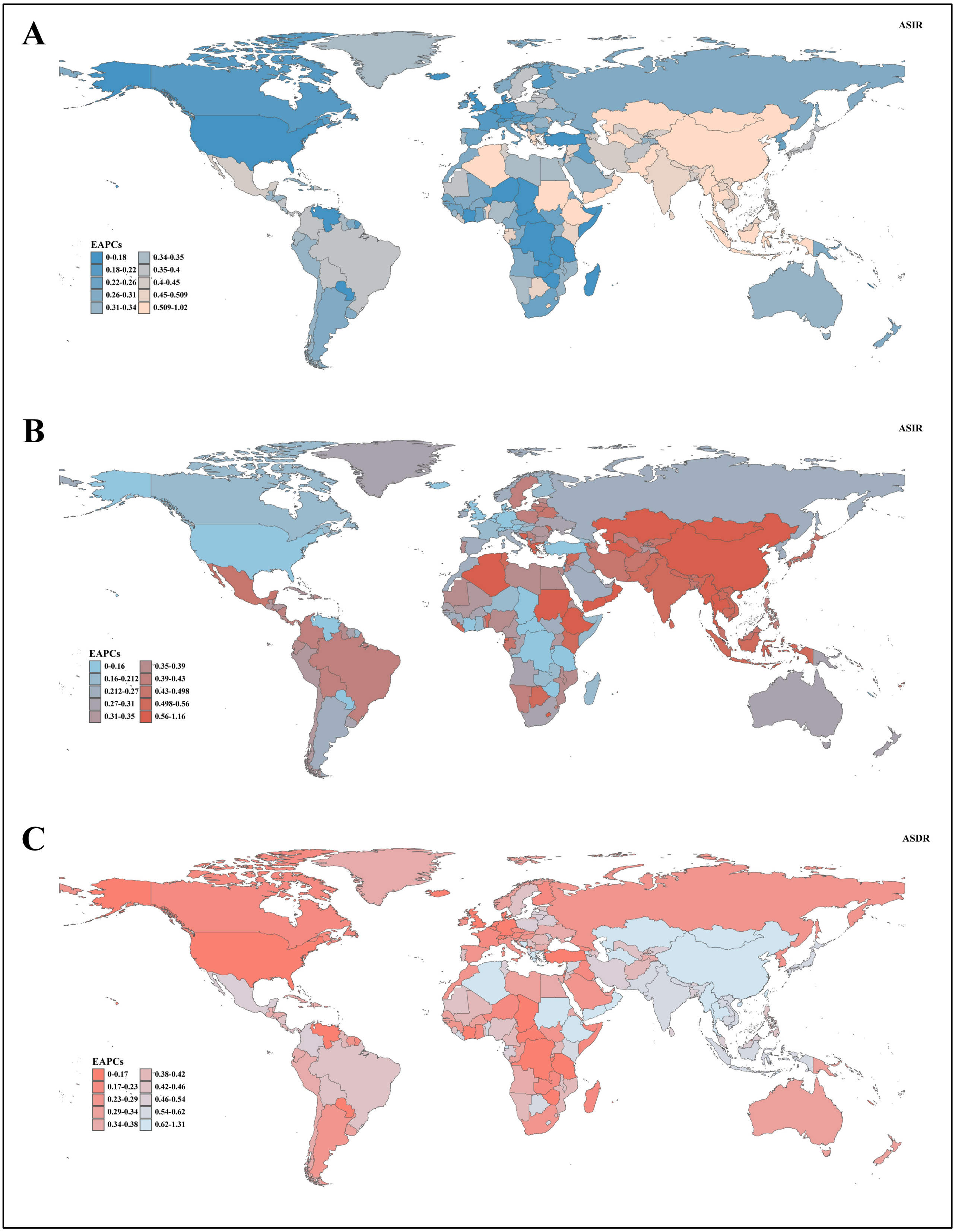


**Figure S5.** The EAPCs of ASRs of osteoarthritis at the national level from 1990 to 2021. **(A)** ASIR. **(B)** ASPR. **(C)** ASDR. EAPC: estimated annual percentage change; DALYs: disability-adjusted life years rate; ASRs: age-standardized rates; ASIR: age-standardized incidence rate; ASPR: age-standardized prevalence rate; ASDR: age-standardized disability-adjusted life years rate.

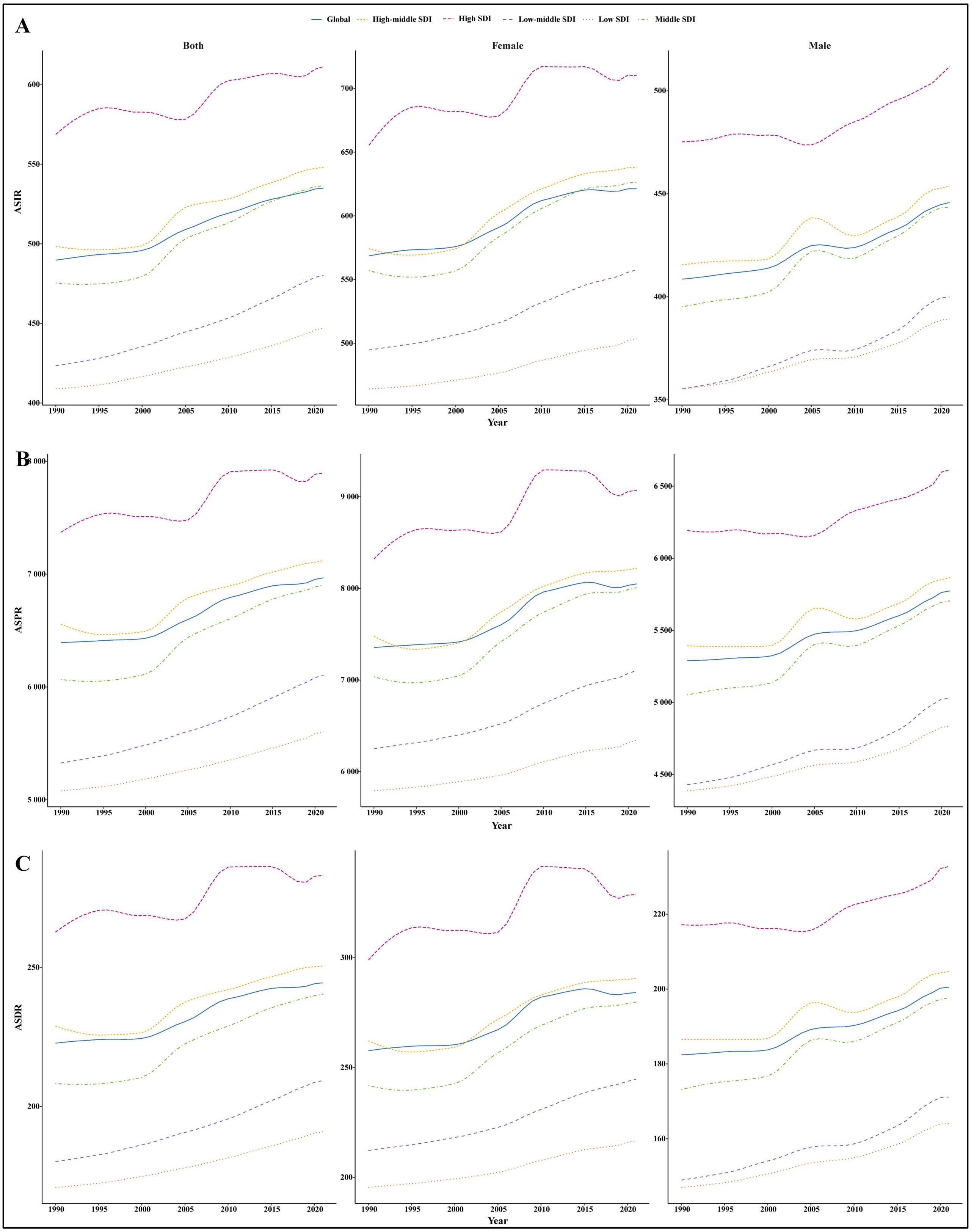


**Figure S6.** The ASRs (per 100,000 population) of osteoarthritis by sex at the global and SDI quintile levels from 1990 to 2021. **(A)** ASIR. **(B)** ASPR. **(C)** ASDR. SDI: socio-demographic index; ASRs: age-standardized rates; ASIR: age-standardized incidence rate; ASPR: age-standardized prevalence rate; ASDR: age-standardized disability-adjusted life years rate.

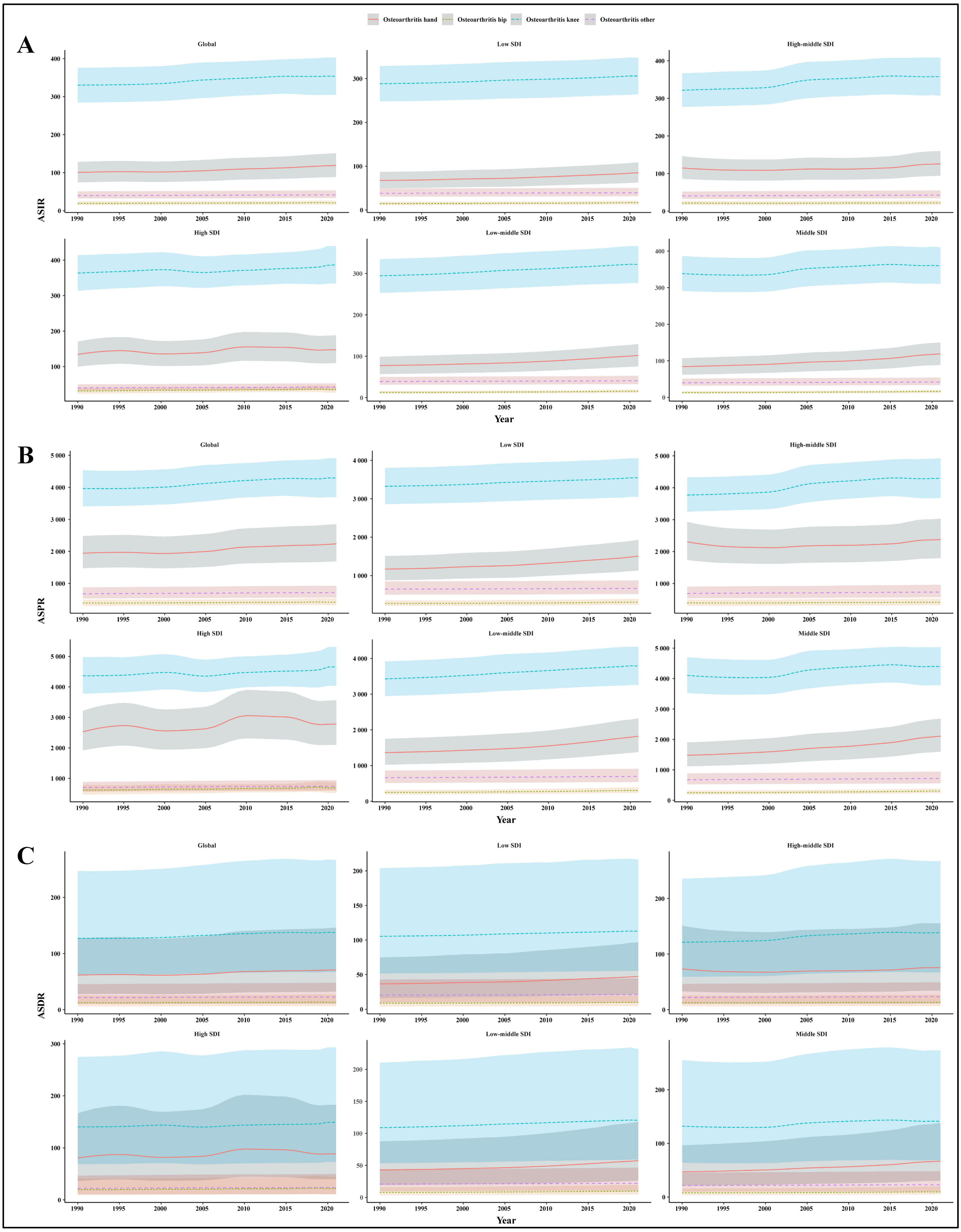


**Figure S7.** The ASRs (per 100,000 population) of four types of osteoarthritis among both sexes at the global and SDI quintile levels from 1990 to 2021. **(A)** ASIR. **(B)** ASPR. **(C)** ASDR. SDI: socio-demographic index; ASRs: age-standardized rates; ASIR: age-standardized incidence rate; ASPR: age-standardized prevalence rate; ASDR: age-standardized disability-adjusted life years rate.

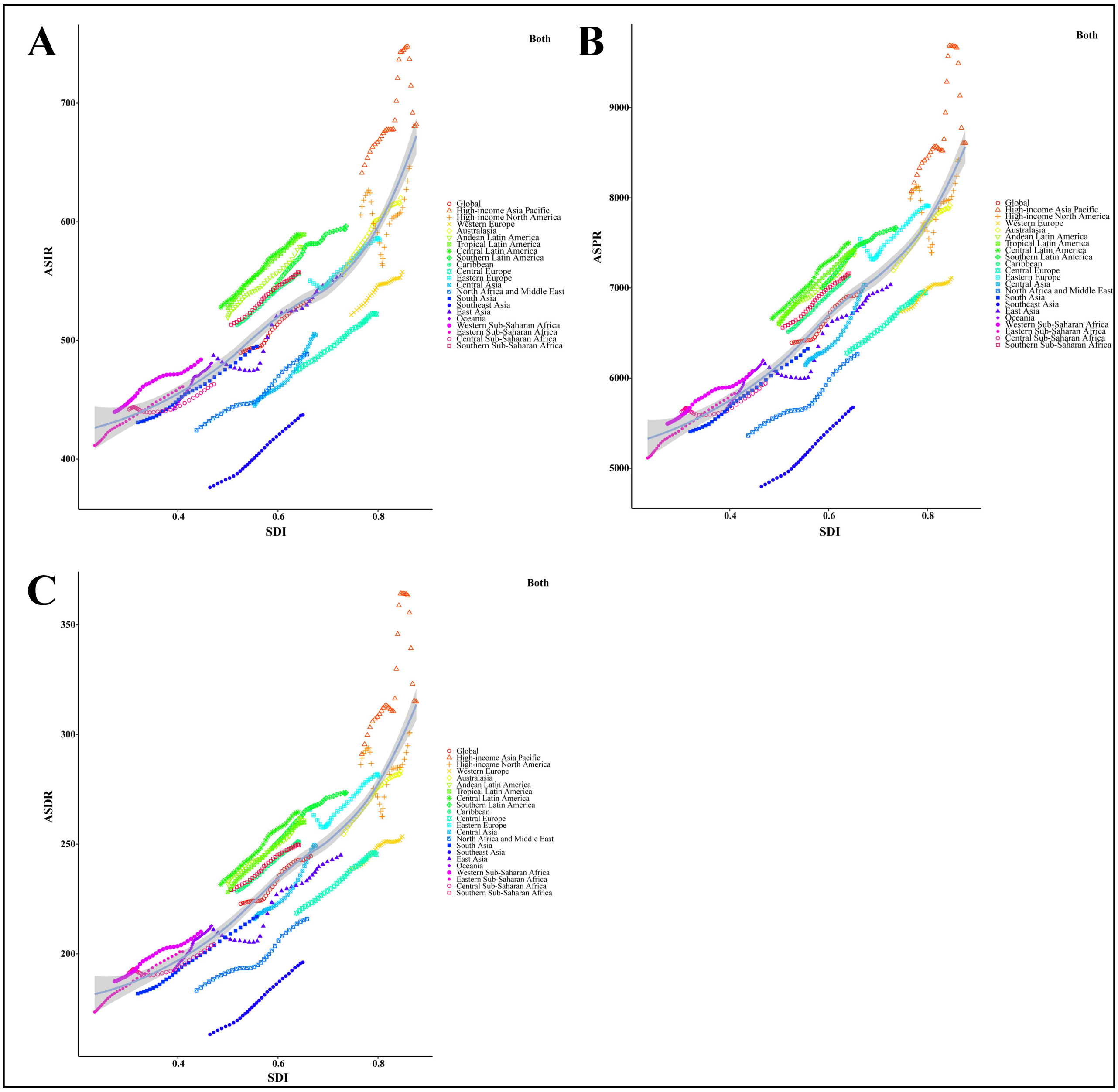


**Figure S8.** Correlation between SDI and ASRs of osteoarthritis among both sexes at the global and regional levels. **(A)** ASIR, r=0.7794, *P*<0.001. **(B)** ASPR, r=0.8353, *P*<0.001. **(C)** ASDR, r=0.8479, *P*<0.001. Colored lines show global and regional values for ASRs, and each point in a line represents one year. Expected values based on the SDI and ASRs in all locations are shown as the line. SDI: socio-demographic index; ASRs: age-standardized rates; ASIR: age-standardized incidence rate; ASPR: age-standardized prevalence rate; ASDR: age-standardized disability-adjusted life years rate.

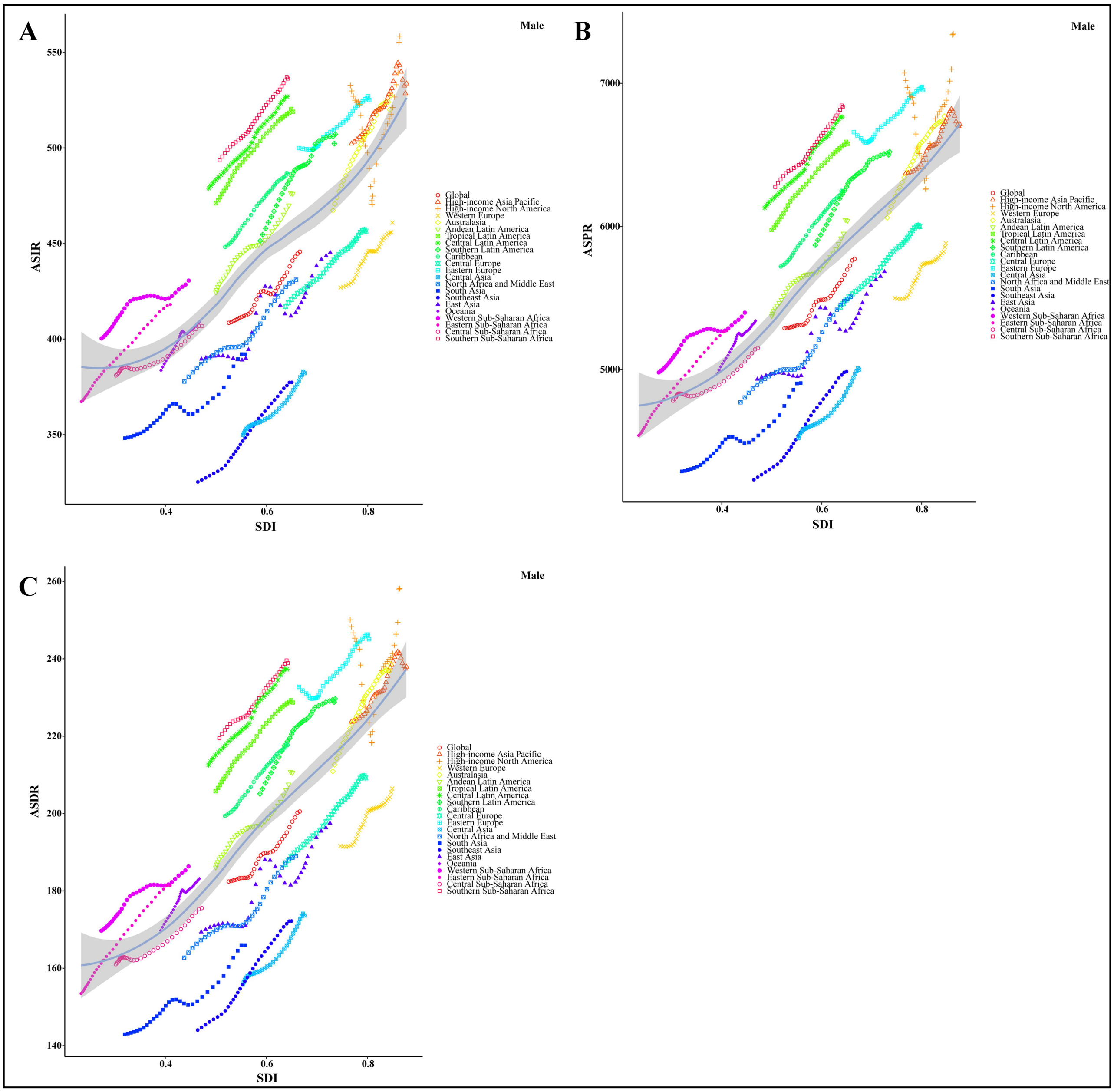


**Figure S9.** Correlation between SDI and ASRs of osteoarthritis among males at the global and regional levels. **(A)** ASIR, r=0.6503, *P*<0.001. (B) ASPR, r=0.6956, *P*<0.001. (C) ASDR, r=0.7111, *P*<0.001. Colored lines show global and region values for ASRs, and each point in a line represents one year. Expected values based on the SDI and ASRs in all locations are shown as the line. SDI, socio-demographic index; ASRs, age-standardized rates; SIR, age-standardized incidence rate; ASPR, age-standardized prevalence rate; ASDR, age-standardized disability-adjusted life years rate.

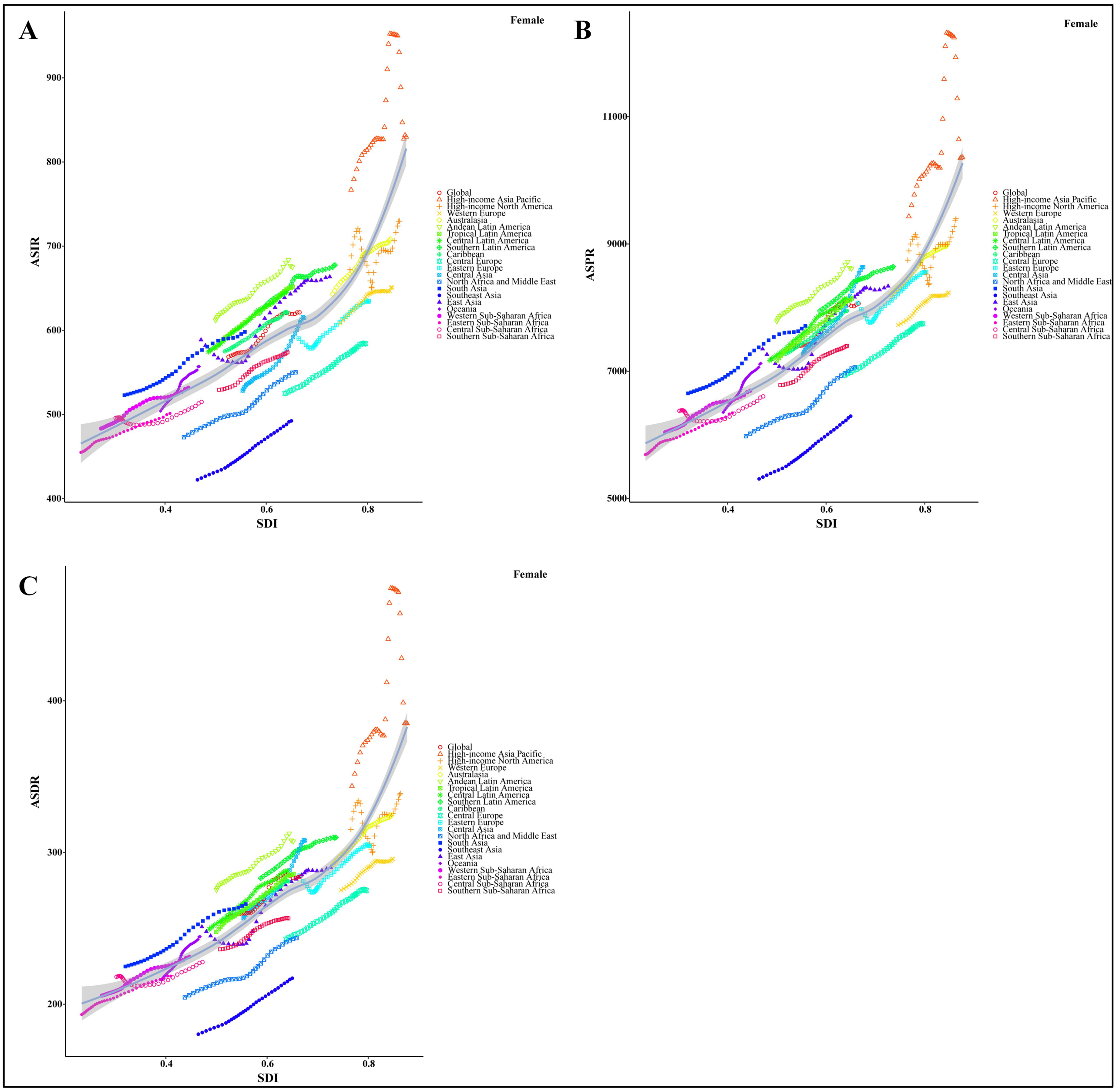


**Figure S10.** Correlation between SDI and ASRs of osteoarthritis among females at the global and regional levels. **(A)** ASIR, r=0.7880, *P*<0.001. **(B)** ASPR, r=0.8371, *P*<0.001. **(C)** ASDR, r=0.8477, *P*<0.001. Colored lines show global and region values for ASRs, and each point in a line represents one year. Expected values based on the SDI and ASRs in all locations are shown as the line. SDI, socio-demographic index; ASRs, age-standardized rates; ASIR, age-standardized incidence rate; ASPR, age-standardized prevalence rate; ASDR, age-standardized disability-adjusted life years rate

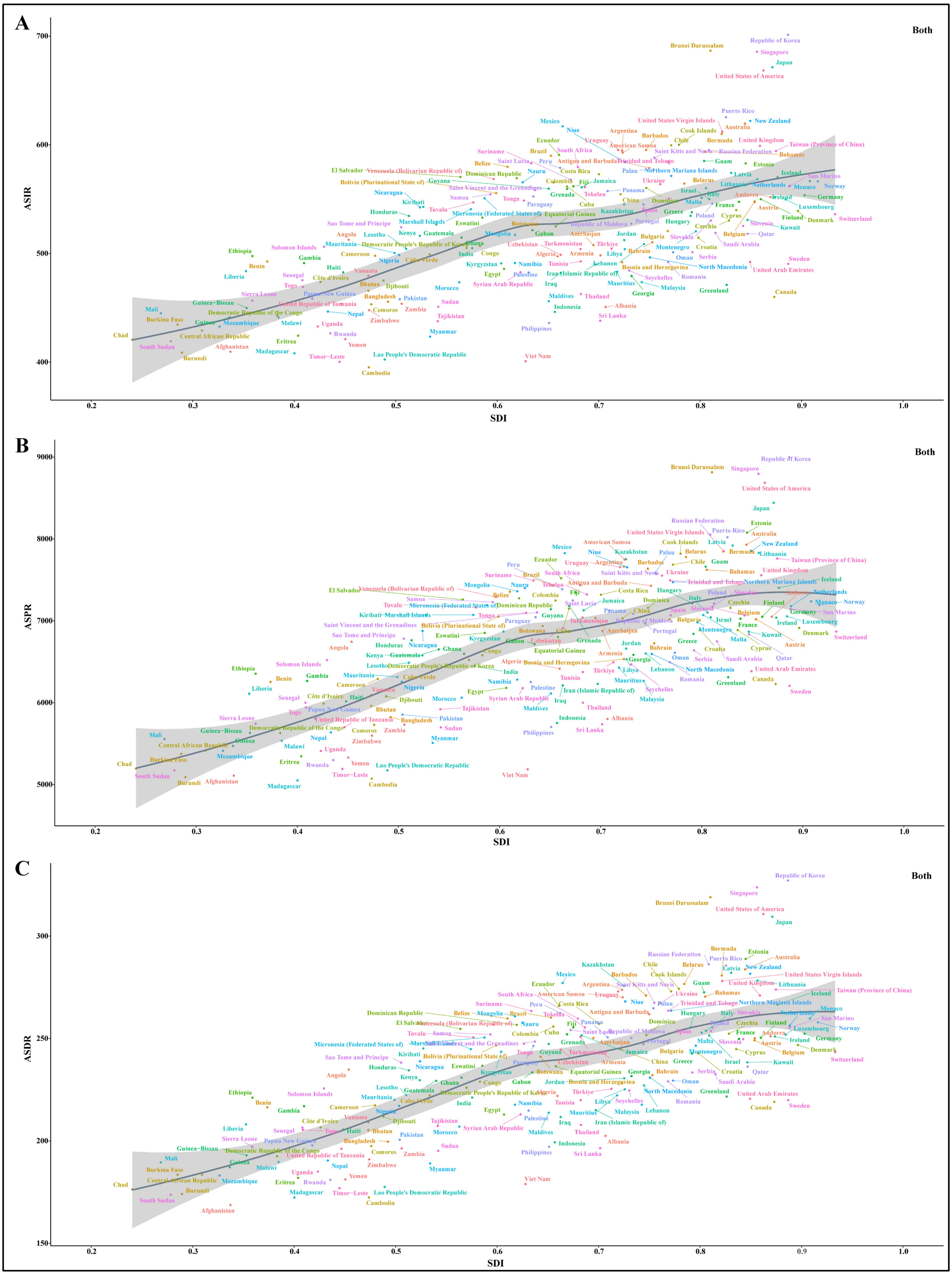


**Figure S11.** Correlation between SDI and ASRs of osteoarthritis among both sexes at the national level. **(A)** ASIR, r=0.6396, *P*<0.001. **(B)** ASPR, r=0.6687, *P*<0.001. **(C)** ASDR, r=0.7029, *P*<0.001. Each colored dot above represents a country, and the line represents the average expected value based on the SDI and ASRs in 204 countries and territories. SDI, socio-demographic index; ASRs, age-standardized rates; ASIR, age-standardized incidence rate; ASPR, age-standardized prevalence rate; ASDR, age-standardized disability-adjusted life years rate.

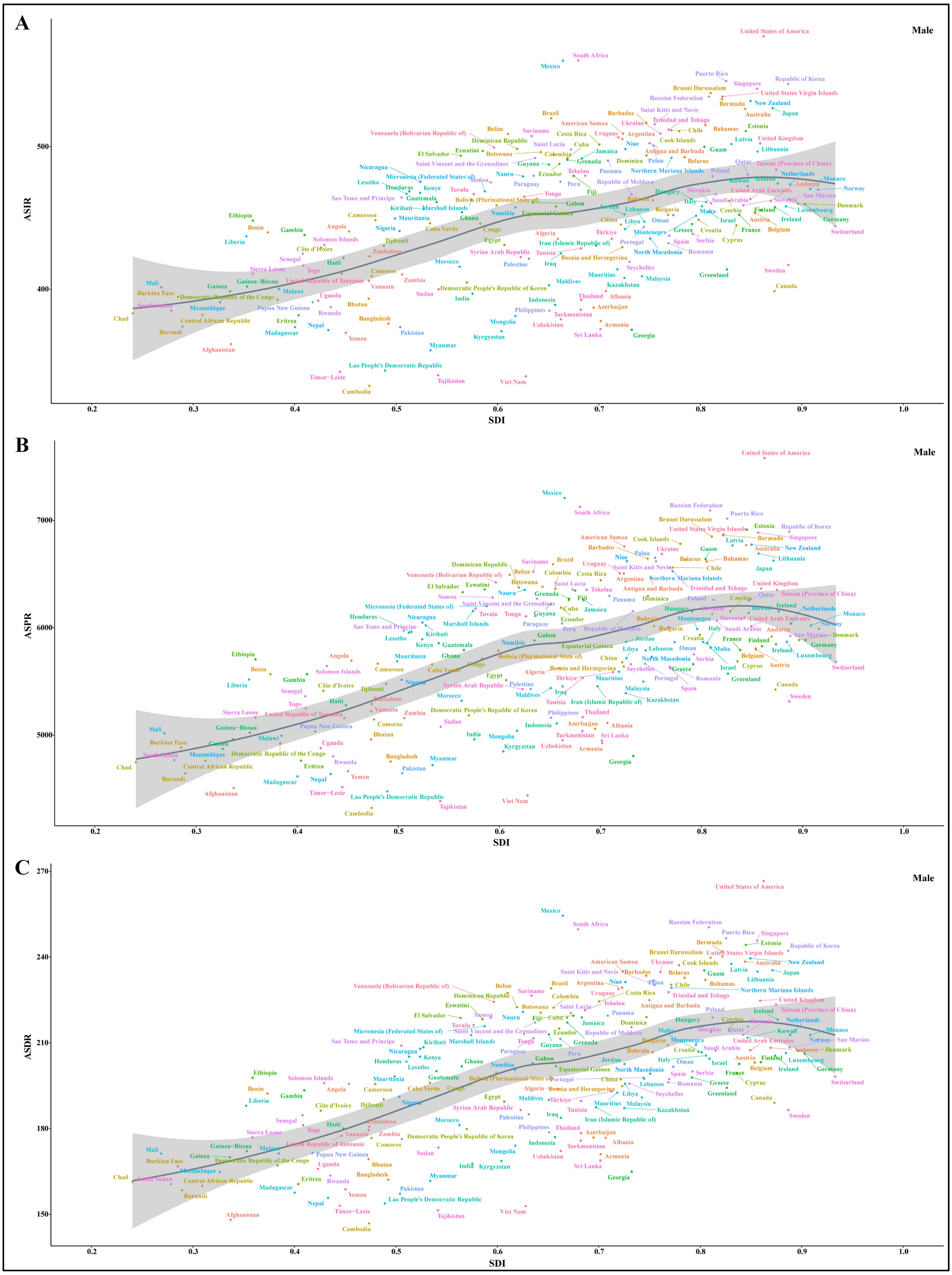


**Figure S12.** Correlation between SDI and ASRs osteoarthritis among males at the national level. **(A)** ASIR, r=0.5402, *P*<0.001. **(B)** ASPR, r=0.5905, *P*<0.001. **(C)** ASDR, r=0.6171, *P*<0.001. Each colored dot above represents a country, and the line represents the average expected value based on the SDI and ASRs in 204 countries and territories. SDI, socio-demographic index; ASRs, age-standardized rates; ASIR, age-standardized incidence rate; ASPR, age-standardized prevalence rate; ASDR, age-standardized disability-adjusted life years rate.

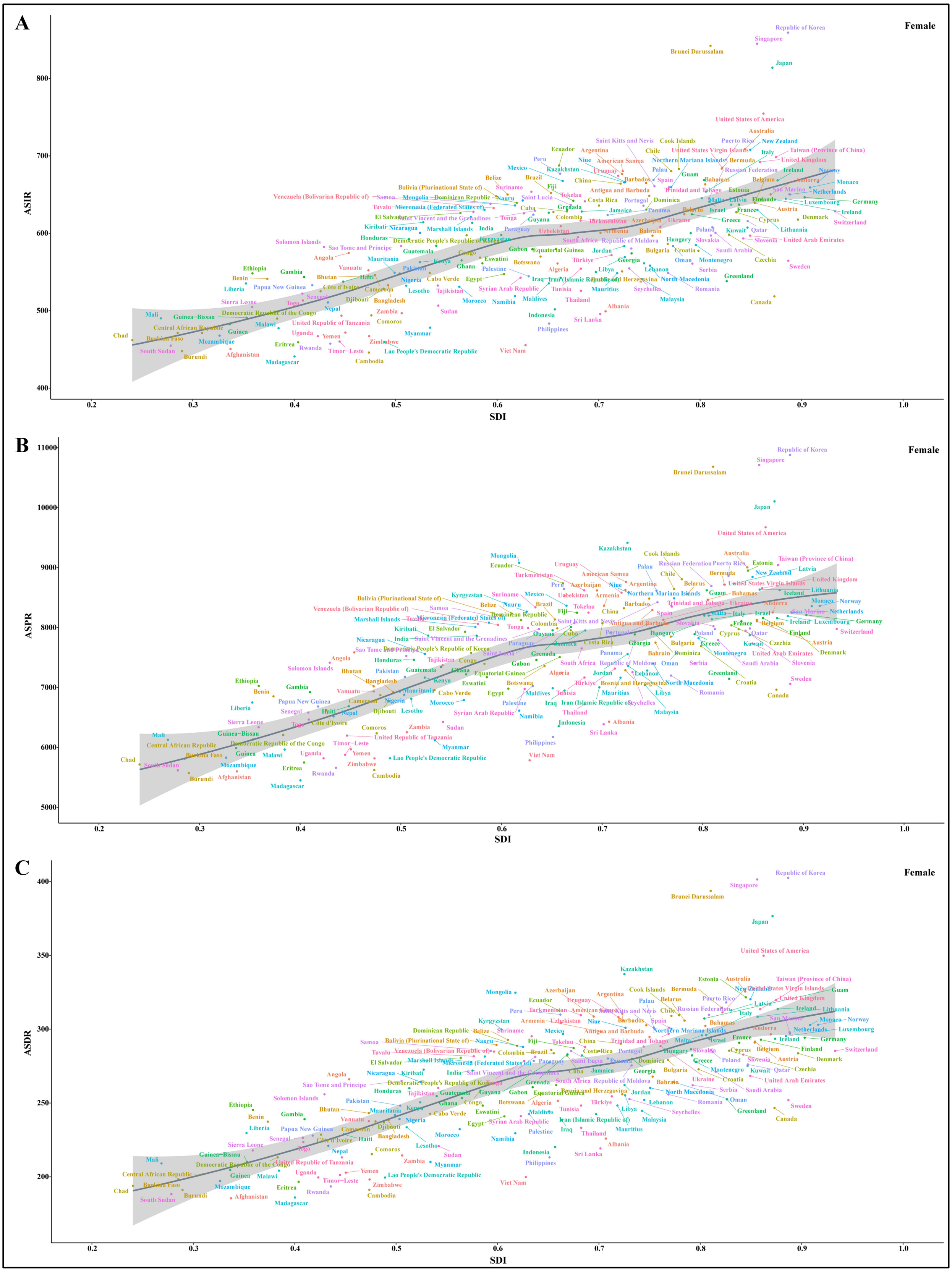


**Figure S13.** Correlation between SDI and ASRs of osteoarthritis among females at the national level. **(A)** ASIR, r=0.7014, *P*<0.001. **(B)** ASPR, r=0.7127, *P*<0.001. **(C)** ASDR, r=0.7450, *P*<0.001. Each colored dot above represents a country, and the line represents the average expected value based on the SDI and ASRs in 204 countries and territories. SDI, socio-demographic index; ASRs, age-standardized rates; ASIR, age-standardized incidence rate; ASPR, age-standardized prevalence rate; ASDR, age-standardized disability-adjusted life years rate.

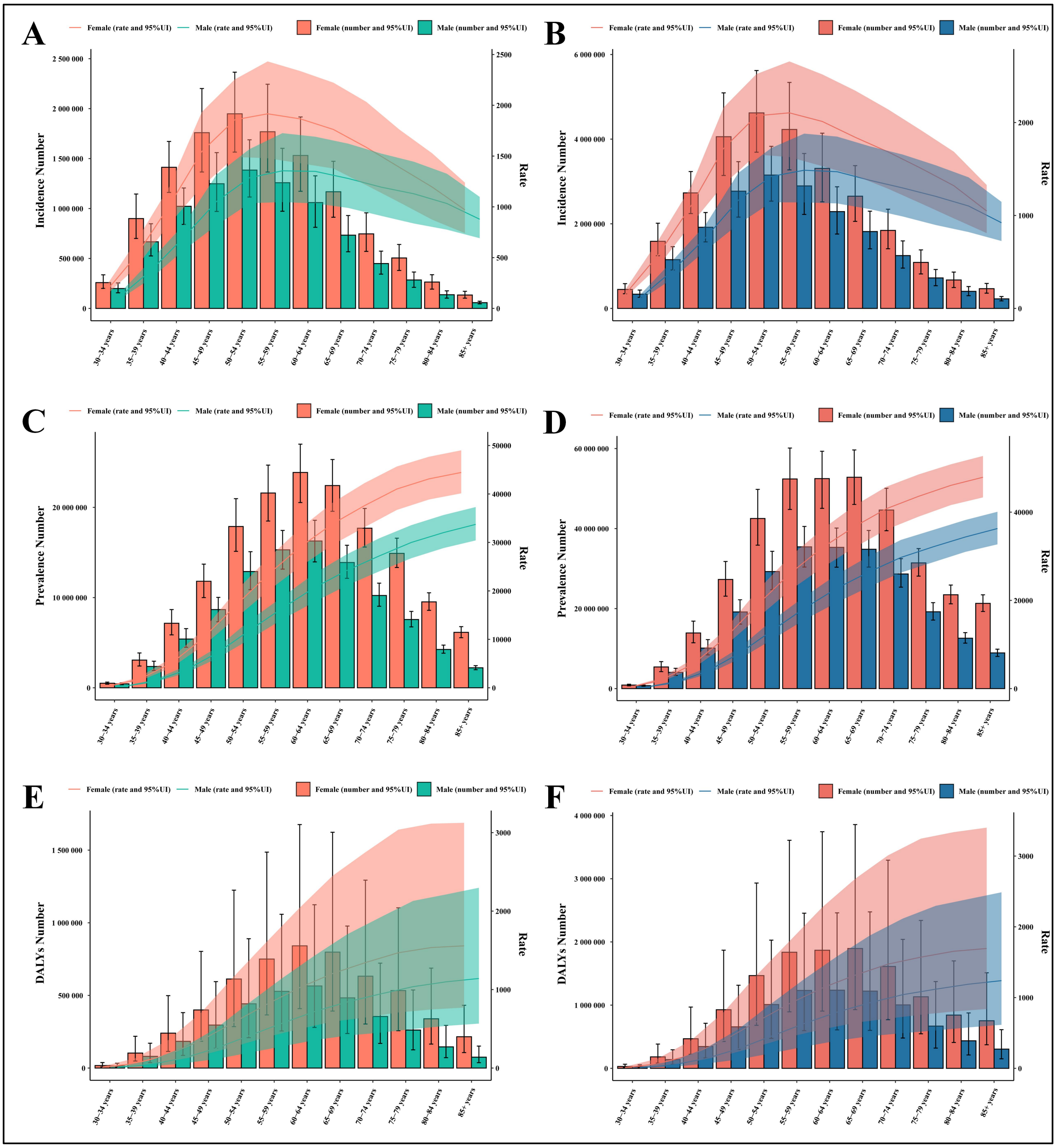


**Figure S14**. The global disease burden of osteoarthritis by age and sex in 1990 and 2021. **(A-B)** Incident case and incidence rate. **(C-D)** Prevalent cases and prevalence rate. **(E-F)** DALYs and DALYs rate. Error bars and shadow bands indicate 95% uncertainty intervals. DALYs: disability-adjusted life years.

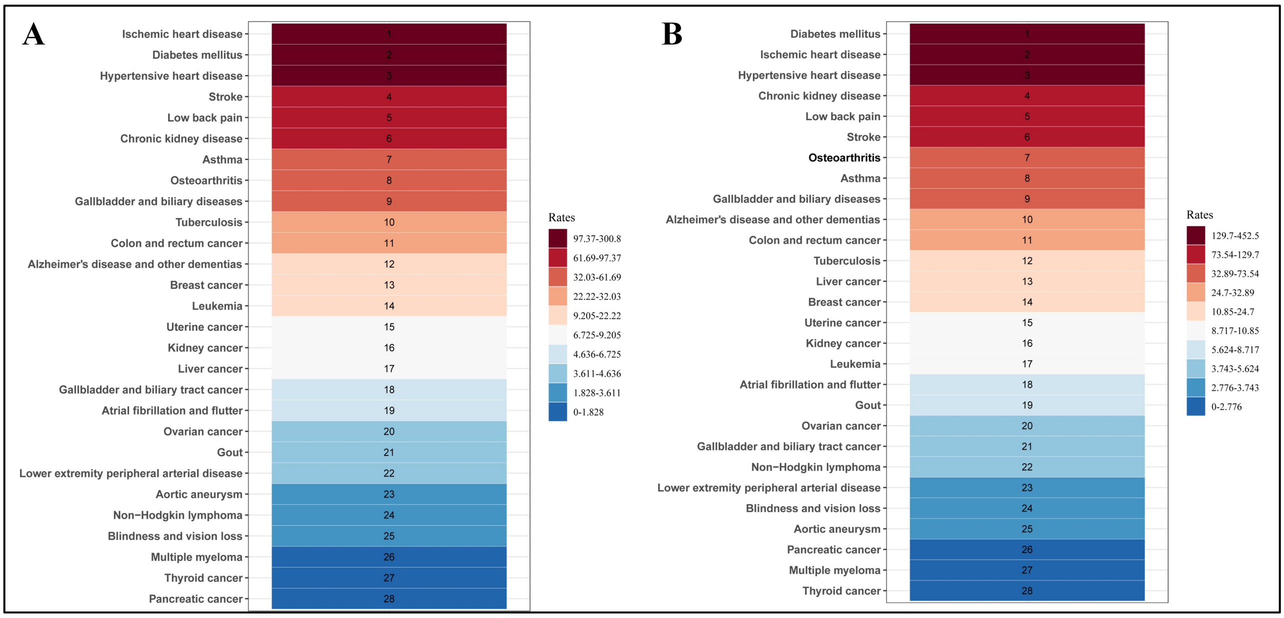


**Figure S15.** The ASDR ranking of diseases due to high BMI in 1990 and 2021. **(A)** 1990. **(B)** 2021. ASDR, age-standardized disability-adjusted life years rate.

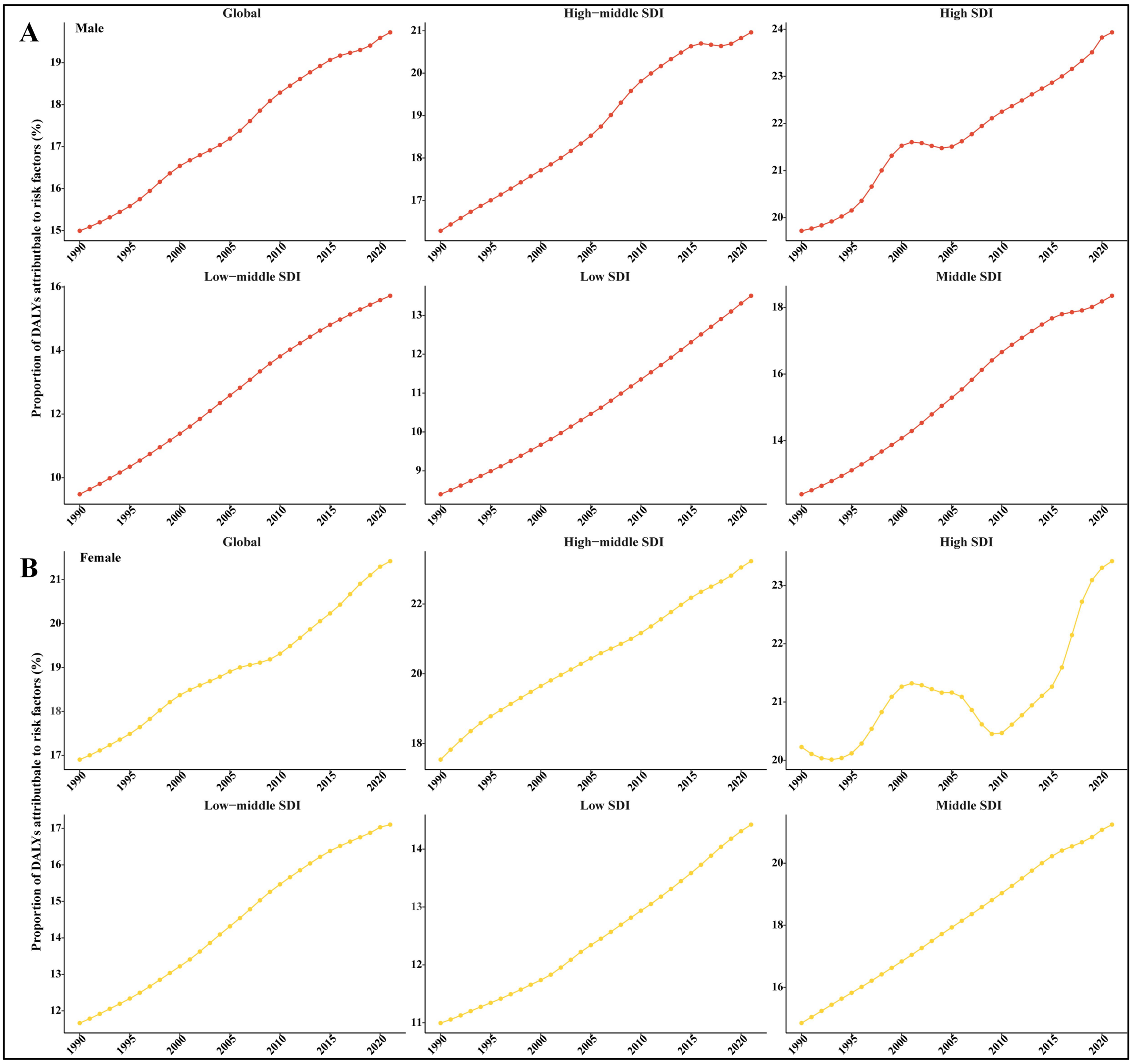


**Figure S16.** The proportion of DALYs of osteoarthritis attributable to high BMI among males and females at the global and SDI quintile levels from 1990 to 2021. **(A)** Males. **(B)** Females. DALYs: disability-adjusted life years; BMI: body-mass index; SDI: socio-demographic index.

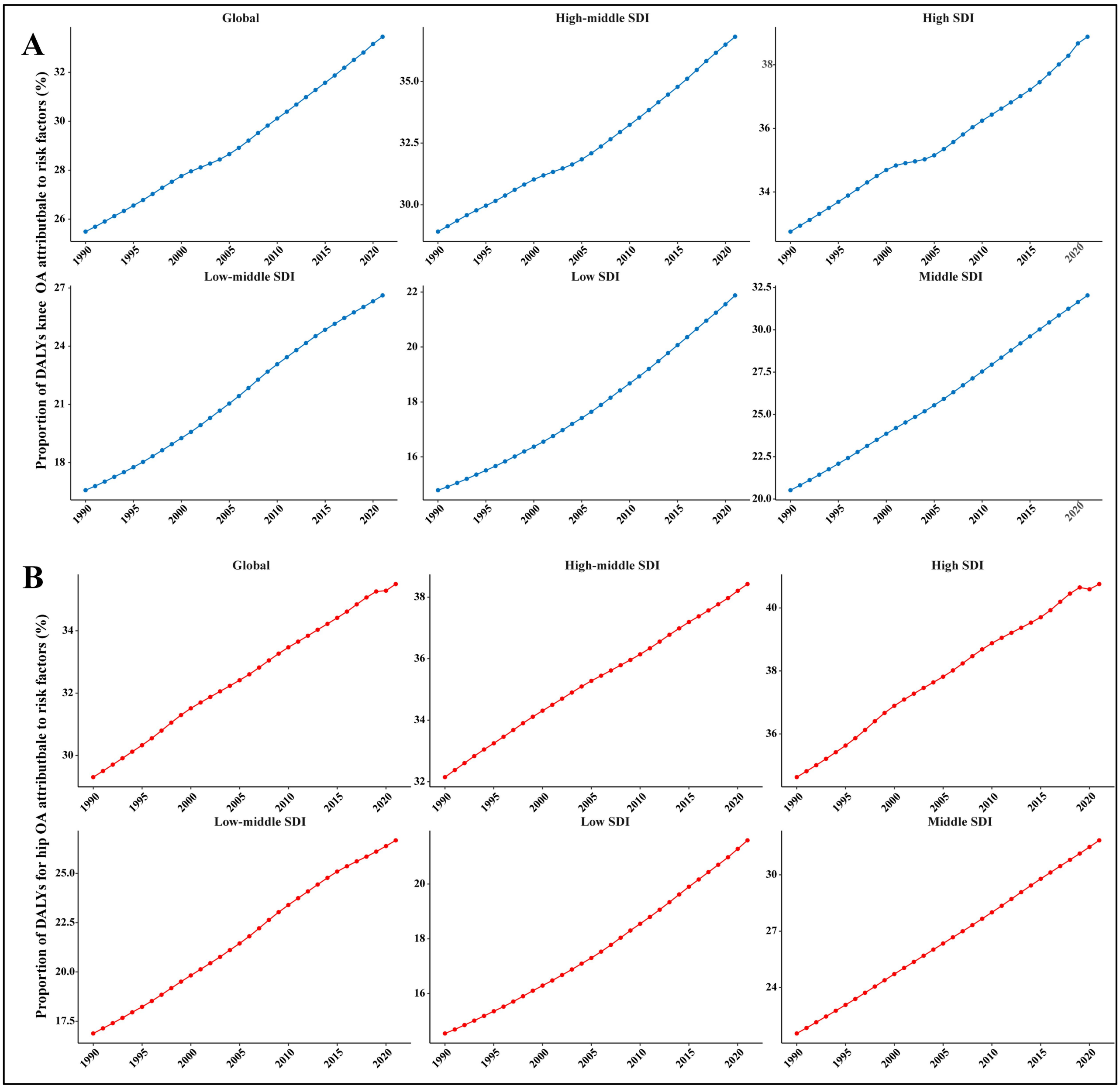


**Figure S17.** The proportion of DALYs of knee and hip osteoarthritis attributable to high BMI among both sexes at the global and SDI quintile levels from 1990 to 2021. **(A)** Knee osteoarthritis. **(B)** Hip osteoarthritis. DALYs: disability-adjusted life years; BMI: body-mass index; SDI: socio-demographic index.

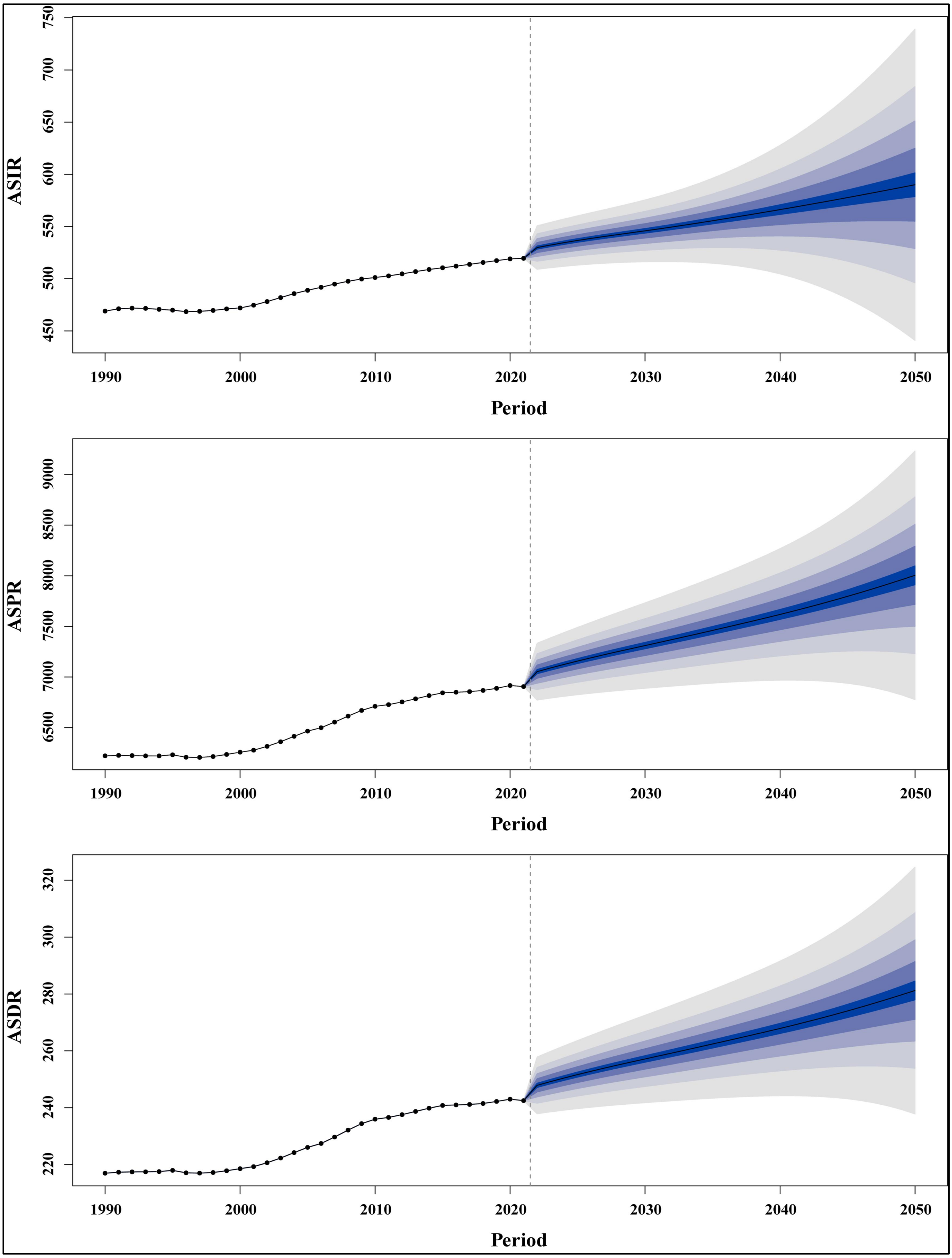


**Figure S18.** The projection of ASRs (per 100,000 population) of osteoarthritis globally from 2021 to 2050. DALYs: disability-adjusted life years rate; ASRs: age-standardized rates; ASIR: age-standardized incidence rate; ASPR: age-standardized prevalence rate; ASDR: age-standardized disability-adjusted life years rate.

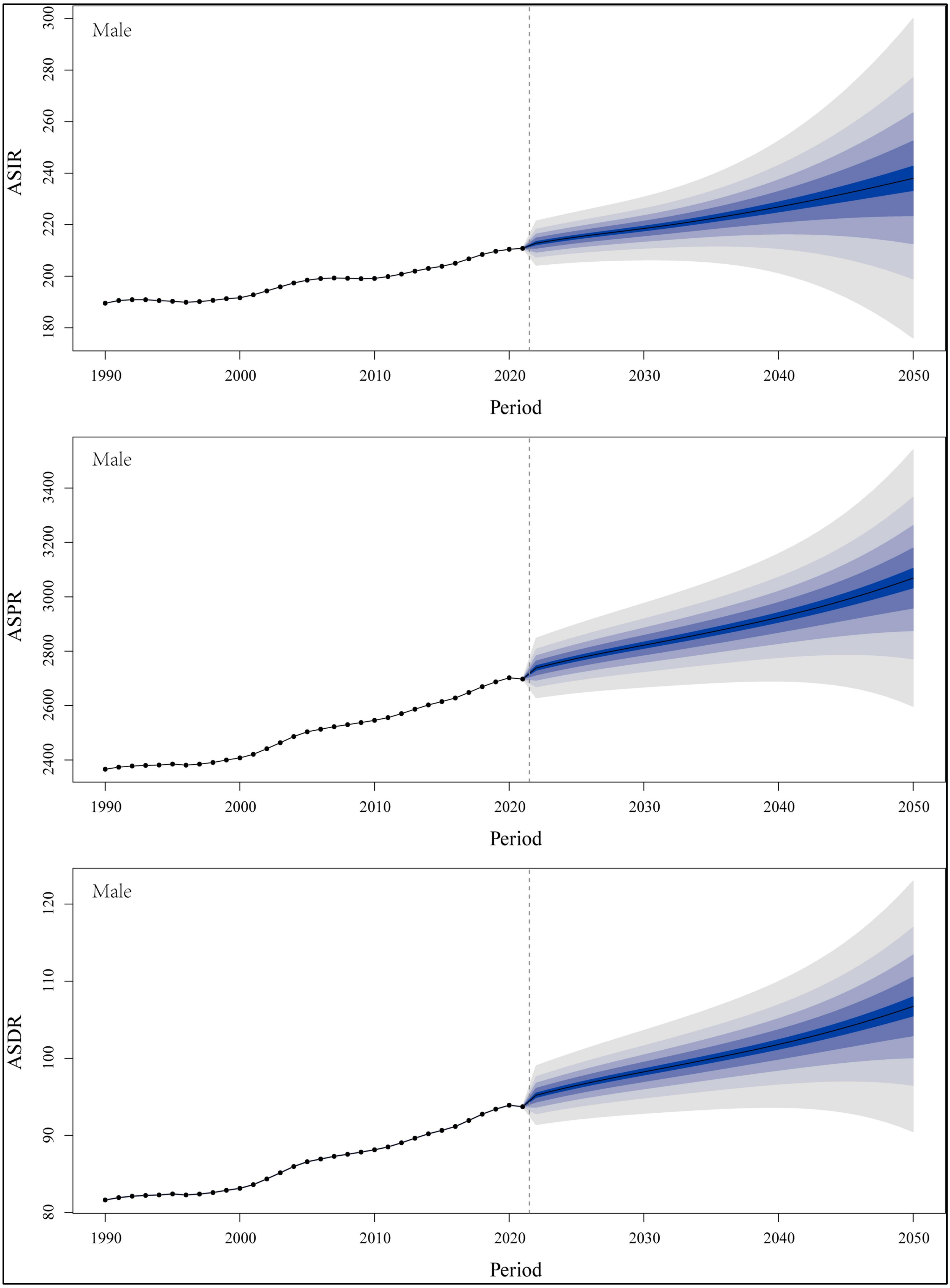


**Figure S19.** The projection of ASRs (per 100,000 population) of osteoarthritis among males globally from 2021 to 2050. DALYs: disability-adjusted life years rate; ASRs: age-standardized rates; ASIR: age-standardized incidence rate; ASPR: age-standardized prevalence rate; ASDR: age-standardized disability-adjusted life years rate.

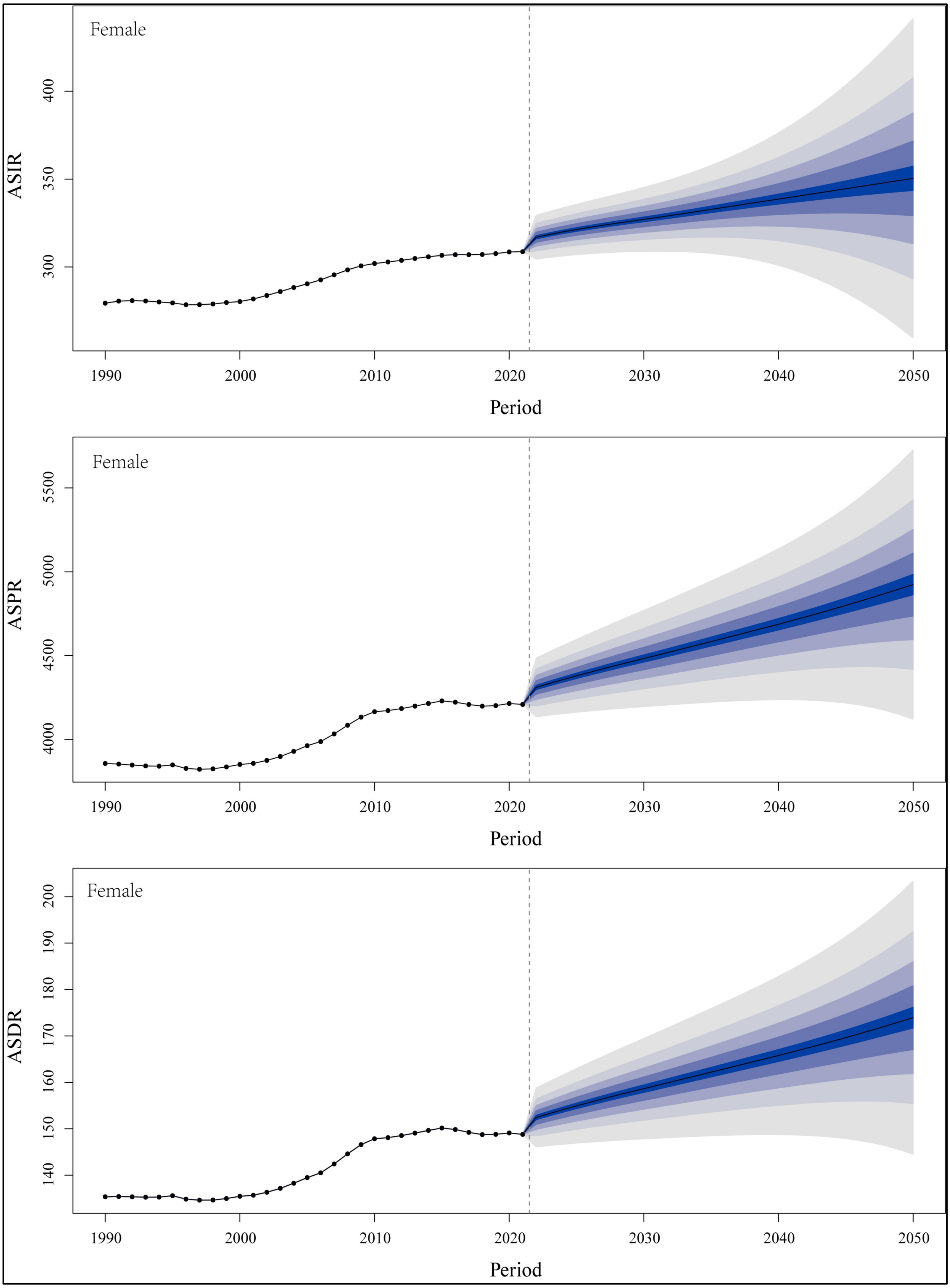


**Figure S20.** The projection of ASRs (per 100,000 population) of osteoarthritis among females globally from 2021 to 2050. DALYs: disability-adjusted life years rate; ASRs: age-standardized rates; ASIR: age-standardized incidence rate; ASPR: age-standardized prevalence rate; ASDR: age-standardized disability-adjusted life years rate.

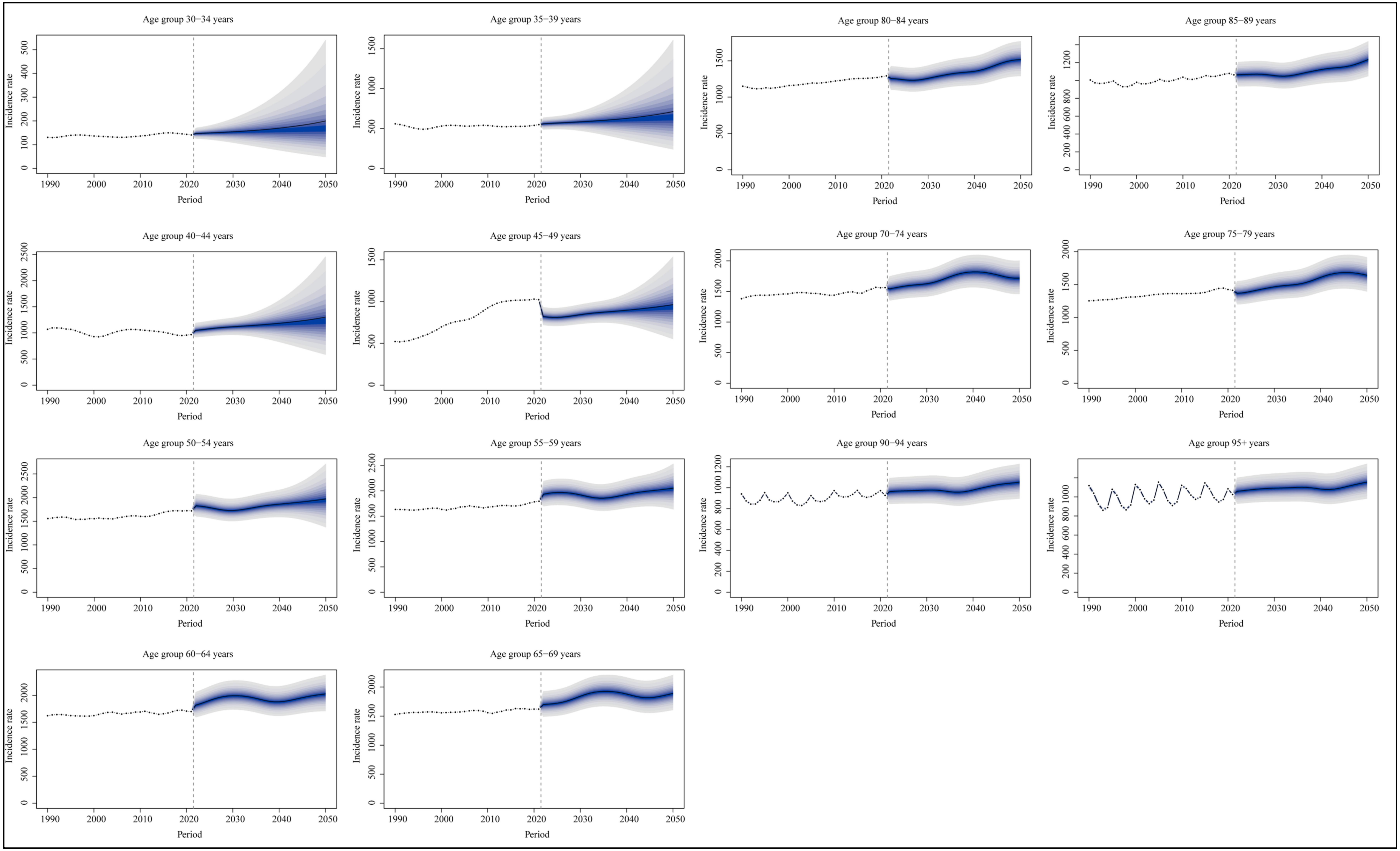


**Figure S21.** The projection of incidence rate (per 100,000 population) of osteoarthritis across different age groups globally from 2021 to 2050.

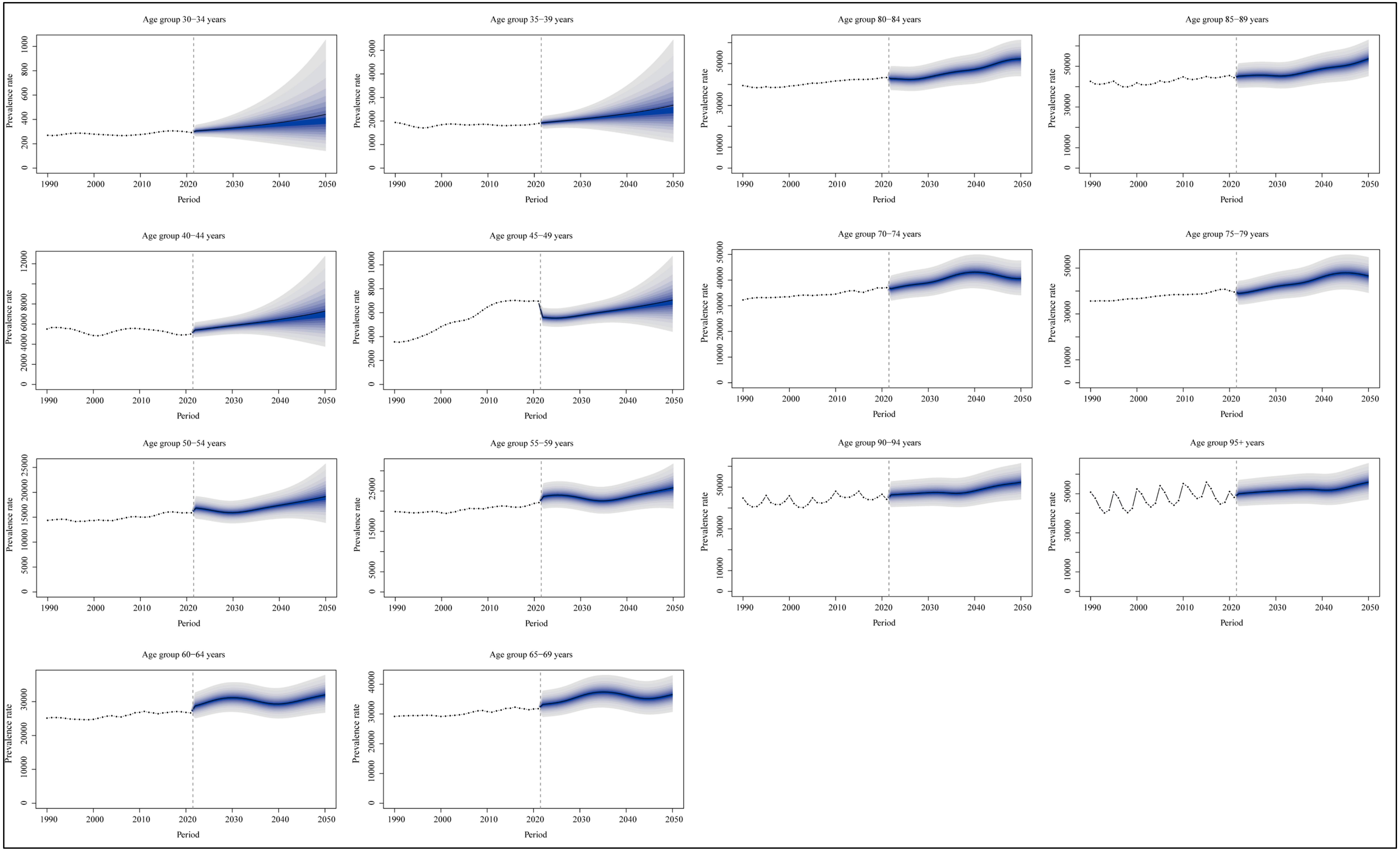


**Figure S22.** The projection of prevalence rate (per 100,000 population) of osteoarthritis across different age groups globally from 2021 to 2050.

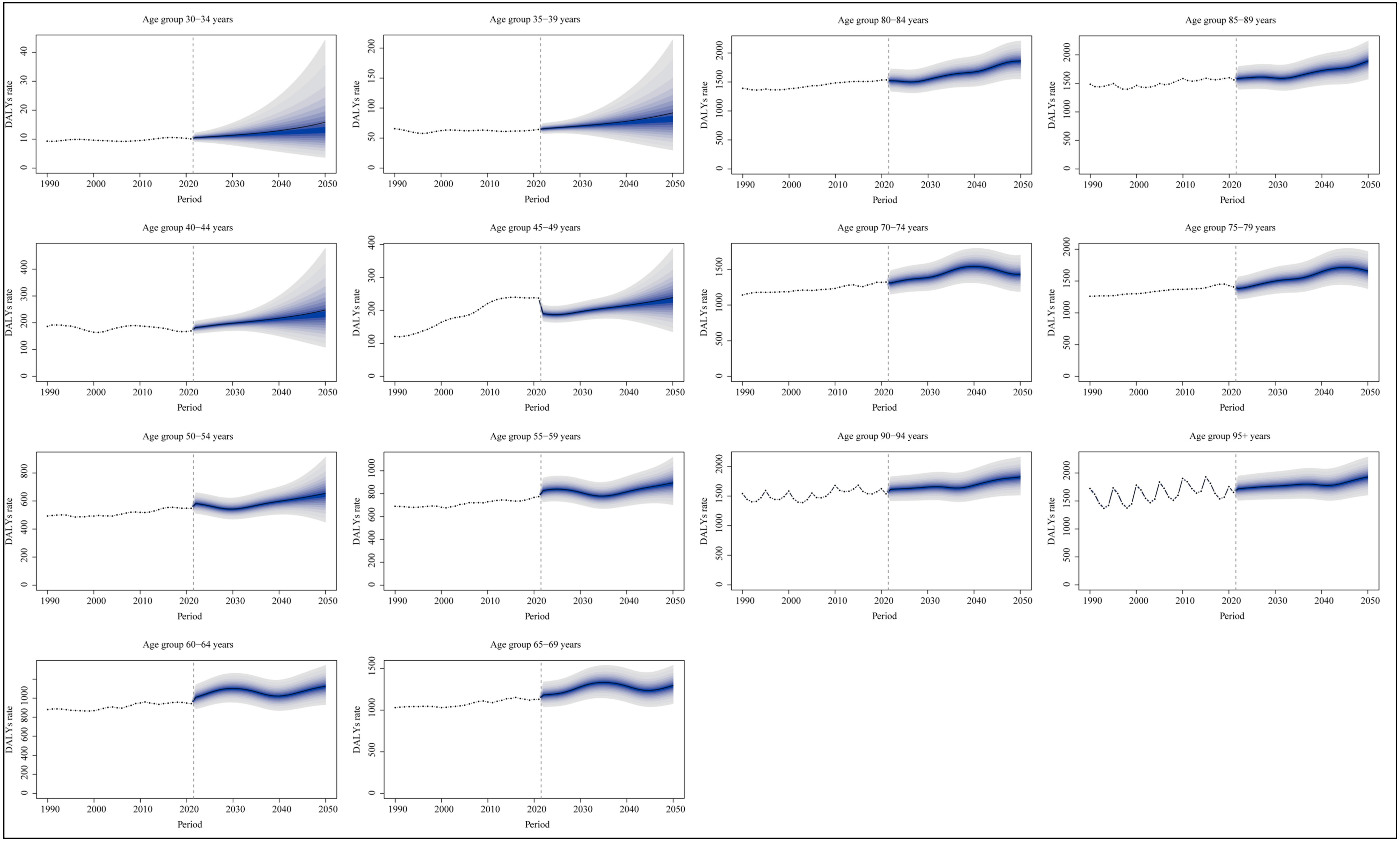


**Figure S23.** The projection of DALYs rate (per 100,000 population) of osteoarthritis across different age groups globally from 2021 to 2050. DALYs: disability-adjusted life years.

**Table S1.** The incident case, prevalent case, DALYs, and increasing changes in osteoarthritis from 1990 to 2021

| Characteristics | Incident case | | | Prevalent case | | | DALYs | | |
|---|---|---|---|---|---|---|---|---|---|
| | 1990 | 2021 | 1990-2021 | 1990 | 2021 | 1990-2021 | 1990 | 2021 | 1990-2021 |
| | Both (95%UI) | Both (95%UI) | Change (%, 95%UI) | Both (95%UI) | Both (95%UI) | Change (%, 95%UI) | Both (95%UI) | Both (95%UI) | Change (%, 95%UI) |
| **Global** | 20900510.49 (18467652.64-23104315.92) | 46632144.37 (41122052.73-51644430.77) | 123.11 (120.62-125.21) | 256076699.93 (227119748.27-283438464.92) | 606989319.00 (537873607.70-670519617.28) | 137.03 (135.49-138.71) | 8918856.67 (4264150.65-17983775.68) | 21304565.82 (10189161.47-42935419.74) | 138.87 (136.83-140.75) |
| **Sex** | | | | | | | | | |
| Male | 8503791.70 (7501087.56-9450602.28) | 18929482.73 (16687692.13-21023898.06) | 122.60 (119.90-125.06) | 99511973.72 (88583766.52-110775303.96) | 238385345.60 (211383648.34-265579452.04) | 139.55 (137.55-141.80) | 3433701.53 (1641848.61-6908410.21) | 8283306.36 (3957189.75-16670635.30) | 141.24 (138.69-143.89) |
| Female | 12396718.80 (10971066.63-13667494.85) | 27702661.64 (24480498.43-30566645.19) | 123.47 (120.93-125.59) | 156564726.21 (138991857.11-172606175.02) | 368603973.40 (326841458.56-407392577.82) | 135.43 (134.00-136.97) | 5485155.14 (2622686.94-11075365.47) | 13021259.45 (6241449.89-26272385.74) | 137.39 (135.46-139.26) |
| **Types** | | | | | | | | | |
| Osteoarthritis knee | 14134283.86 (12150932.91-16079380.86) | 30845890.76 (26534151.15-35188904.82) | 118.23 (115.28-120.72) | 159798909.45 (137277437.41-182882553.69) | 374738744.12 (321858981.94-428353219.52) | 134.51 (132.72-136.41) | 5145338.70 (2507481.29-9953258.48) | 12019069.90 (5858107.88-23267858.34) | 133.59 (131.71-135.70) |
| Osteoarthritis hand | 4282225.41 (3174006.24-5467718.71) | 10367241.35 (7686291.00-13251881.07) | 142.10 (136.56-146.64) | 76317645.29 (57748915.37-97654829.09) | 194284753.65 (146479864.83-248177667.64) | 154.57 (151.02-157.98) | 2432663.83 (1103099.91-5033222.20) | 6167307.93 (2803411.05-12737380.26) | 153.52 (149.79-156.97) |
| Osteoarthritis hip | 811983.72 (618955.85-1030152.49) | 1796781.41 (1368838.77-2284910.02) | 121.28 (116.45-126.36) | 15397009.44 (11798401.89-19637630.40) | 35886278.68 (27630665.50-45745600.75) | 133.07 (129.91-136.13) | 491843.53 (231455.91-994652.97) | 1139666.93 (535835.99-2306309.68) | 131.71 (128.41-135.13) |
| Osteoarthritis other | 1672017.50 (1342156.56-2140831.34) | 3622230.84 (2883048.46-4655164.14) | 116.64 (113.10-119.66) | 26561599.27 (20775159.95-34645571.57) | 62159810.96 (48515474.90-80998531.09) | 134.02 (131.81-135.90) | 849010.61 (392623.58-1779078.48) | 1978521.05 (913347.14-4152837.93) | 133.04 (130.60-135.36) |
| **SDI** | | | | | | | | | |
| High | 5864566.47 (5225613.42-6479489.06) | 10189235.97 (9088793.27-11269284.92) | 73.74 (71.42-76.18) | 79661768.00 (71442384.37-87921586.82) | 152390475.55 (136382302.92-168139763.52) | 91.30 (89.83-92.86) | 2845199.11 (1366268.18-5724654.95) | 5486878.63 (2648510.33-11052365.17) | 92.85 (91.02-94.96) |
| High-middle | 5133784.52 (4537917.93-5675082.80) | 10350051.40 (9099041.92-11468241.96) | 101.61 (99.02-104.22) | 65704498.76 (57969290.22-72989869.94) | 140867093.61 (124370457.12-156193663.52) | 114.39 (112.03-116.96) | 2294596.33 (1091955.68-4621567.45) | 4962550.30 (2367023.68-9974958.53) | 116.27 (113.85-118.84) |
| Middle | 5750921.82 (5058231.66-6410278.28) | 15437968.58 (13569373.27-17143283.15) | 168.44 (162.44-173.96) | 65080909.87 (57656740.57-72193595.36) | 192598780.54 (170454604.52-213634900.65) | 195.94 (190.22-201.67) | 2235431.10 (1076653.89-4486204.48) | 6708208.99 (3204061.77-13465746.07) | 200.09 (193.73-206.34) |
| Low-middle | 3035072.54 (2675262.62-3375779.45) | 7812257.58 (6892042.84-8676931.11) | 157.40 (153.90-160.50) | 33532956.94 (29739517.92-37224728.26) | 90721550.13 (80468537.50-100582853.83) | 170.54 (166.72-174.21) | 1135603.53 (547159.37-2278708.24) | 3110554.90 (1492430.03-6248885.29) | 173.91 (169.57-178.11) |
| Low | 1094622.36 (965733.07-1219371.20) | 2804976.17 (2479497.58-3122531.67) | 156.25 (153.06-159.98) | 11819920.82 (10482916.14-13238336.76) | 29900895.83 (26519031.31-33164828.91) | 152.97 (149.87-156.34) | 398377.03 (192603.83-798822.18) | 1018404.36 (489392.97-2037321.76) | 155.64 (152.16-159.48) |
| **Regions** | | | | | | | | | |

| | | | | | | | | | |
|---|---|---|---|---|---|---|---|---|---|
| Andean Latin America | 121136.54 (107214.18-134640.11) | 363396.50 (319747.57-402547.11) | 199.99 (193.11-206.57) | 1383605.90 (1228225.66-1528853.09) | 4431104.54 (3933345.88-4885803.62) | 220.26 (214.08-226.86) | 48493.67 (23257.34-97369.85) | 156693.11 (75061.50-316245.49) | 223.12 (216.04-230.24) |
| Australasia | 124185.65 (110781.01-137553.28) | 269451.73 (239374.24-301169.39) | 116.97 (110.86-123.94) | 1659800.14 (1490004.80-1834681.75) | 3922513.66 (3537650.57-4335715.14) | 136.32 (130.65-142.13) | 58806.52 (28504.14-118543.38) | 140906.06 (69471.52-286855.99) | 139.61 (132.45-146.50) |
| Caribbean | 139108.27 (123460.78-154387.50) | 296169.90 (262546.75-330668.29) | 112.91 (108.41-117.44) | 1698190.21 (1502560.56-1877188.61) | 3852891.90 (3412424.59-4252980.06) | 126.88 (123.48-130.72) | 59540.73 (28498.77-120118.39) | 135591.29 (64907.66-274656.30) | 127.73 (123.67-131.95) |
| Central Asia | 221868.54 (194411.32-249129.85) | 475854.12 (413121.22-535995.46) | 114.48 (110.03-119.11) | 2890239.45 (2514609.64-3288610.03) | 6010309.78 (5207826.24-6870321.27) | 107.95 (103.81-112.09) | 101371.35 (48801.93-204167.09) | 212364.52 (101660.30-425621.82) | 109.49 (104.77-114.13) |
| Central Europe | 699280.23 (618327.98-779465.69) | 960143.69 (849389.50-1070329.04) | 37.30 (34.72-40.16) | 9379902.77 (8274711.42-10510034.04) | 14519446.39 (12794343.18-16234190.97) | 54.79 (52.72-57.05) | 327000.82 (157196.16-659825.31) | 514447.14 (248116.66-1044283.53) | 57.32 (55.06-59.80) |
| Central Latin America | 509647.54 (450945.99-568025.64) | 1561604.80 (1379180.83-1731311.41) | 206.41 (200.75-211.99) | 5697761.23 (5042111.66-6304605.49) | 19197614.19 (16954459.18-21143360.14) | 236.93 (232.02-242.13) | 197933.26 (94851.15-399110.39) | 676745.24 (322648.75-1368409.61) | 241.91 (236.75-247.95) |
| Central Sub-Saharan Africa | 122007.42 (106783.23-135467.16) | 339701.23 (298430.94-378042.51) | 178.43 (171.70-185.29) | 1314104.88 (1159130.89-1464371.46) | 3517437.90 (3115894.36-3914394.31) | 167.67 (161.31-173.48) | 44741.36 (21341.87-90400.27) | 120682.34 (57012.18-241713.73) | 169.73 (162.08-177.16) |
| East Asia | 4836774.22 (4234495.29-5416116.68) | 12051700.47 (10562111.16-13556244.21) | 149.17 (141.74-155.84) | 55506569.16 (48473138.62-62109117.53) | 158285423.73 (139469182.88-176824968.19) | 185.17 (178.43-191.41) | 1904131.52 (915864.14-3832084.43) | 5518037.40 (2633493.69-11061496.89) | 189.79 (182.36-197.27) |
| Eastern Europe | 1495247.02 (1312342.06-1682318.80) | 1833037.93 (1612583.96-2058219.98) | 22.59 (20.55-24.59) | 21046221.16 (18447721.00-23746860.72) | 27107907.86 (23769327.16-30423620.18) | 28.80 (26.85-30.61) | 742362.50 (355455.53-1508176.60) | 965580.26 (465099.27-1951744.99) | 30.07 (28.04-32.23) |
| Eastern Sub-Saharan Africa | 366642.93 (322665.00-409381.62) | 1000971.00 (885083.79-1115451.19) | 173.01 (168.05-178.09) | 3912596.70 (3470701.35-4378271.80) | 10409572.22 (9265845.83-11557319.14) | 166.05 (161.11-171.11) | 132919.26 (63594.55-267611.27) | 358150.97 (171683.75-719704.24) | 169.45 (164.21-174.57) |
| High-income Asia Pacific | 1345776.30 (1189858.25-1491372.54) | 2189402.79 (1958142.42-2413152.29) | 62.69 (57.94-67.89) | 16572952.08 (14690573.82-18303601.17) | 34625938.50 (31148249.61-37978128.35) | 108.93 (104.50-114.01) | 597447.47 (286963.68-1207310.03) | 1276815.38 (611788.85-2579653.53) | 113.71 (108.72-119.51) |
| High-income North America | 1899644.64 (1695714.25-2093719.63) | 3457087.12 (3062826.32-3850765.87) | 81.99 (78.08-86.01) | 26834458.55 (24152313.72-29613959.41) | 51749678.99 (46318473.26-57318852.49) | 92.85 (91.14-94.56) | 964586.71 (463191.13-1948962.97) | 1857796.38 (900403.45-3761748.19) | 92.60 (90.60-94.99) |
| North Africa and Middle East | 839234.42 (738099.20-934460.23) | 2730761.38 (2402789.30-3044966.79) | 225.39 (220.33-230.34) | 9337845.50 (8286676.08-10395164.46) | 30491685.02 (27064615.42-33709288.17) | 226.54 (221.64-231.14) | 319579.97 (153014.32-643843.03) | 1049857.28 (503110.84-2115548.78) | 228.51 (223.18-234.36) |
| Oceania | 16658.74 (14676.47-18606.53) | 47742.61 (41835.00-53394.35) | 186.59 (178.78-194.27) | 176695.89 (156977.99-197592.99) | 508188.97 (451273.12-565164.81) | 187.61 (180.07-194.33) | 6038.08 (2899.44-12078.19) | 17457.20 (8450.50-34943.08) | 189.12 (181.49-197.41) |
| South Asia | 2993307.58 (2640026.16-3326475.62) | 8220377.67 (7241991.21-9115916.28) | 174.63 (169.92-179.16) | 32454744.62 (28765904.84-35895034.85) | 96531168.75 (85576493.91-106691000.59) | 197.43 (191.85-202.97) | 1094409.84 (528655.91-2197896.30) | 3311235.91 (1583933.88-6656918.84) | 202.56 (196.55-208.81) |
| Southeast Asia | 1128949.85 (993293.25-1259580.33) | 3261524.60 (2872404.58-3641133.22) | 188.90 (182.77-194.87) | 12662656.42 (11238788.45-14120227.79) | 39227934.85 (34567606.77-43609405.11) | 209.79 (203.92-215.72) | 432057.76 (206865.03-871587.99) | 1357627.76 (645150.32-2713214.37) | 214.22 (207.55-221.01) |
| Southern Latin America | 252629.79 (224152.65-281234.07) | 483946.18 (431133.20-536648.45) | 91.56 (87.42-95.94) | 3248110.10 (2899780.01-3599930.27) | 6538637.68 (5891362.39-7213429.02) | 101.31 (97.56-105.74) | 115112.19 (54895.53-232022.07) | 233579.59 (112226.69-468574.49) | 102.91 (98.14-108.12) |
| Southern Sub-Saharan Africa | 157577.09 (139016.14-175111.98) | 375227.74 (330227.08-417302.16) | 138.12 (135.35-140.84) | 1801904.29 (1590782.81-2000266.42) | 4289178.52 (3773086.32-4755346.42) | 138.04 (135.57-140.63) | 62840.69 (30076.93-125999.28) | 148991.49 (71570.91-298108.23) | 137.09 (133.91-140.69) |
| Tropical Latin America | 551639.12 (485844.29-614415.82) | 1568533.39 (1385103.16-1733205.81) | 184.34 (179.65-188.75) | 6184012.29 (5480737.52-6851449.28) | 19391654.78 (17180011.76-21545740.64) | 213.58 (209.54-217.77) | 213399.11 (102014.98-428853.21) | 678601.96 (325614.55-1368809.77) | 218.00 (213.08-223.10) |
| Western Europe | 2630868.23 (2349666.53-2928270.32) | 3918887.82 (3498381.84-4363204.13) | 48.96 (47.23-50.83) | 37369610.85 (33648956.22-41316152.02) | 59567388.69 (53848953.98-65960012.88) | 59.40 (57.90-60.97) | 1327348.42 (642542.56-2667248.36) | 2131317.24 (1038439.59-4287322.73) | 60.57 (58.74-62.55) |

| | | | | | | | | | |
|---|---|---|---|---|---|---|---|---|---|
| Western Sub-Saharan Africa | 448326.40 (395307.50-501301.02) | 1226621.72 (1078016.62-1368996.58) | 173.60 (169.58-177.46) | 4944717.74 (4378451.62-5533517.31) | 12813642.06 (11400708.56-14172789.53) | 159.14 (155.87-162.32) | 168735.44 (81206.00-339506.21) | 442087.29 (212438.64-888883.98) | 162.00 (158.37-165.66) |

**Table S2.** The ASIR, ASPR, ASDR, and EAPCs of osteoarthritis from 1990 to 2021

| Characteristics | ASIR (per 100,000 population) | | | ASPR (per 100,000 population) | | | ASDR (per 100,000 population) | | |
|---|---|---|---|---|---|---|---|---|---|
| | 1990 | 2021 | EAPC (95%CI) | 1990 | 2021 | EAPC (95%CI) | 1990 | 2021 | EAPC (95%CI) |
| | Both (95%UI) | Both (95%UI) | | Both (95%UI) | Both (95%UI) | | Both (95%UI) | Both (95%UI) | |
| **Global** | 489.78 (433.10-541.51) | 535.00 (472.38-591.97) | 0.33 (0.31-0.35) | 6393.12 (5683.20-7059.53) | 6967.29 (6180.70-7686.06) | 0.34 (0.31-0.37) | 222.80 (106.65-450.29) | 244.50 (117.06-493.11) | 0.37 (0.33-0.40) |
| **Sex** | | | | | | | | | |
| Male | 408.56 (361.23-452.94) | 445.74 (393.68-493.98) | 0.28 (0.25-0.30) | 5290.02 (4712.01-5894.75) | 5773.36 (5125.94-6420.96) | 0.29 (0.27-0.32) | 182.42 (87.58-369.04) | 200.52 (95.88-404.87) | 0.31 (0.29-0.34) |
| Female | 568.60 (502.58-627.86) | 621.32 (548.49-686.94) | 0.36 (0.32-0.39) | 7354.53 (6532.33-8109.16) | 8049.41 (7137.25-8892.72) | 0.38 (0.34-0.43) | 257.65 (123.39-520.40) | 284.14 (136.29-573.11) | 0.42 (0.37-0.47) |
| **Types** | | | | | | | | | |
| Osteoarthritis knee | 330.26 (284.34-375.75) | 353.67 (304.56-402.50) | 0.28 (0.26-0.30) | 3964.75 (3411.86-4536.40) | 4294.27 (3695.04-4910.76) | 0.33 (0.30-0.35) | 127.14 (62.17-246.99) | 137.59 (67.08-266.87) | 0.33 (0.30-0.36) |
| Osteoarthritis hand | 100.57 (74.51-128.13) | 119.09 (88.73-151.13) | 0.55 (0.48-0.61) | 1944.84 (1476.85-2478.43) | 2237.78 (1693.67-2851.21) | 0.52 (0.44-0.60) | 61.67 (27.93-127.04) | 70.94 (32.23-146.27) | 0.52 (0.45-0.60) |
| Osteoarthritis hip | 19.27 (14.72-24.41) | 20.67 (15.76-26.19) | 0.28 (0.26-0.30) | 391.66 (303.26-496.51) | 416.01(321.23-529.74) | 0.26 (0.23-0.28) | 12.43 (5.86-25.21) | 13.19 (6.20-26.65) | 0.26 (0.24-0.29) |
| Osteoarthritis other | 39.68 (32.01-50.71) | 41.57 (33.45-53.36) | 0.15 (0.15-0.16) | 678.37 (532.31-876.68) | 716.69 (560.15-930.47) | 0.18 (0.18-0.18) | 21.56 (10.01-45.31) | 22.78 (10.54-47.81) | 0.18 (0.18-0.18) |
| **SDI** | | | | | | | | | |
| High | 568.82 (505.90-627.74) | 611.30 (542.71-675.91) | 0.21 (0.17-0.25) | 7371.33 (6609.68-8130.17) | 7897.27 (7067.13-8689.88) | 0.24 (0.19-0.29) | 262.74 (126.00-529.15) | 283.13 (136.04-570.53) | 0.27 (0.21-0.33) |
| High-middle | 498.45 (441.11-549.27) | 548.07 (481.66-608.49) | 0.38 (0.35-0.42) | 6557.50 (5802.43-7265.76) | 7120.38 (6297.95-7879.76) | 0.37 (0.33-0.41) | 228.99 (109.20-462.13) | 250.58 (119.78-503.69) | 0.40 (0.36-0.44) |
| Middle | 475.42 (419.58-527.87) | 536.49 (473.16-595.02) | 0.48 (0.44-0.52) | 6066.13 (5375.33-6744.29) | 6903.80 (6123.00-7643.11) | 0.52 (0.48-0.56) | 208.27 (100.47-419.68) | 240.41 (115.09-483.99) | 0.57 (0.53-0.61) |
| Low-middle | 423.51 (374.68-469.50) | 480.13 (424.96-533.01) | 0.42 (0.40-0.44) | 5326.73 (4722.18-5924.73) | 6106.25 (5419.32-6763.20) | 0.45 (0.43-0.48) | 180.18 (86.93-364.19) | 209.35 (100.40-422.62) | 0.50 (0.48-0.53) |
| Low | 408.88 (362.19-452.55) | 447.12 (395.36-493.37) | 0.29 (0.28-0.31) | 5080.55 (4507.60-5674.54) | 5605.58 (4967.54-6230.60) | 0.32 (0.31-0.34) | 170.90 (82.56-345.11) | 190.93 (91.63-384.26) | 0.37 (0.35-0.39) |
| **Regions** | | | | | | | | | |
| Andean Latin America | 519.51 (461.53-577.69) | 578.18 (511.33-641.10) | 0.35 (0.34-0.37) | 6602.95 (5861.30-7291.11) | 7370.44 (6552.10-8123.13) | 0.35 (0.34-0.37) | 231.91 (111.18-466.16) | 260.94 (125.19-526.82) | 0.38 (0.36-0.40) |
| Australasia | 555.78 (493.55-617.22) | 620.09 (550.76-686.53) | 0.34 (0.31-0.36) | 7195.28 (6466.74-7950.86) | 7917.60 (7098.38-8735.71) | 0.31 (0.29-0.33) | 254.48 (123.19-513.81) | 283.38 (139.24-577.97) | 0.34 (0.31-0.36) |

| | | | | | | | | | |
|---|---|---|---|---|---|---|---|---|---|
| Caribbean | 513.03<br>(455.98-570.50) | 555.77<br>(493.24-617.51) | 0.29<br>(0.27-0.30) | 6514.84<br>(5762.16-7198.00) | 7134.56<br>(6327.26-7876.59) | 0.32<br>(0.31-0.33) | 228.48<br>(109.52-461.20) | 251.06<br>(120.09-508.25) | 0.34<br>(0.32-0.35) |
| Central Asia | 444.99<br>(392.55-498.51) | 504.46<br>(442.21-565.75) | 0.43<br>(0.39-0.46) | 6143.82<br>(5368.48-6965.53) | 7034.89<br>(6120.08-8010.15) | 0.47<br>(0.42-0.51) | 215.80<br>(104.18-434.30) | 249.29<br>(119.63-500.56) | 0.50<br>(0.45-0.55) |
| Central Europe | 474.27<br>(419.37-525.88) | 522.05<br>(460.81-580.35) | 0.33<br>(0.32-0.35) | 6276.97<br>(5554.86-7001.94) | 6948.51<br>(6129.15-7752.72) | 0.36<br>(0.34-0.37) | 218.64<br>(104.72-441.82) | 245.41<br>(117.63-496.21) | 0.41<br>(0.39-0.43) |
| Central Latin America | 527.96<br>(468.13-587.05) | 589.49<br>(521.45-652.52) | 0.37<br>(0.36-0.37) | 6661.13<br>(5900.48-7353.18) | 7499.49<br>(6635.38-8259.93) | 0.40<br>(0.39-0.41) | 231.66<br>(110.81-468.03) | 264.58<br>(126.15-535.65) | 0.45<br>(0.44-0.46) |
| Central Sub-Saharan Africa | 441.86<br>(390.35-491.24) | 463.08<br>(409.68-515.24) | 0.12<br>(0.07-0.16) | 5622.81<br>(4965.80-6260.93) | 5940.49<br>(5268.27-6589.54) | 0.13<br>(0.08-0.18) | 191.36<br>(91.09-387.34) | 204.30<br>(97.78-412.23) | 0.17<br>(0.11-0.23) |
| East Asia | 487.30<br>(428.32-544.04) | 554.47<br>(486.91-619.37) | 0.58<br>(0.50-0.65) | 6157.54<br>(5425.37-6866.85) | 7036.10<br>(6216.29-7835.76) | 0.60<br>(0.52-0.69) | 211.00<br>(102.08-424.58) | 245.04<br>(117.45-492.41) | 0.66<br>(0.58-0.75) |
| Eastern Europe | 550.43<br>(484.03-614.18) | 584.97<br>(515.25-651.42) | 0.29<br>(0.26-0.31) | 7541.08<br>(6611.08-8496.07) | 7906.11<br>(6954.04-8880.09) | 0.28<br>(0.24-0.32) | 265.83<br>(126.81-540.87) | 280.78<br>(134.37-567.03) | 0.32<br>(0.27-0.37) |
| Eastern Sub-Saharan Africa | 411.50<br>(363.20-457.47) | 461.02<br>(407.42-509.93) | 0.39<br>(0.37-0.40) | 5113.71<br>(4544.28-5704.44) | 5829.96<br>(5160.63-6476.62) | 0.44<br>(0.43-0.46) | 173.52<br>(83.30-351.43) | 200.97<br>(96.11-405.27) | 0.51<br>(0.49-0.52) |
| High-income Asia Pacific | 641.16<br>(568.18-707.78) | 682.07<br>(606.06-752.84) | 0.35<br>(0.22-0.49) | 8071.98<br>(7169.35-8905.87) | 8608.63<br>(7674.07-9485.19) | 0.42<br>(0.24-0.59) | 291.10<br>(139.90-587.96) | 314.98<br>(150.55-636.77) | 0.51<br>(0.29-0.73) |
| High-income North America | 605.72<br>(535.69-670.62) | 646.38<br>(572.29-715.37) | 0.07<br>(-0.07-0.21) | 7987.16<br>(7188.90-8824.97) | 8421.62<br>(7534.98-9282.03) | 0.09<br>(-0.04-0.23) | 286.25<br>(137.18-576.08) | 300.89<br>(144.87-606.97) | 0.07<br>(-0.08-0.22) |
| North Africa and Middle East | 424.31<br>(375.36-470.67) | 488.31<br>(433.70-542.33) | 0.44<br>(0.41-0.47) | 5362.22<br>(4751.55-5979.22) | 6265.22<br>(5572.94-6946.23) | 0.49<br>(0.45-0.52) | 183.41<br>(87.76-371.84) | 215.92<br>(103.37-437.62) | 0.51<br>(0.47-0.55) |
| Oceania | 441.66<br>(389.80-491.69) | 480.95<br>(423.12-536.36) | 0.25<br>(0.22-0.27) | 5637.24<br>(5022.89-6261.36) | 6196.48<br>(5474.55-6895.02) | 0.28<br>(0.25-0.30) | 192.33<br>(92.49-385.92) | 212.87<br>(102.99-428.52) | 0.30<br>(0.28-0.33) |
| South Asia | 430.77<br>(382.03-477.59) | 495.01<br>(436.64-548.03) | 0.47<br>(0.44-0.49) | 5407.04<br>(4798.72-5985.59) | 6326.13<br>(5612.39-7009.64) | 0.53<br>(0.51-0.56) | 181.92<br>(87.80-368.21) | 216.90<br>(104.00-438.04) | 0.60<br>(0.58-0.63) |
| Southeast Asia | 376.04<br>(332.26-418.37) | 437.13<br>(386.13-485.01) | 0.51<br>(0.50-0.51) | 4796.58<br>(4256.53-5377.23) | 5675.80<br>(5001.76-6320.89) | 0.56<br>(0.55-0.57) | 163.30<br>(78.14-329.58) | 196.18<br>(93.56-393.42) | 0.61<br>(0.61-0.62) |
| Southern Latin America | 540.18<br>(478.83-600.89) | 596.27<br>(530.39-660.42) | 0.29<br>0.26-0.32) | 7001.84<br>(6250.94-7759.15) | 7669.24<br>(6896.46-8466.30) | 0.29<br>(0.26-0.31) | 247.99<br>(118.32-499.99) | 273.53<br>(131.22-548.73) | 0.31<br>(0.28-0.34) |
| Southern Sub-Saharan Africa | 513.31<br>(454.83-568.81) | 557.24<br>(493.49-618.18) | 0.28<br>(0.27-0.29) | 6559.80<br>(5794.82-7289.01) | 7161.23<br>(6333.34-7951.33) | 0.31<br>(0.30-0.32) | 229.15<br>(110.10-460.99) | 249.45<br>(120.36-500.63) | 0.30<br>(0.29-0.32) |
| Tropical Latin America | 527.59<br>(467.44-585.25) | 589.12<br>(521.39-650.60) | 0.38<br>(0.37-0.39) | 6604.05<br>(5863.57-7307.66) | 7424.65<br>(6582.79-8241.00) | 0.40<br>(0.39-0.41) | 228.15<br>(109.07-460.14) | 259.93<br>(124.74-524.63) | 0.45<br>(0.43-0.46) |
| Western Europe | 521.51<br>(465.66-578.73) | 557.66<br>(497.27-618.53) | 0.19<br>(0.17-0.21) | 6736.67<br>(6071.81-7425.01) | 7113.44<br>(6407.11-7867.10) | 0.17<br>(0.15-0.19) | 238.56<br>(115.19-479.88) | 253.58<br>(123.06-510.55) | 0.19<br>(0.17-0.22) |
| Western Sub-Saharan Africa | 439.61<br>(387.27-489.07) | 483.84<br>(427.08-536.68) | 0.32<br>(0.30-0.33) | 5494.22<br>(4872.02-6124.65) | 6075.81<br>(5385.72-6757.27) | 0.34<br>(0.32-0.35) | 187.48<br>(90.32-378.77) | 210.09<br>(101.11-424.67) | 0.39<br>(0.37-0.40) |

**Table S3. The top three regions with the burden of osteoarthritis in 1990 and 2021**

| | Both | Number | Male | Number | Female | Number |
|---|---|---|---|---|---|---|
| Incident | East Asia | 12051700.47 | East Asia | 4826671.02 | East Asia | 7225029.45 |
| | South Asia | 8220377.67 | South Asia | 3222647.24 | South Asia | 4997730.43 |
| | Western Europe | 3918887.82 | Western Europe | 1564422.70 | Western Europe | 2354465.12 |
| Prevalent | East Asia | 158285423.70 | East Asia | 62255515.86 | East Asia | 96029907.87 |
| | South Asia | 96531168.75 | South Asia | 36747956.49 | South Asia | 59783212.25 |
| | Western Europe | 59567388.69 | Western Europe | 22700410.39 | Western Europe | 36866978.29 |
| DALYs | East Asia | 5518037.40 | East Asia | 2165589.00 | East Asia | 3352448.40 |
| | South Asia | 3311235.91 | South Asia | 1245901.34 | South Asia | 2065334.58 |
| | Western Europe | 2131317.24 | Western Europe | 797771.47 | Western Europe | 1333545.77 |
| ASIR | High-income Asia Pacific | 682.07 | High-income North America | 558.51 | High-income Asia Pacific | 829.80 |
| | High-income North America | 646.38 | Southern Sub-Saharan Africa | 536.31 | High-income North America | 729.65 |
| | Australasia | 620.09 | High-income Asia Pacific | 533.69 | Australasia | 708.49 |
| ASPR | High-income Asia Pacific | 8608.63 | High-income North America | 7344.41 | High-income Asia Pacific | 10345.22 |
| | High-income North America | 8421.62 | Eastern Europe | 6952.17 | High-income North America | 9384.59 |
| | Australasia | 7917.60 | Southern Sub-Saharan Africa | 6835.26 | Australasia | 8979.31 |
| ASDR | High-income Asia Pacific | 314.98 | High-income North America | 258.15 | High-income Asia Pacific | 384.97 |
| | High-income North America | 300.89 | Eastern Europe | 245.07 | High-income North America | 338.83 |
| | Australasia | 283.38 | Southern Sub-Saharan Africa | 238.88 | Australasia | 324.73 |

**Table S4. The incident case, prevalent case, DALYs and incresing changes of osteoarthritis among 204 countries and territories from 1990 to 2021**

| Country | Incident case | | | Prevalent case | | | DALYs | | |
|---|---|---|---|---|---|---|---|---|---|
| | 1990 | 2021 | 1990-2021 | 1990 | 2021 | 1990-2021 | 1990 | 2021 | 1990-2021 |
| | Both (95%UI) | Both (95%UI) | Change (%, 95%UI) | Both (95%UI) | Both (95%UI) | Change (%, 95%UI) | Both (95%UI) | Both (95%UI) | Change (%, 95%UI) |
| Afghanistan | 27287.14 (23791.40-30990.44) | 57027.01 (49382.26-64358.73) | 108.99 (94.53-123.73) | 321991.29 (283428.68-361803.75) | 546859.25 (480077.52-609142.83) | 69.84 (61.98-78.43) | 10777.63 (5227.44-21149.16) | 18084.93 (8774.79-36668.69) | 67.80 (59.04-77.24) |
| Albania | 9185.71 (8123.81-10211.06) | 17460.82 (15374.09-19596.73) | 90.09 (82.14-98.45) | 106048.15 (94170.97-117357.18) | 248148.77 (218659.47-276763.93) | 134.00 (125.92-142.37) | 3648.68 (1770.33-7290.51) | 8677.42 (4127.20-17605.22) | 137.82 (128.70-147.86) |
| Algeria | 58393.22 (51616.03-65377.18) | 209506.85 (183863.57-233742.72) | 258.79 (245.55-272.37) | 677464.87 (597902.38-758233.11) | 2432559.20 (2150380.55-2697828.39) | 259.07 (247.36-270.27) | 23230.05 (11038.45-47246.95) | 84149.99 (40665.34-169768.89) | 262.25 (248.84-276.36) |
| American Samoa | 159.64 (139.93-178.21) | 323.39 (283.81-361.15) | 102.57 (94.50-110.52) | 1736.72 (1537.19-1950.03) | 3972.39 (3516.21-4396.81) | 128.73 (121.12-135.92) | 60.82 (28.72-122.66) | 138.44 (66.50-282.11) | 127.65 (119.12-136.51) |
| Andorra | 308.07 (272.45-344.21) | 806.21 (714.75-900.19) | 161.69 (152.78-171.04) | 3839.88 (3446.04-4260.12) | 10783.66 (9708.90-11941.60) | 180.83 (173.70-188.03) | 136.40 (65.57-277.15) | 383.97 (184.47-765.25) | 181.50 (174.02-191.39) |
| Angola | 24000.87 (21032.81-26933.40) | 83357.00 (72517.33-93086.30) | 247.31 (235.91-258.64) | 252573.55 (218756.26-284310.81) | 869768.55 (761428.36-975501.47) | 244.36 (234.58-254.29) | 8723.32 (4124.01-17531.13) | 30158.42 (14452.69-60591.86) | 245.72 (232.86-257.99) |
| Antigua and Barbuda | 261.91 (234.54-289.52) | 663.09 (581.25-741.16) | 153.17 (144.41-163.06) | 3537.37 (3151.96-3899.99) | 8259.78 (7313.21-9225.89) | 133.50 (127.24-140.72) | 125.18 (59.09-252.46) | 290.95 (138.89-585.96) | 132.42 (124.55-141.24) |
| Argentina | 175240.51 (154713.47-195453.94) | 311889.59 (277214.89-345791.60) | 77.98 (72.67-83.68) | 2279648.28 (2035021.59-2531459.93) | 4194310.64 (3775256.87-4616497.10) | 83.99 (79.04-89.07) | 80960.14 (38452.28-162324.99) | 150106.55 (71901.74-303301.74) | 85.41 (79.39-91.32) |
| Armenia | 12389.73 (10850.76-13989.92) | 19889.61 (17445.58-22467.64) | 60.53 (54.59-66.86) | 156998.66 (136161.01-177816.25) | 296565.56 (256205.25-339560.73) | 88.90 (82.10-96.17) | 5466.94 (2629.43-10899.93) | 10518.06 (5107.34-21121.97) | 92.39 (84.34-100.34) |
| Australia | 103534.97 (92498.77-114683.22) | 225080.14 (199821.48-251381.91) | 117.40 (110.30-124.99) | 1383918.40 (1243299.24-1529471.11) | 3294936.82 (2968752.85-3631540.28) | 138.09 (131.22-145.15) | 49047.85 (23740.21-98909.20) | 118305.78 (58625.77-241248.79) | 141.20 (132.64-149.53) |
| Austria | 51884.78 (46358.05-57454.54) | 78648.62 (70337.56-87916.28) | 51.58 (47.46-56.38) | 750066.72 (675574.61-827423.14) | 1167787.79 (1058282.84-1289388.54) | 55.69 (51.91-59.80) | 26566.64 (12792.12-53033.56) | 41716.45 (20002.70-84033.57) | 57.03 (52.39-62.41) |
| Azerbaijan | 25059.25 (21622.61-28310.85) | 61652.84 (53118.12-70036.64) | 146.03 (137.64-155.09) | 323778.81 (279068.31-372561.75) | 771551.15 (660881.70-879386.85) | 138.30 (129.72-146.95) | 11420.29 (5453.69-23077.56) | 27347.57 (13226.90-54528.69) | 139.46 (129.47-148.79) |
| Bahamas | 993.23 (876.28-1107.19) | 2722.32 (2370.90-3044.24) | 174.09 (164.05-186.66) | 11361.45 (10136.65-12639.17) | 32754.01 (28800.53-36382.87) | 188.29 (180.03-197.38) | 402.66 (190.94-809.57) | 1159.35 (558.16-2332.32) | 187.92 (178.43-198.24) |
| Bahrain | 1351.88 (1194.94-1500.38) | 8147.28 (7061.02-9159.45) | 502.66 (474.56-531.18) | 12686.25 (11231.54-14184.64) | 80203.38 (70945.51-90083.32) | 532.21 (511.00-553.10) | 438.81 (205.89-900.47) | 2765.90 (1306.16-5571.85) | 530.32 (507.75-557.83) |
| Bangladesh | 223684.26 (198212.01-248765.26) | 690026.90 (612219.56-767880.58) | 208.48 (198.21-219.50) | 2470610.86 (2192025.94-2737062.55) | 8297327.05 (7370625.42-9214630.37) | 235.84 (225.54-245.94) | 83618.70 (40288.47-168313.21) | 284515.64 (137093.17-577735.07) | 240.25 (227.58-254.29) |
| Barbados | 1359.66 (1202.53-1509.04) | 2709.65 (2387.50-3047.14) | 99.29 (90.32-107.32) | 19303.96 (17252.74-21345.22) | 38371.00 (33784.92-42487.32) | 98.77 (93.12-104.20) | 687.86 (321.83-1389.17) | 1365.63 (649.64-2778.74) | 98.53 (91.75-104.64) |
| Belarus | 63143.24 (55568.29-70531.80) | 80484.3 1(70517.89-90198.29) | 27.46 (23.91-31.25) | 894797.22 (788274.07-1008658.33) | 1219042.64 (1073903.34-1368927.41) | 36.24 (32.60-39.95) | 31504.48 (15062.61-64042.55) | 43468.46 (20750.22-86959.82) | 37.98 (33.28-42.60) |
| Belgium | 68465.60 (60991.54-76513.42) | 97578.64 (86702.98-108987.00) | 42.52 (38.44-47.40) | 982108.45 (881171.33-1084366.44) | 1478912.90 (1337143.06-1635290.53) | 50.59 (47.12-54.72) | 34843.91 (16682.98-69416.58) | 52625.47 (25527.62-106465.48) | 51.03 (46.49-55.67) |
| Belize | 496.98 (440.28-550.02) | 2061.49 (1815.72-2294.14) | 314.81 (300.32-331.15) | 5979.89 (5313.62-6634.81) | 23216.26 (20541.14-25750.71) | 288.24 (276.60-302.31) | 209.83 (101.45-419.47) | 816.83 (390.96-1663.82) | 289.29 (275.97-305.81) |
| Benin | 9508.32 (8400.25-10545.90) | 32568.98 (28506.51-36258.31) | 242.53 (230.12-253.58) | 106754.11 (94726.97-119330.90) | 343780.31 (306791.56-381589.96) | 222.03 (211.40-233.09) | 3618.89 (1742.58-7290.42) | 11869.55 (5697.97-23986.80) | 227.99 (214.86-241.00) |

| | | | | | | | | | |
|---|---|---|---|---|---|---|---|---|---|
| Bermuda | 384.12 (338.53-426.61) | 670.47 (592.72-745.56) | 74.55 (67.76-83.02) | 4755.37 (4236.07-5249.93) | 9996.99 (8837.57-10989.95) | 110.23 (104.37-117.53) | 170.12 (80.51-345.29) | 359.06 (172.79-735.43) | 111.06 (104.01-119.13) |
| Bhutan | 1259.13 (1111.55-1408.84) | 3180.50 (2819.72-3532.29) | 152.60 (144.30-161.59) | 13318.69 (11793.63-14892.34) | 37238.23 (33076.40-41199.34) | 179.59 (170.35-189.61) | 452.93 (219.90-908.85) | 1281.59 (621.12-2600.86) | 182.96 (171.36-194.61) |
| Bolivia (Plurinational State of) | 18351.33 (16218.11-20443.03) | 57029.84 (50392.13-63358.58) | 210.77 (200.87-220.54) | 203683.60 (180657.93-226607.72) | 666485.83 (590520.74-738699.58) | 227.22 (216.97-237.31) | 7050.14 (3371.39-14207.89) | 23288.59 (11170.09-47119.68) | 230.33 (218.43-242.62) |
| Bosnia and Herzegovina | 19585.38 (17166.30-21960.72) | 26326.54 (23132.59-29466.07) | 34.42 (28.99-39.92) | 234845.20 (206726.13-262201.70) | 391736.09 (346442.37-440350.45) | 66.81 (60.57-73.22) | 8100.04 (3852.38-16233.06) | 13707.03 (6542.58-27707.40) | 69.22 (61.94-76.64) |
| Botswana | 3011.99 (2675.04-3378.67) | 10298.12 (9102.60-11420.24) | 241.90 (229.19-254.36) | 33411.74 (29776.22-37407.51) | 109840.34 (97516.75-122269.24) | 228.75 (218.62-238.85) | 1156.19 (547.72-2324.35) | 3800.02 (1831.39-7654.76) | 228.67 (217.89-241.46) |
| Brazil | 538758.49 (474540.52-599987.99) | 1533312.78 (1353748.32-1694669.80) | 184.60 (179.87-189.11) | 6033181.80 (5347514.03-6684708.36) | 18970857.79 (16804901.87-21077260.10) | 214.44 (210.36-218.66) | 208123.58 (99503.48-418167.95) | 663901.66 (318773.54-1339004.65) | 218.99 (214.05-224.24) |
| Brunei Darussalam | 906.73 (797.36-1003.88) | 3203.59 (2817.53-3554.52) | 253.31 (239.69-266.33) | 9011.38 (8058.41-9964.31) | 34031.74 (30247.67-37597.12) | 277.65 (266.87-288.94) | 321.78 (153.33-649.16) | 1223.46 (582.86-2448.41) | 280.22 (267.22-293.12) |
| Bulgaria | 57146.41 (50341.94-64002.68) | 59409.74 (52593.76-65848.04) | 3.96 (0.39-7.53) | 791590.39 (692118.02-889939.47) | 936043.36 (826382.05-1044846.14) | 18.25 (14.88-21.59) | 27784.25 (13266.96-55751.93) | 33206.64 (16137.48-67206.41) | 19.52 (15.87-23.85) |
| Burkina Faso | 19593.17 (17143.28-21970.66) | 49304.03 (43078.96-54825.90) | 151.64 (142.32-161.04) | 217159.12 (192068.00-243494.94) | 521316.59 (463357.74-586311.20) | 140.06 (132.63-148.10) | 7331.69 (3537.23-14818.35) | 17798.28 (8611.92-35905.27) | 142.76 (133.44-152.05) |
| Burundi | 10533.50 (9293.49-11673.84) | 26111.53 (22939.76-29320.36) | 147.89 (140.45-155.14) | 116072.34 (103027.25-129637.18) | 264422.54 (233713.33-296285.47) | 127.81 (121.50-134.85) | 3955.43 (1913.25-7939.87) | 9035.49 (4359.22-18146.02) | 128.43 (120.79-137.76) |
| Cabo Verde | 877.51 (774.37-988.10) | 2532.98 (2238.00-2844.88) | 188.66 (175.76-200.88) | 11780.94 (10397.80-13212.12) | 29550.82 (26139.30-32888.11) | 150.84 (141.20-160.24) | 405.14 (195.79-813.01) | 1027.46 (494.19-2071.01) | 153.61 (142.44-165.08) |
| Cambodia | 18793.04 (16448.32-21070.10) | 56902.76 (50230.93-63537.98) | 202.79 (191.84-216.01) | 204929.54 (181011.33-228887.53) | 659749.49 (587709.16-739198.60) | 221.94 (209.71-234.60) | 6904.52 (3331.86-14077.32) | 22484.74 (10740.19-45064.47) | 225.65 (211.08-241.01) |
| Cameroon | 23980.27 (21000.70-26764.92) | 82772.35 (73055.74-92139.82) | 245.17 (231.78-257.42) | 259289.49 (230455.47-290263.92) | 852478.86 (762205.78-940130.55) | 228.77 (217.69-239.78) | 8831.21 (4271.94-17831.07) | 29381.20 (14159.32-59080.32) | 232.70 (219.97-246.46) |
| Canada | 128827.85 (114244.35-144137.73) | 263975.92 (232772.41-297164.84) | 104.91 (97.85-111.89) | 1824017.73 (1600678.65-2046086.79) | 4163156.38 (3679838.46-4663100.96) | 128.24 (122.10-133.97) | 63957.74 (31071.65-130702.97) | 146844.93 (71051.19-296531.55) | 129.60 (121.61-137.44) |
| Central African Republic | 5883.78 (5145.16-6580.74) | 13041.90 (11395.03-14582.93) | 121.66 (113.19-129.27) | 61668.37 (54203.05-68965.73) | 129289.96 (114759.95-144249.08) | 109.65 (102.31-116.53) | 2079.57 (1000.01-4212.52) | 4377.99 (2111.72-8874.33) | 110.52 (101.96-119.62) |
| Chad | 12181.15 (10745.41-13630.81) | 30986.72 (27232.40-34601.58) | 154.38 (145.00-165.12) | 139179.70 (123429.64-155558.52) | 318880.39 (282303.88-358853.94) | 129.11 (120.68-138.49) | 4708.41 (2276.20-9519.13) | 10825.32 (5277.49-21890.88) | 129.91 (119.83-140.17) |
| Chile | 58213.53 (51807.45-64572.81) | 145187.62 (129824.00-162035.35) | 149.41 (140.75-157.90) | 701689.44 (626386.77-776446.26) | 1951585.56 (1759442.64-2155187.41) | 178.13 (169.93-186.00) | 24661.87 (11880.33-50150.57) | 69420.82 (33532.08-138325.97) | 181.49 (171.91-191.12) |
| China | 4654141.48 (4075192.05-5212885.65) | 11652721.19 (10207638.21-13107929.18) | 150.37 (142.75-157.10) | 53352514.68 (46603087.10-59685781.45) | 152848105.93 (134655962.16-170842262.84) | 186.49 (179.62-192.93) | 1829415.63 (880107.77-3682519.91) | 5327389.98 (2541781.20-10678674.95) | 191.21 (183.62-198.69) |
| Colombia | 104721.43 (93002.31-117050.64) | 311332.96 (273697.43-346614.46) | 197.30 (186.34-208.86) | 1169852.16 (1033377.13-1299615.46) | 4025156.32 (3565880.18-4466444.72) | 244.07 (233.32-255.40) | 40679.96 (19449.83-81303.31) | 142072.89 (67444.44-289099.67) | 249.25 (235.03-262.05) |
| Comoros | 956.02 (837.75-1072.32) | 2635.83 (2323.09-2942.15) | 175.71 (166.33-184.21) | 10342.70 (9169.82-11583.52) | 29374.78 (25981.92-32840.57) | 184.01 (174.26-193.01) | 353.04 (170.19-715.41) | 1012.17 (485.55-2055.19) | 186.70 (175.23-197.46) |
| Congo | 5796.06 (5101.27-6478.71) | 19068.59 (16720.31-21208.70) | 228.99 (216.45-241.66) | 65397.22 (57773.19-72900.54) | 196134.98 (174147.16-218303.13) | 199.91 (189.93-209.39) | 2254.86 (1070.60-4562.74) | 6791.77 (3215.27-13707.01) | 201.21 (189.73-213.07) |
| Cook Islands | 74.93 (66.09-83.69) | 140.94 (124.11-157.69) | 88.10 (81.13-95.99) | 892.11 (791.93-990.89) | 2010.43 (1774.72-2225.18) | 125.36 (117.58-132.99) | 31.17 (14.92-61.02) | 70.64 (34.04-142.35) | 126.60 (118.31-135.46) |
| Costa Rica | 9940.82 (8816.29-11032.31) | 31516.89 (27880.71-35173.45) | 217.05 (205.51-227.57) | 115531.98 (102589.18-127712.24) | 404928.35 (358473.04-447915.59) | 250.49 (240.31-260.42) | 4031.73 (1924.49-8121.14) | 14231.75 (6859.95-28750.09) | 252.99 (241.13-264.18) |
| Croatia | 29753.09 (26249.29-33364.16) | 35691.03 (31552.11-39616.34) | 19.96 (15.66-24.69) | 395928.51 (347321.39-445002.64) | 565409.43 (499041.10-630962.62) | 42.81 (38.67-47.05) | 13849.69 (6614.20-28014.50) | 19990.65 (9654.29-40472.21) | 44.34 (39.42-49.51) |

| | | | | | | | | | |
|---|---|---|---|---|---|---|---|---|---|
| Cuba | 52466.00 (46435.46-58324.29) | 99435.27 (87342.40-111905.97) | 89.52 (82.27-97.67) | 659381.32 (583439.64-733482.26) | 1372809.99 (1213433.06-1525482.71) | 108.20 (102.48-114.38) | 23141.68 (11051.41-46779.99) | 48463.88 (23028.96-97796.59) | 109.42 (102.02-116.29) |
| Cyprus | 4013.93 (3569.11-4449.11) | 10305.97 (9253.59-11402.55) | 156.76 (148.78-165.36) | 51643.29 (46463.25-57278.09) | 139029.87 (124996.46-153247.27) | 169.21 (162.33-176.49) | 1817.83 (872.79-3643.38) | 4955.66 (2402.53-9910.35) | 172.61 (164.46-182.03) |
| Czechia | 64113.83 (56721.67-72019.40) | 91249.26 (81003.96-101781.74) | 42.32 (37.75-46.55) | 907575.23 (800750.92-1018733.77) | 1420152.14 (1247133.11-1591851.31) | 56.48 (52.49-60.65) | 31730.75 (15387.70-64336.89) | 50206.82 (24413.49-101810.86) | 58.23 (53.42-63.48) |
| Côte d'Ivoire | 23224.20 (20351.59-25968.09) | 73190.20 (63803.77-81621.09) | 215.15 (204.97-225.04) | 230694.22 (205327.51-256963.09) | 733425.47 (653903.50-817875.59) | 217.92 (208.12-227.62) | 7780.95 (3776.21-15610.06) | 25227.82 (12158.00-51055.60) | 224.23 (211.30-236.40) |
| Democratic People's Republic of Korea | 90678.62 (79002.70-103016.81) | 175530.47 (153632.18-197148.03) | 93.57 (84.19-101.89) | 1049463.99 (928192.12-1174686.51) | 2222257.29 (1964539.46-2482777.60) | 111.75 (101.53-120.33) | 36130.55 (17172.37-72746.09) | 77009.62 (37281.74-155704.45) | 113.14 (102.47-123.93) |
| Democratic Republic of the Congo | 82694.31 (72457.72-92054.28) | 213656.55 (186933.96-237905.56) | 158.37 (149.69-167.12) | 891752.44 (786537.97-994780.98) | 2209192.10 (1952823.80-2450224.17) | 147.74 (139.83-154.89) | 30223.05 (14461.50-61245.11) | 75436.98 (35464.48-151454.24) | 149.60 (139.89-158.64) |
| Denmark | 36836.64 (32957.21-40594.90) | 48847.84 (43441.28-54516.68) | 32.61 (27.23-38.73) | 543684.57 (485653.43-603097.84) | 744525.08 (669360.85-823259.52) | 36.94 (30.76-43.72) | 19537.51 (9558.00-39975.81) | 26730.06 (12922.90-54646.46) | 36.81 (29.78-44.67) |
| Djibouti | 781.05 (686.81-876.05) | 4367.79 (3830.24-4867.23) | 459.22 (438.81-482.03) | 7545.72 (6712.94-8440.38) | 43542.62 (38604.56-48600.45) | 477.05 (456.94-497.66) | 258.78 (124.05-522.18) | 1513.32 (720.02-3030.18) | 484.79 (460.33-511.48) |
| Dominica | 285.45 (254.40-316.27) | 469.16 (413.65-522.71) | 64.36 (58.51-70.14) | 3753.89 (3340.68-4163.48) | 6092.77 (5387.90-6793.40) | 62.31 (57.67-67.28) | 131.68 (63.23-266.59) | 214.28 (102.56-429.25) | 62.73 (57.52-68.51) |
| Dominican Republic | 21754.22 (19211.65-24251.40) | 60247.27 (53094.17-67341.16) | 176.95 (168.48-186.37) | 247859.76 (219596.97-275061.42) | 737841.02 (654420.77-816064.74) | 197.68 (189.64-206.65) | 8702.35 (4128.44-17379.75) | 25997.26 (12460.17-52425.54) | 198.74 (188.69-209.14) |
| Ecuador | 32268.04 (28495.96-35886.48) | 100807.30 (88994.85-111969.16) | 212.41 (201.60-222.80) | 366819.69 (326443.66-403917.82) | 1246863.63 (1106360.22-1378365.25) | 239.91 (229.76-250.01) | 12889.78 (6151.41-25626.59) | 44084.07 (21040.07-87905.46) | 242.01 (230.48-253.43) |
| Egypt | 145297.21 (127232.23-162962.00) | 395500.13 (346708.88-441377.26) | 172.20 (161.87-181.40) | 1545496.84 (1357026.50-1727343.32) | 4288855.64 (3812604.45-4764960.99) | 177.51 (167.13-186.32) | 52976.93 (25225.48-107326.45) | 147792.63 (71251.85-303928.39) | 178.98 (167.94-190.48) |
| El Salvador | 16197.47 (14204.34-17967.28) | 34373.97 (30447.28-38032.10) | 112.22 (105.49-119.52) | 191965.61 (169692.56-213064.51) | 443375.15 (395271.73-487949.71) | 130.97 (124.47-138.44) | 6668.21 (3151.57-13493.65) | 15547.76 (7438.88-31269.02) | 133.16 (125.69-142.66) |
| Equatorial Guinea | 929.05 (812.62-1043.91) | 3776.05 (3310.50-4195.17) | 306.44 (286.46-327.64) | 10007.26 (8846.77-11210.93) | 37815.17 (33405.54-42295.39) | 277.88 (261.45-295.77) | 336.01 (163.76-681.50) | 1308.13 (622.68-2612.33) | 289.31 (270.26-310.99) |
| Eritrea | 6019.71 (5274.97-6752.18) | 16112.85 (14178.94-18023.32) | 167.67 (157.09-178.34) | 57678.48 (51105.32-64536.90) | 159725.62 (141263.01-178274.41) | 176.92 (166.72-186.93) | 1954.85 (934.37-3916.13) | 5430.56 (2571.64-10768.52) | 177.80 (163.62-190.58) |
| Estonia | 10258.10 (9077.34-11433.97) | 11714.70 (10394.73-13129.72) | 14.20 (10.61-18.23) | 147423.89 (129808.22-165855.44) | 194934.07 (171215.15-218687.17) | 32.23 (28.58-35.84) | 5205.46 (2497.89-10404.20) | 7009.67 (3367.59-14039.51) | 34.66 (30.00-39.40) |
| Eswatini | 1688.03 (1487.87-1891.38) | 3821.29 (3366.88-4244.42) | 126.38 (117.44-134.72) | 17749.15 (15782.33-19682.37) | 40512.49 (36024.77-45098.67) | 128.25 (120.20-136.45) | 613.89 (291.83-1241.89) | 1392.87 (670.26-2793.90) | 126.89 (117.70-137.22) |
| Ethiopia | 99638.17 (87384.88-111461.84) | 270741.86 (237202.63-300718.03) | 171.73 (161.96-180.80) | 1040402.55 (920397.03-1164693.65) | 2881339.99 (2528598.05-3201411.61) | 176.94 (166.27-187.31) | 35251.73 (16935.10-71267.94) | 100011.78 (47612.03-199996.70) | 183.71 (171.57-195.04) |
| Fiji | 2368.63 (2091.44-2655.62) | 5074.51 (4459.45-5702.04) | 114.24 (105.50-123.02) | 24590.71 (21890.82-27545.86) | 60348.78 (53323.43-67176.20) | 145.41 (136.77-153.43) | 848.38 (405.51-1689.36) | 2098.46 (1015.88-4237.81) | 147.35 (136.37-157.06) |
| Finland | 33541.08 (29882.46-37178.18) | 49029.86 (43897.84-54528.36) | 46.18 (40.76-51.40) | 461234.50 417021.55-508410.45) | 782910.16 (706234.31-867239.94) | 69.74 (65.07-74.34) | 16276.74 (7927.66-32884.35) | 27939.39 (13503.13-56189.50) | 71.65 (66.55-77.51) |
| France | 365820.91 (325843.18-407123.37) | 558962.11 (496888.97-624475.41) | 52.80 (48.50-58.04) | 5232168.25 (4717987.83-5780825.67) | 8701539.81 (7878884.73-9534116.68) | 66.31 (62.20-70.60) | 185036.14 (89539.89-372835.49) | 310427.51 (150026.94-625920.60) | 67.77 (62.75-73.27) |
| Gabon | 2703.35 (2373.30-3026.86) | 6801.14 (5969.26-7587.90) | 151.58 (141.83-160.92) | 32706.04 (28948.16-36480.07) | 75237.14 (67262.57-83892.72) | 130.04 (122.63-138.33) | 1124.54 (539.36-2295.03) | 2609.06 (1240.78-5238.45) | 132.01 (122.66-141.50) |
| Gambia | 1848.90 (1624.41-2066.21) | 6002.80 (5266.59-6646.15) | 224.67 (212.99-235.79) | 19411.31 (17255.44-21783.29) | 64113.82 (56832.18-70970.49) | 230.29 (218.29-241.85) | 664.08 (321.13-1334.01) | 2214.59 (1060.95-4490.50) | 233.48 (218.48-247.20) |
| Georgia | 27867.53 (24387.12-31363.36) | 24890.41 (21859.79-27991.84) | -10.68 (-13.67--7.68) | 383419.92 (332432.40-434597.41) | 383788.96 (333862.79-434176.64) | 0.10 (-2.97-3.21) | 13507.47 (6569.29-27037.37) | 13535.84 (6550.61-27155.19) | 0.21 (-3.54-3.89) |

| | | | | | | | | | |
|---|---|---|---|---|---|---|---|---|---|
| Germany | 579625.02<br>(515357.90-644918.67) | 787712.39<br>(698574.45-873750.11) | 35.90<br>(31.64-40.65) | 8283522.19<br>(7453152.16-9115020.16) | 12268606.80<br>(11049241.94-13609527.68) | 48.11<br>(44.11-51.56) | 294049.23<br>(142784.64-593358.30) | 437790.86<br>(215922.65-879846.36) | 48.88<br>(43.84-53.50) |
| Ghana | 37544.76<br>(32916.40-41932.30) | 110975.97<br>(96894.81-123575.19) | 195.58<br>(186.02-206.87) | 411219.36<br>(361888.03-458705.81) | 1183031.13<br>(1050661.29-1309266.24) | 187.69<br>(178.44-198.43) | 14299.58<br>(6761.04-28809.14) | 41273.33<br>(19680.79-82749.66) | 188.63<br>(176.95-201.79) |
| Greece | 68731.58<br>(61358.99-77471.38) | 92595.48<br>(82685.76-102213.93) | 34.72<br>(29.08-39.47) | 936119.70<br>(843678.89-1038256.66) | 1436278.45<br>(1297936.74-1588189.63) | 53.43<br>(48.44-57.74) | 32944.31<br>(15987.31-66126.53) | 51134.34<br>(24852.32-103742.30) | 55.21<br>(49.96-60.33) |
| Greenland | 191.57<br>(167.37-215.74) | 355.46<br>(311.36-401.72) | 85.55<br>(75.45-95.94) | 2090.33<br>(1831.10-2337.98) | 4610.22<br>(4034.08-5172.24) | 120.55<br>(112.65-127.83) | 72.35<br>(34.63-148.00) | 162.20<br>(78.22-326.78) | 124.17<br>(114.31-133.49) |
| Grenada | 313.17<br>(279.61-346.44) | 682.40<br>(601.65-764.37) | 117.90<br>(108.45-128.23) | 4378.35<br>(3881.50-4886.00) | 8483.62<br>(7510.71-9412.90) | 93.76<br>(87.10-100.32) | 153.47<br>(73.79-309.41) | 297.28<br>(141.80-600.57) | 93.71<br>(86.28-101.15) |
| Guam | 525.08<br>(460.61-585.83) | 1156.80<br>(1019.10-1289.41) | 120.31<br>(109.74-131.00) | 5724.05<br>(5078.31-6418.67) | 16202.16<br>(14300.39-17994.77) | 183.05<br>(172.08-193.56) | 201.86<br>(96.92-404.67) | 576.88<br>(277.95-1150.01) | 185.78<br>(174.33-199.59) |
| Guatemala | 19813.99<br>(17425.97-21949.64) | 63651.95<br>(56005.20-70927.60) | 221.25<br>(211.66-233.36) | 214789.96<br>(190429.18-238337.88) | 748843.34<br>(663036.55-831469.32) | 248.64<br>(238.90-260.55) | 7374.50<br>(3516.16-14877.88) | 25805.62<br>(12422.04-52785.37) | 249.93<br>(237.49-263.44) |
| Guinea | 14609.02<br>(12924.97-16365.56) | 29813.74<br>(26205.26-33039.84) | 104.08<br>(96.59-111.78) | 167938.02<br>(148986.97-187462.09) | 320099.56<br>(284903.23-358791.66) | 90.61<br>(83.78-97.15) | 5700.16<br>(2768.42-11612.48) | 10941.10<br>(5275.45-21930.95) | 91.94<br>(84.29-99.90) |
| Guinea-Bissau | 1958.89<br>(1718.92-2193.52) | 4439.20<br>(3882.55-4936.63) | 126.62<br>(118.03-134.84) | 20806.93<br>(18382.44-23276.36) | 43794.78<br>(38803.14-48753.24) | 110.48<br>(103.67-117.17) | 705.20<br>(344.95-1425.89) | 1497.58<br>(715.81-2994.93) | 112.36<br>(103.52-120.73) |
| Guyana | 2239.53<br>(1980.51-2486.45) | 4056.52<br>(3553.92-4547.71) | 81.13<br>(73.87-87.79) | 24709.67<br>(21920.59-27494.33) | 48016.70<br>(42379.57-53318.82) | 94.32<br>(87.26-99.98) | 848.84<br>(406.54-1720.27) | 1659.59<br>(797.39-3373.31) | 95.51<br>(87.75-102.05) |
| Haiti | 16740.09<br>(14742.48-18656.33) | 44247.15<br>(39083.19-49488.76) | 164.32<br>(154.71-174.24) | 180842.66<br>(161578.40-203285.33) | 461322.52<br>(408504.21-515425.72) | 155.10<br>(145.88-163.86) | 6139.73<br>(2993.49-12364.46) | 15722.29<br>(7588.42-31426.85) | 156.07<br>(145.72-167.55) |
| Honduras | 11445.56<br>(10046.37-12716.05) | 39418.14<br>(34602.20-43968.59) | 244.40<br>(233.72-256.17) | 127604.09<br>(112175.55-142053.65) | 446901.99<br>(397249.99-497171.51) | 250.23<br>(241.27-260.69) | 4415.74<br>(2127.23-8787.25) | 15482.73<br>(7462.67-31056.56) | 250.63<br>(239.87-263.66) |
| Hungary | 68521.47<br>(60690.56-76505.92) | 84064.81<br>(74011.72-93556.28) | 22.68<br>(18.80-26.43) | 970946.61<br>(856018.03-1089127.31) | 1310359.18<br>(1158647.09-1472743.41) | 34.96<br>(31.46-38.40) | 33900.37<br>(16393.85-68585.64) | 46542.49<br>(22389.95-93905.81) | 37.29<br>(33.00-41.41) |
| Iceland | 1419.37<br>(1269.63-1569.62) | 2753.77<br>(2458.83-3049.80) | 94.01<br>(86.39-101.47) | 19611.01<br>(17550.22-21782.15) | 40058.36<br>(36184.11-44272.24) | 104.26<br>(96.16-111.85) | 706.58<br>(345.79-1451.70) | 1447.42<br>(705.50-2897.03) | 104.85<br>(94.86-114.71) |
| India | 2474842.12<br>(2182570.85-2747485.14) | 6701764.08<br>(5911471.07-7428209.67) | 170.80<br>(165.73-175.74) | 26639507.16<br>(23639835.22-29480348.87) | 79214251.02<br>(70296947.65-87586539.76) | 197.36<br>(191.83-203.39) | 897470.86<br>(433854.40-1802635.58) | 2718457.05<br>(1302654.53-5462599.64) | 202.90<br>(196.48-209.82) |
| Indonesia | 457055.30<br>(400852.38-509725.69) | 1308805.81<br>(1146853.19-1469964.88) | 186.36<br>(180.53-192.70) | 4983948.48<br>(4411432.41-5586203.05) | 14891306.50<br>(13109591.73-16672857.83) | 198.79<br>(192.65-205.58) | 170423.96<br>(81686.92-343678.02) | 516616.26<br>(245937.64-1031740.82) | 203.14<br>(195.93-210.90) |
| Iran<br>(Islamic Republic of) | 132275.54<br>(116401.28-147760.95) | 449414.40<br>(395815.84-499366.80) | 239.76<br>(233.35-246.65) | 1469041.32<br>(1295481.49-1641745.77) | 5083977.42<br>(4515256.82-5645682.27) | 246.07<br>(240.74-251.73) | 50356.09<br>(24041.12-101206.43) | 175263.35<br>(83503.05-351986.30) | 248.05<br>(242.26-254.61) |
| Iraq | 40791.43<br>(35892.06-45167.52) | 149558.21<br>(130580.64-167726.17) | 266.64<br>(254.59-280.02) | 462766.89<br>(411019.96-512938.21) | 1615817.36<br>(1436788.21-1801750.95) | 249.16<br>(239.06-259.93) | 15832.91<br>(7529.69-31775.40) | 54964.14<br>(26620.05-111028.98) | 247.15<br>(234.15-259.65) |
| Ireland | 18952.62<br>(16953.86-20966.30) | 38498.71<br>(34402.66-42749.67) | 103.13<br>(97.32-109.86) | 259982.82<br>(232118.82-287443.39) | 532942.69<br>(479969.35-589994.26) | 104.99<br>(99.61-110.10) | 9216.68<br>(4439.55-18587.97) | 19052.17<br>(9230.39-38296.94) | 106.71<br>(100.35-112.99) |
| Israel | 23382.42<br>(20990.96-25916.57) | 58968.86<br>(52884.43-65862.66) | 152.19<br>(144.28-158.83) | 312003.67<br>(280035.35-345863.29) | 821935.73<br>(742487.46-906819.96) | 163.44<br>(157.41-169.59) | 11092.53<br>(5293.47-22314.96) | 29412.45<br>(14049.79-59274.77) | 165.16<br>(157.89-173.16) |
| Italy | 415924.73<br>(369369.27-465184.00) | 595991.15<br>(530002.05-664508.76) | 43.29<br>(41.15-45.53) | 5826406.63<br>(5210209.46-6466884.18) | 9171839.29<br>(8231986.28-10143387.16) | 57.42<br>(55.59-59.46) | 206444.99<br>(99871.63-413613.59) | 328910.43<br>(159768.21-663710.96) | 59.32<br>(57.14-61.60) |
| Jamaica | 8487.99<br>(7542.14-9431.57) | 17246.76<br>(15255.65-19199.34) | 103.19<br>(96.78-110.22) | 111740.41<br>(99300.49-124096.12) | 221285.81<br>(197000.13-244088.71) | 98.04<br>(93.14-104.67) | 3937.80<br>(1874.53-7953.26) | 7804.75<br>(3738.67-15767.42) | 98.20<br>(92.19-105.78) |
| Japan | 1083158.09<br>(956009.66-1200740.90) | 1510140.68<br>(1348295.04-1664133.39) | 39.42<br>(34.48-44.67) | 13672546.22<br>(12142308.67-15127789.68) | 25378142.19<br>(22883664.81-27840274.16) | 85.61<br>(81.32-90.94) | 493607.60<br>(237042.85-994223.83) | 939484.99<br>(450468.54-1907691.61) | 90.33<br>(85.18-96.13) |
| Jordan | 7851.69<br>(6901.51-8788.87) | 51742.34<br>(45551.99-57344.94) | 559.00<br>(540.91-580.77) | 83781.76<br>(73939.73-93468.72) | 560038.55<br>(499447.26-624459.51) | 568.45<br>(550.94-589.85) | 2886.27<br>(1381.40-5762.06) | 19358.87<br>(9213.75-38956.11) | 570.72<br>(546.55-596.09) |

| | | | | | | | | | |
|---|---|---|---|---|---|---|---|---|---|
| Kazakhstan | 64184.63 (56185.52-72297.88) | 109192.03 (95013.72-122962.51) | 70.12 (64.66-75.47) | 831969.93 (719540.75-950666.79) | 1438645.55 (1254327.49-1650412.67) | 72.92 (67.20-78.26) | 29186.07 (13995.08-58956.77) | 51049.60 (24478.99-103160.59) | 74.91 (68.12-81.80) |
| Kenya | 43388.94 (38339.81-48283.16) | 151394.48 (132774.60-168327.27) | 248.92 (242.27-255.15) | 470145.02 (416631.36-523478.38) | 1600840.38 (1411451.46-1776032.71) | 240.50 (233.74-247.20) | 16150.49 (7727.88-32612.75) | 55737.26 (26430.41-112102.99) | 245.11 (238.04-252.68) |
| Kiribati | 223.50 (195.57-249.36) | 496.41 (435.91-555.58) | 122.10 (114.17-129.35) | 2482.17 (2202.99-2782.02) | 5506.49 (4882.55-6151.79) | 121.84 (114.77-128.62) | 85.41 (40.75-170.69) | 190.79 (91.17-379.01) | 123.39 (115.35-132.23) |
| Kuwait | 4803.24 (4239.96-5357.35) | 27283.61 (23711.49-30552.98) | 468.02 (442.77-492.71) | 44075.53 (39167.41-49291.28) | 256931.13 (225941.61-287094.89) | 482.93 (461.25-503.18) | 1529.23 (730.07-3069.46) | 8875.19 (4216.79-17984.91) | 480.37 (455.74-505.66) |
| Kyrgyzstan | 13633.64 (11891.22-15303.54) | 28256.31 (24309.26-32132.68) | 107.25 (99.94-115.49) | 182459.46 (158195.50-208786.05) | 347582.58 (298907.18-399945.42) | 90.50 (85.02-97.34) | 6396.76 (3090.37-12803.79) | 12275.84 (5873.37-24608.89) | 91.91 (85.22-99.46) |
| Lao People's Democratic Republic | 8442.30 (7402.99-9417.87) | 23166.86 (20272.10-25800.72) | 174.41 (164.05-186.83) | 94897.46 (84571.51-106999.75) | 256116.12 (225951.68-284449.16) | 169.89 (159.39-180.84) | 3222.47 (1550.70-6532.22) | 8798.74 (4204.83-17778.08) | 173.04 (161.05-186.52) |
| Latvia | 17577.14 (15584.46-19600.57) | 17147.20 (15281.60-19092.01) | -2.45 (-5.56-0.98) | 251241.20 (221943.88-281862.97) | 285051.22 (251601.01-319883.83) | 13.46 (10.47-16.49) | 8833.66 (4204.52-17933.79) | 10204.96 (4910.59-20813.73) | 15.52 (11.94-19.38) |
| Lebanon | 10501.69 (9259.15-11791.71) | 29680.77 (26182.25-32856.27) | 182.63 (172.77-192.39) | 125297.98 (110889.87-140499.66) | 386963.44 (345476.98-427385.05) | 208.83 (199.25-218.78) | 4298.07 (2050.71-8673.36) | 13309.11 (6394.88-26581.43) | 209.65 (199.23-222.41) |
| Lesotho | 4036.43 (3571.16-4497.67) | 6345.26 (5632.66-7046.41) | 57.20 (51.39-62.82) | 46613.76 (41366.31-52080.47) | 71755.68 (63483.90-79522.46) | 53.94 (48.53-59.40) | 1603.29 (774.79-3193.90) | 2454.86 (1172.19-4980.74) | 53.11 (46.74-59.15) |
| Liberia | 5475.55 (4876.03-6106.23) | 14829.84 (13025.39-16591.72) | 170.84 (158.37-183.61) | 62346.51 (55424.33-69475.24) | 144111.87 (128548.62-160569.46) | 131.15 (122.21-140.41) | 2099.93 (1008.99-4193.81) | 4885.42 (2358.73-9921.85) | 132.65 (122.45-142.59) |
| Libya | 10069.64 (8831.64-11224.24) | 35916.30 (31462.32-40045.09) | 256.68 (244.38-269.14) | 114661.77 (101657.63-126779.74) | 381938.86 (337903.69-424560.33) | 233.10 (223.48-243.77) | 3966.63 (1873.49-8054.90) | 13140.63 (6298.46-26535.06) | 231.28 (220.12-242.93) |
| Lithuania | 22206.94 (19615.64-24958.49) | 24886.23 (21924.02-27897.06) | 12.07 (8.44-15.70) | 312085.66 (274934.46-351694.31) | 412409.86 (363536.30-464231.18) | 32.15 (28.34-36.08) | 10970.20 (5259.83-22184.35) | 14729.69 (7077.75-30377.17) | 34.27 (30.12-38.97) |
| Luxembourg | 2603.67 (2328.11-2913.47) | 5250.30 (4678.89-5850.11) | 101.65 (96.09-108.16) | 35959.31 (32429.80-39645.32) | 72050.81 (64856.85-79546.68) | 100.37 (95.38-105.95) | 1277.16 (619.58-2581.01) | 2575.73 (1255.54-5199.88) | 101.68 (95.14-107.78) |
| Madagascar | 22700.59 (19850.57-25437.82) | 62379.53 (54683.44-69430.13) | 174.79 (164.84-186.21) | 246450.19 (218907.37-275234.82) | 618495.67 (547973.12-692259.24) | 150.96 (142.58-160.22) | 8359.98 (4039.87-16918.07) | 21115.81 (10250.18-42216.18) | 152.58 (141.81-162.51) |
| Malawi | 18371.91 (16138.62-20422.28) | 41598.86 (36586.74-46465.90) | 126.43 (118.89-134.63) | 196013.78 (174247.72-220147.51) | 430596.20 (381850.83-480035.16) | 119.68 (112.62-126.71) | 6653.06 (3212.43-13377.23) | 14742.49 (7111.09-29707.08) | 121.59 (113.48-130.10) |
| Malaysia | 46814.10 (41143.95-52105.40) | 149066.36 (130991.97-165936.76) | 218.42 (207.53-230.67) | 525256.35 (464559.50-582937.29) | 1844480.99 (1623467.15-2046052.29) | 251.16 (239.90-262.89) | 18052.20 (8536.87-36195.92) | 64150.46 (30558.32-130650.94) | 255.36 (241.63-268.96) |
| Maldives | 442.55 (384.45-497.09) | 2284.88 (1997.12-2541.56) | 416.30 (393.40-441.71) | 4977.03 (4373.61-5600.38) | 23523.03 (20543.55-26422.98) | 372.63 (354.72-389.97) | 171.52 (81.65-346.87) | 820.87 (388.85-1648.84) | 378.58 (357.88-398.51) |
| Mali | 18969.55 (16611.99-21293.85) | 48917.47 (42975.14-54786.19) | 157.87 (147.82-166.12) | 205635.09 (180947.42-232196.94) | 518280.50 (460243.85-576197.95) | 152.04 (142.22-160.14) | 6952.47 (3320.62-14004.14) | 17679.07 (8525.66-35194.22) | 154.28 (143.31-166.17) |
| Malta | 2205.31 (1957.06-2451.21) | 3974.10 (3570.22-4417.06) | 80.21 (72.71-88.09) | 28417.91 (25361.57-31464.91) | 61872.52 (55817.38-68505.28) | 117.72 (111.26-124.80) | 1011.24 (492.25-2019.69) | 2226.73 (1083.36-4473.32) | 120.20 (112.79-128.46) |
| Marshall Islands | 96.74 (85.36-106.98) | 254.01 (222.26-283.91) | 162.56 (151.37-172.16) | 1052.53 (930.58-1170.40) | 2734.54 (2405.68-3060.88) | 159.80 (151.94-167.69) | 36.41 (17.29-73.31) | 94.57(44.64-190.85) | 159.74 (150.52-169.63) |
| Mauritania | 4902.59 (4314.66-5482.70) | 12721.50 (11193.83-14180.53) | 159.49 (152.25-168.48) | 55960.72 (49822.40-62499.76) | 142473.72 (126133.12-157303.37) | 154.60 (147.60-163.03) | 1920.88 (932.12-3910.42) | 4959.34 (2362.72-10023.13) | 158.18 (149.04-167.60) |
| Mauritius | 3553.81 (3136.85-3951.78) | 8641.30 (7560.43-9666.42) | 143.16 (132.23-153.17) | 41941.83 (36871.79-46837.37) | 120219.34 (105220.27-134579.30) | 186.63 (177.27-194.94) | 1444.84 (684.57-2922.64) | 4163.99 (1992.20-8329.16) | 188.20 (176.75-198.04) |
| Mexico | 271007.03 (239139.82-301174.48) | 845738.53 (741869.99-936364.25) | 212.07 (206.23-217.58) | 3019238.13 (2680552.58-3343444.48) | 10208236.96 (9013451.69-11312756.72) | 238.11 (232.79-243.41) | 104802.83 (50161.79-212108.24) | 360985.49 (172240.82-727106.19) | 244.44 (237.98-250.84) |
| Micronesia (Federated States of) | 266.95 (236.53-296.26) | 499.60( 434.91-559.80) | 87.15 (78.98-94.73) | 3093.93 (2748.92-3428.92) | 5779.57 (5072.55-6444.79) | 86.80 (80.71-93.41) | 107.03 (50.78-214.09) | 201.32 (96.39-401.46) | 88.10 (81.08-96.23) |

| | | | | | | | | | |
|---|---|---|---|---|---|---|---|---|---|
| Monaco | 274.62 (246.27-306.83) | 391.11 (349.54-437.03) | 42.42 (38.63-47.07) | 4351.72 (3921.96-4804.52) | 6228.23 (5607.05-6869.27) | 43.12 (39.50-46.74) | 156.95 (75.43-319.92) | 225.05 (108.80-450.99) | 43.39 (38.90-48.00) |
| Mongolia | 4849.16 (4251.14-5444.52) | 15754.25 (13472.49-18098.73) | 224.89 (210.42-240.83) | 60106.12 (52373.38-68053.96) | 183223.00 (157823.06-212343.29) | 204.83 (190.37-219.90) | 2094.75 (1000.87-4190.42) | 6452.60 (3066.68-13168.92) | 208.04 (192.30-224.88) |
| Montenegro | 3187.67 (2803.09-3554.85) | 4653.46 (4083.11-5215.08) | 45.98 (40.69-51.02) | 41463.57 (36197.10-46588.69) | 67345.04 (59160.38-75352.44) | 62.42 (57.72-67.21) | 1460.24 (699.99-2946.32) | 2383.02 (1146.27-4867.88) | 63.19 (57.47-68.92) |
| Morocco | 68064.12 (60211.80-76074.19) | 180998.03 (159986.89-202804.35) | 165.92 (156.87-176.04) | 795767.21 (706124.42-884295.97) | 2171352.04 (1923170.17-2402600.66) | 172.86 (164.82-181.88) | 27325.43 (13115.79-55791.70) | 74163.92 (35516.30-151542.03) | 171.41 (162.34-182.81) |
| Mozambique | 28521.44 (25086.65-31791.52) | 62053.04 (54601.24-69190.88) | 117.57 (109.43-125.67) | 302314.65 (266627.31-338109.88) | 638138.13 (563557.68-713789.51) | 111.08 (103.36-118.04) | 10166.69 (4848.95-20528.77) | 21568.69 (10342.31-43534.45) | 112.15 (103.44-120.83) |
| Myanmar | 94100.31 (83053.46-104575.74) | 233234.37 (205139.10-260510.89) | 147.86 (137.34-157.87) | 1077954.41 (954126.96-1205274.71) | 2815942.99 (2485900.57-3143670.43) | 161.23 (150.68-171.18) | 36582.99 (17525.52-74377.90) | 96779.11 (46237.61-192236.94) | 164.55 (152.23-175.85) |
| Namibia | 3250.46 (2855.14-3639.44) | 8343.57 (7333.97-9309.93) | 156.69 (147.32-165.53) | 36282.00 (32170.08-40576.45) | 91168.76 (81462.02-101415.01) | 151.28 (142.87-159.19) | 1251.09 (609.87-2523.89) | 3149.25 (1497.08-6318.67) | 151.72 (141.38-161.43) |
| Nauru | 30.51 (26.65-34.04) | 42.40 (37.22-47.33) | 38.99 (33.95-45.13) | 318.47 (282.71-355.55) | 457.91 (405.26-513.28) | 43.79 (39.33-49.21) | 11.06 (5.26-22.08) | 15.92 (7.60-31.64) | 43.96 (38.27-50.39) |
| Nepal | 44228.63 (39302.50-49166.64) | 115211.17 (102658.20-128001.50) | 160.49 (151.56-170.05) | 477016.06 (423058.26-531853.49) | 1354754.22 (1202295.88-1508078.41) | 184.01 (174.92-193.30) | 15938.63 (7779.87-32320.51) | 45782.02 (22135.94-92351.74) | 187.24 (176.24-199.29) |
| Netherlands | 97291.47 (87433.12-107937.62) | 154895.17 (137844.91-173273.19) | 59.21 (51.57-66.37) | 1351065.82 (1239207.08-1478700.98) | 2323332.19 (2095824.25-2566694.80) | 71.96 (64.62-79.46) | 48435.51 (23580.36-97160.91) | 83317.18 (40222.11-169267.26) | 72.02 (63.36-81.34) |
| New Zealand | 20650.68 (18415.42-22849.17) | 44371.59 (39437.71-49394.72) | 114.87 (109.14-121.37) | 275881.74 (247471.52-305446.77) | 627576.84 (562573.67-693133.76) | 127.48 (122.18-133.08) | 9758.67 (4729.71-19669.07) | 22600.28 (10845.75-45607.20) | 131.59 (125.62-138.66) |
| Nicaragua | 8667.75 (7627.60-9609.35) | 30198.12 (26577.04-33715.71) | 248.40 (237.31-261.52) | 95091.97 (84560.40-105551.04) | 348503.36 (308821.41-386203.33) | 266.49 (256.07-279.38) | 3277.05 (1582.84-6601.62) | 12087.17 (5854.33-24321.79) | 268.84 (256.14-282.76) |
| Niger | 14143.62 (12458.62-15854.20) | 43201.64 (38133.16-48220.56) | 205.45 (195.20-217.65) | 143841.00 (127677.95-160715.60) | 454880.92 (401577.73-510376.47) | 216.24 (206.26-227.87) | 4875.55 (2377.20-9773.40) | 15513.57 (7468.38-31212.23) | 218.19 (205.26-231.37) |
| Nigeria | 227291.16 (200073.97-254306.46) | 593768.16 (521287.00-662832.29) | 161.24 (156.44-165.97) | 2538174.06 (2252320.71-2836865.39) | 6171576.50 (5479715.56-6850080.01) | 143.15 (139.29-147.07) | 86775.39 (41754.81-174519.99) | 213565.72 (102289.81-428806.70) | 146.11 (141.84-150.32) |
| Niue | 10.74 (9.49-11.97) | 12.34 (10.90-13.75) | 14.96 (10.65-19.30) | 146.90 (130.74-164.72) | 168.54 (148.03-187.40) | 14.73 (11.22-18.79) | 5.12 (2.45-10.30) | 5.91 (2.86-11.79) | 15.34 (11.47-19.95) |
| North Macedonia | 9175.41 (8090.51-10209.20) | 16079.71 (14210.58-18001.51) | 75.25 (69.87-81.05) | 113854.61 (100497.19-127263.64) | 220221.20 (194054.80-247312.66) | 93.42 (88.29-98.18) | 3957.22 (1891.12-8022.74) | 7720.21 (3729.91-15666.96) | 95.09 (88.30-101.32) |
| Northern Mariana Islands | 165.37 (145.23-185.03) | 361.82 (314.37-409.81) | 118.80 (104.16-136.00) | 1449.75 (1277.38-1629.58) | 4360.66 (3861.99-4852.81) | 200.79 (187.18-216.07) | 50.74 (24.28-102.84) | 153.78 (72.67-307.87) | 203.10 (188.24-221.32) |
| Norway | 28574.34 (25448.95-31541.83) | 45754.69 (40756.00-51003.94) | 60.13 (57.31-62.81) | 426724.99 (382913.51-472687.54) | 665457.98 (598229.62-735909.27) | 55.95 (54.24-57.59) | 15180.68 (7350.49-30580.07) | 23939.19 (11585.31-48114.64) | 57.70 (55.42-59.61) |
| Oman | 4163.80 (3667.82-4635.66) | 18373.47 (16230.35-20498.67) | 341.27 (325.45-356.71) | 41160.90 (36553.33-46150.93) | 166344.32 (148368.10-186292.88) | 304.13 (291.21-317.43) | 1408.82 (676.59-2859.37) | 5753.23 (2747.36-11540.36) | 308.37 (292.54-325.41) |
| Pakistan | 249293.45 (218771.59-280263.07) | 710195.02 (618261.31-798895.91) | 184.88 (178.11-192.33) | 2854291.85 (2512824.79-3197228.70) | 7627598.23 (6679952.77-8492196.76) | 167.23 (161.43-173.69) | 96928.72 (46086.79-195297.34) | 261199.60 (124700.72-522198.52) | 169.48 (162.54-178.17) |
| Palau | 59.36 (52.31-65.82) | 155.67 (135.54-175.92) | 162.24 (150.10-174.65) | 684.56 (609.74-761.93) | 1907.48 (1680.63-2138.60) | 178.64 (169.63-188.22) | 23.89 (11.35-48.86) | 66.63 (31.77-132.66) | 178.91 (168.92-189.97) |
| Palestine | 4112.41 (3617.44-4598.69) | 15980.91 (14017.08-17900.93) | 288.60 (274.67-304.90) | 47974.07 (42524.49-53957.46) | 174021.23 (153510.79-193335.41) | 262.74 (249.78-276.29) | 1643.55 (797.10-3289.82) | 5968.35 (2849.38-12076.82) | 263.14 (247.71-278.96) |
| Panama | 8020.19 (7082.41-8912.34) | 24713.84 (21961.12-27430.65) | 208.15 (197.58-218.29) | 93919.84 (83215.42-104408.50) | 315880.70 (280593.22-347772.08) | 236.33 (226.55-244.96) | 3269.32 (1561.63-6569.60) | 11102.53 (5340.56-22255.18) | 239.60 (227.28-251.43) |
| Papua New Guinea | 9687.45 (8500.77-10874.23) | 32236.46 (28233.33-36149.52) | 232.77 (219.18-246.25) | 101622.67 (89857.07-113863.98) | 328342.50 (290264.50-366544.41) | 223.10 (210.25-234.47) | 3445.04 (1655.09-6904.44) | 11205.47 (5416.85-22417.12) | 225.26 (212.40-239.34) |

| | | | | | | | | | |
|---|---|---|---|---|---|---|---|---|---|
| Paraguay | 12880.63 (11366.87-14299.89) | 35220.61 (31206.44-39088.60) | 173.44 (164.76-182.73) | 150830.49 (133482.46-166899.67) | 420797.00 (371504.60-467033.14) | 178.99 (171.62-186.97) | 5275.53 (2519.57-10630.20) | 14700.30 (6967.63-29805.12) | 178.65 (169.23-188.16) |
| Peru | 70517.17 (62193.27-78558.21) | 205559.36 (180552.05-227854.12) | 191.50 (181.13-200.96) | 813102.61 (720158.97-903442.32) | 2517755.09 (2228671.61-2782727.44) | 209.65 (200.65-217.98) | 28553.75 (13791.43-57638.06) | 89320.44 (42787.35-180730.52) | 212.82 (202.71-222.65) |
| Philippines | 136804.55 (119814.54-152613.58) | 419468.91 (366177.69-471535.90) | 206.62 (201.44-211.53) | 1518301.16 (1328784.72-1694013.52) | 4905958.00 (4283100.35-5490220.80) | 223.12 (218.94-227.42) | 51853.01 (24542.09-104045.22) | 169673.12 (80661.21-339752.86) | 227.22 (222.33-232.95) |
| Poland | 208141.04 (183527.77-232356.05) | 328301.95 (290504.70-366346.01) | 57.73 (54.74-60.98) | 2760678.31 (2435920.37-3094921.33) | 4892574.97 (4294492.74-5479011.74) | 77.22 (74.73-79.90) | 96022.36 (46325.16-193634.29) | 173973.80 (84080.61-351958.02) | 81.18 (78.28-84.07) |
| Portugal | 64091.05 (57341.64-71476.21) | 99153.66 (88674.31-109697.25) | 54.71 (49.75-59.63) | 879207.36 (785026.09-974590.00) | 1531034.98 (1387657.91-1690288.92) | 74.14 (69.47-78.59) | 30963.47 (14989.00-62443.28) | 54607.12 (26327.40-109973.50) | 76.36 (70.77-82.23) |
| Puerto Rico | 20663.80 (18419.37-23018.99) | 32518.89 (29014.96-35959.46) | 57.37 (51.32-63.28) | 267146.76 (238537.30-295265.73) | 512073.83 (458024.98-565681.30) | 91.68 (85.85-97.74) | 9504.52 (4541.60-19067.25) | 18333.09 (8816.43-37052.57) | 92.89 (85.83-100.01) |
| Qatar | 1287.83 (1128.22-1448.67) | 13822.30 (12031.47-15488.75) | 973.30 (928.64-1023.88) | 10014.12 (8795.68-11369.36) | 108211.42 (95937.41-122252.93) | 980.59 (936.34-1027.55) | 345.83 (164.48-698.60) | 3732.31 (1781.75-7558.47) | 979.22 (923.78-1041.07) |
| Republic of Korea | 243702.24 (215123.00-270409.48) | 615471.97 (547205.31-688215.64) | 152.55 (143.06-162.88) | 2692411.38 (2400450.42-2973646.95) | 8441324.38 (7578238.02-9289492.95) | 213.52 (203.94-224.62) | 96296.45 (46362.81-195129.91) | 307676.79 (148214.59-623772.02) | 219.51 (208.47-230.77) |
| Republic of Moldova | 21213.86 (18723.08-23577.43) | 28864.06 (25566.45-32253.82) | 36.06 (32.25-40.46) | 273920.89 (241769.33-305215.09) | 414221.66 (365281.13-462181.94) | 51.22 (47.01-55.91) | 9526.52 (4559.95-19320.81) | 14657.60 (7021.23-29353.45) | 53.86 (48.72-59.50) |
| Romania | 125422.77 (110912.37-140299.44) | 151599.31 (133962.17-168487.14) | 20.87 (17.10-25.42) | 1664333.33 (1465507.82-1861361.16) | 2276904.19 (2014050.11-2545520.16) | 36.81 (33.12-41.01) | 57921.93 (27934.07-116376.07) | 80579.54 (38213.16-163255.15) | 39.12 (34.42-44.15) |
| Russian Federation | 1004302.62 (881009.94-1125128.18) | 1283831.31 (1130169.17-1446183.75) | 27.83 (25.66-30.34) | 14229933.06 (12375792.60-16136118.40) | 18882624.58 (16518692.41-21233249.88) | 32.70 (30.03-35.13) | 502936.73 (239783.88-1018932.61) | 673207.72 (324711.53-1363422.48) | 33.86 (31.30-36.71) |
| Rwanda | 13013.08 (11506.58-14539.83) | 33947.88 (29699.02-37790.72) | 160.87 (151.46-170.59) | 138831.39 (123262.68-155502.75) | 354898.61 (313940.33-399397.96) | 155.63 (146.71-162.84) | 4711.85 (2267.34-9557.50) | 12116.23 (5847.10-24410.12) | 157.14 (146.18-166.98) |
| Saint Kitts and Nevis | 168.81 (149.56-186.78) | 467.83 (410.28-523.06) | 177.14 (161.67-191.70) | 2440.56 (2170.39-2706.46) | 5643.42 (4965.64-6285.03) | 131.23 (122.00-140.32) | 86.18 (41.12-173.38) | 198.93 (94.48-400.84) | 130.85 (121.03-140.44) |
| Saint Lucia | 443.80 (395.10-492.27) | 1384.94 (1220.62-1539.29) | 212.07 (199.86-225.23) | 5545.16 (4924.90-6182.49) | 17764.36 (15747.46-19673.52) | 220.36 (210.56-231.21) | 192.59 (92.73-392.45) | 622.82 (300.17-1257.51) | 223.40 (212.24-235.46) |
| Saint Vincent and the Grenadines | 348.13 (311.61-387.74) | 798.14 (705.29-892.81) | 129.27 (120.53-137.02) | 4460.46 (3969.73-4938.32) | 10319.11 (9104.70-11423.73) | 131.35 (124.44-137.97) | 155.81 (74.14-313.77) | 361.31 (174.89-743.54) | 131.89 (124.12-140.18) |
| Samoa | 477.17 (419.44-532.85) | 905.55 (795.80-1019.18) | 89.77 (83.27-96.48) | 5669.06 (5062.90-6305.32) | 10896.83 (9617.00-12172.10) | 92.22 (85.93-98.10) | 196.53 (94.07-390.05) | 379.73 (181.52-754.24) | 93.22 (86.52-100.79) |
| San Marino | 159.65 (142.43-178.00) | 314.00 (280.47-351.39) | 96.68 (90.95-102.72) | 2272.43 (2046.73-2510.85) | 4760.39 (4286.57-5293.62) | 109.48 (104.37-114.13) | 81.12 (39.67-162.95) | 170.57 (82.73-345.03) | 110.27 (104.48-116.24) |
| Sao Tome and Principe | 299.17 (264.02-335.07) | 763.89 (668.78-851.29) | 155.34 (144.02-167.53) | 3683.85 (3245.27-4140.47) | 8195.01 (7248.72-9176.89) | 122.46 (113.49-131.28) | 127.40 (61.30-262.05) | 287.00 (137.57-579.95) | 125.28 (115.67-135.31) |
| Saudi Arabia | 35182.44 (30925.89-39296.34) | 184905.73 (161454.22-206867.00) | 425.56 (405.17-447.36) | 358102.26 (317568.12-400082.29) | 1669212.28 (1468855.45-1864087.27) | 366.13 (348.09-385.57) | 12217.23 (5871.01-24414.47) | 57366.54 (27050.04-114815.17) | 369.55 (348.03-393.15) |
| Senegal | 16080.31 (14163.01-17955.18) | 44160.78 (38545.73-49138.95) | 174.63 (165.18-183.83) | 178535.82 (158271.63-200005.13) | 487696.08 (431955.89-543741.15) | 173.16 (164.73-181.09) | 6090.89 (2937.01-12334.38) | 16773.28 (8031.28-33485.34) | 175.38 (165.18-185.37) |
| Serbia | 53619.46 (47007.88-60118.14) | 68586.13 (60636.78-76247.57) | 27.91 (22.49-32.89) | 688243.42 (601329.22-772951.61) | 1032459.01 (910056.15-1152671.49) | 50.01 (45.33-55.03) | 24007.24 (11468.56-48135.34) | 36404.12 (17445.25-73708.35) | 51.64 (46.34-57.67) |
| Seychelles | 232.31 (205.62-258.46) | 634.96 (551.71-712.61) | 173.32 162.27-185.31) | 3101.34 (2740.11-3456.96) | 8011.72 (7010.33-8933.87) | 158.33 (149.38-167.44) | 107.65 (51.17-214.87) | 278.60 (132.98-559.59) | 158.80 (147.84-170.25) |
| Sierra Leone | 9246.96 (8198.20-10287.02) | 21581.78 (18975.70-23937.05) | 133.39 (123.66-142.29) | 104473.55 (92952.49-117306.42) | 228398.68 (202456.59-254358.92) | 118.62 (110.83-126.66) | 3537.73 (1709.10-7084.24) | 7830.04 (3753.43-15712.36) | 121.33 (112.40-130.68) |
| Singapore | 18009.23 (15810.41-20088.41) | 60586.55 (53459.98-67210.64) | 236.42 (224.14-249.52) | 198983.10 (176481.59-219345.47) | 772440.20 (686704.35-849720.39) | 288.19 (276.56-300.33) | 7221.64 (3412.38-14317.95) | 28430.14 (13694.28-56974.23) | 293.68 (280.62-307.70) |

| | | | | | | | | | |
|---|---|---|---|---|---|---|---|---|---|
| Slovakia | 28455.15<br>(25090.16-31773.66) | 44551.07<br>(39163.91-49636.20) | 56.57<br>(52.36-61.31) | 391659.09<br>(342919.43-440887.94) | 659576.96<br>(581867.78-740170.59) | 68.41<br>(64.29-72.95) | 13705.87<br>(6487.80-27895.00) | 23394.74<br>(11270.21-47392.67) | 70.69<br>(65.48-76.67) |
| Slovenia | 11790.01<br>(10383.91-13186.03) | 18194.62<br>(16122.93-20341.77) | 54.32<br>(49.35-59.92) | 162734.08<br>(142848.87-182921.73) | 287180.50<br>(252520.00-321638.69) | 76.47<br>(71.47-82.19) | 5682.82<br>(2694.50-11430.93) | 10172.71<br>(4917.64-20700.68) | 79.01<br>(72.78-85.13) |
| Solomon Islands | 761.91<br>(666.37-855.14) | 2344.49<br>(2039.23-2634.05) | 207.71<br>(195.95-219.43) | 8226.55<br>(7301.90-9179.61) | 24551.69<br>(21729.56-27515.86) | 198.44<br>(188.35-209.19) | 282.87<br>(135.04-567.00) | 848.82<br>(404.33-1696.17) | 200.07<br>(187.20-212.59) |
| Somalia | 13590.69<br>(11943.50-15233.76) | 35857.13<br>(31434.54-40006.35) | 163.84<br>(153.40-172.64) | 126940.14<br>(112767.45-141973.34) | 339051.60<br>(300120.42-377062.39) | 167.10<br>(157.18-175.51) | 4309.51<br>(2104.87-8656.56) | 11522.88<br>(5543.47-23309.90) | 167.38<br>(155.66-177.81) |
| South Africa | 125033.34<br>(110149.67-139039.89) | 306661.87<br>(269896.02-340954.90) | 145.26<br>(142.05-148.42) | 1443349.54<br>(1271318.53-1603518.53) | 3566080.34<br>(3135481.25-3969604.47) | 147.07<br>(144.45-149.98) | 50501.82<br>(24157.90-101375.11) | 124233.25<br>(59848.76-248721.69) | 146.00<br>(142.50-149.81) |
| South Sudan | 11184.46<br>(9857.93-12478.83) | 22010.08<br>(19217.40-24633.32) | 96.79<br>(88.60-106.23) | 124312.41<br>(110890.20-139114.36) | 219211.11<br>(193939.53-245024.42) | 76.34<br>(70.24-84.31) | 4165.06<br>(2024.89-8314.49) | 7379.15<br>(3595.24-14771.78) | 77.17<br>(69.35-86.04) |
| Spain | 247727.10<br>(219689.38-278141.53) | 420976.27<br>(376268.79-470250.03) | 69.94<br>(64.64-75.27) | 3489572.39<br>(3139552.81-3872738.53) | 6279574.92<br>(5668450.51-6993098.45) | 79.95<br>(75.05-84.92) | 123887.67<br>(59793.30-248851.82) | 224792.41<br>(108762.63-454455.69) | 81.45<br>(75.55-87.49) |
| Sri Lanka | 48932.05<br>(42912.10-54503.09) | 117977.47<br>(103434.96-131691.11) | 141.10<br>(129.13-151.89) | 552485.89<br>(488604.28-613795.84) | 1575155.30<br>(1380242.47-1763543.12) | 185.10<br>(173.32-196.40) | 18854.04<br>(8981.17-38305.20) | 54025.08<br>(25576.21-108718.84) | 186.54<br>(173.30-198.59) |
| Sudan | 39608.54<br>(34928.25-44228.58) | 114477.63<br>(100690.06-127670.73) | 189.02<br>(175.76-200.28) | 438043.32<br>(388820.35-493519.15) | 1200374.40<br>(1063278.17-1331239.81) | 174.03<br>(163.54-183.75) | 14803.80<br>(7071.43-29773.48) | 41062.32<br>(19773.84-82320.50) | 177.38<br>(164.67-190.41) |
| Suriname | 1531.56<br>(1338.49-1705.69) | 3896.00<br>(3417.28-4354.40) | 154.38<br>(146.10-163.33) | 18172.65<br>(16038.43-20192.34) | 48941.48<br>(43092.77-54493.92) | 169.31<br>(161.99-176.73) | 639.83<br>(304.42-1303.95) | 1719.06<br>(825.94-3439.02) | 168.67<br>(159.66-178.26) |
| Sweden | 54140.08<br>(47891.60-60494.79) | 77849.10<br>(68631.12-87509.92) | 43.79<br>(39.95-48.13) | 797704.83<br>(708550.50-892345.98) | 1188697.73<br>(1047613.18-1329028.25) | 49.01<br>(45.46-52.97) | 28113.81<br>(13222.87-57854.31) | 42255.42<br>(20045.75-86461.02) | 50.30<br>(46.01-55.44) |
| Switzerland | 46178.84<br>(41057.31-51247.02) | 76109.05<br>(67951.25-85016.38) | 64.81<br>(60.25-69.53) | 656863.76<br>(587681.65-726269.15) | 1131233.23<br>(1021569.54-1251299.83) | 72.22<br>(68.68-76.52) | 23207.91<br>(11273.75-46797.74) | 40161.71<br>(19424.56-80499.55) | 73.05<br>(68.71-78.28) |
| Syrian Arab Republic | 25783.58<br>(22778.63-28804.98) | 75626.00<br>(65836.59-85154.65) | 193.31<br>(182.20-204.14) | 287369.10<br>(254501.04-319697.49) | 890644.41<br>(783272.90-991866.23) | 209.93<br>(199.89-221.12) | 9839.91<br>(4763.46-19630.86) | 30700.65<br>(14734.72-60901.80) | 212.00<br>(199.96-225.60) |
| Taiwan<br>(Province of China) | 91954.12<br>(80996.16-102954.22) | 223448.81<br>(197853.78-249985.94) | 143.00<br>(133.46-152.71) | 1104590.49<br>(973881.96-1230178.29) | 3215060.50<br>(2855621.84-3595342.09) | 191.06<br>(180.03-200.99) | 38585.33<br>(18394.05-77245.94) | 113637.80<br>(53956.35-229872.21) | 194.51<br>(182.77-205.48) |
| Tajikistan | 11677.89<br>(10186.64-13142.76) | 32556.00<br>(27991.38-36814.38) | 178.78<br>(169.13-189.66) | 147684.49<br>(128100.46-167793.36) | 370401.52<br>(318735.22-423393.26) | 150.81<br>(142.97-160.05) | 5145.09<br>(2472.05-10394.73) | 12933.01<br>(6153.48-25852.96) | 151.37<br>(141.48-162.32) |
| Thailand | 165162.40<br>(144618.60-183600.46) | 484227.60<br>(425932.22-544644.62) | 193.18<br>(179.27-208.71) | 1843097.36<br>(1638257.86-2053298.15) | 6612397.30<br>(5856915.79-7391249.39) | 258.77<br>(243.45-274.24) | 62734.43<br>(30600.65-126180.47) | 229188.05<br>(110605.94-460753.16) | 265.33<br>(248.22-283.83) |
| Timor-Leste | 1404.89<br>(1231.69-1580.46) | 3682.14<br>(3243.76-4097.61) | 162.09<br>(148.84-175.15) | 13763.05<br>(12197.27-15556.85) | 45112.88<br>(39773.29-50300.77) | 227.78<br>(211.13-243.45) | 465.96<br>(223.63-940.86) | 1540.45<br>(732.52-3120.03) | 230.59<br>(212.28-249.35) |
| Togo | 6576.50<br>(5782.57-7296.69) | 24075.39<br>(21234.34-26922.44) | 266.08<br>(252.15-282.26) | 67670.76<br>(60129.75-75120.40) | 247407.54<br>(218789.11-274359.39) | 265.60<br>(253.27-279.24) | 2304.32<br>(1109.67-4594.26) | 8532.45<br>(4036.24-17272.44) | 270.28<br>(255.43-287.29) |
| Tokelau | 6.10<br>(5.37-6.80) | 8.04<br>(7.09-8.91) | 31.72<br>(26.77-36.80) | 82.32<br>(72.55-91.93) | 108.22<br>(96.19-120.06) | 31.47<br>(27.20-35.66) | 2.85<br>(1.38-5.75) | 3.78<br>(1.82-7.63) | 32.42<br>(27.66-37.59) |
| Tonga | 302.82<br>(264.80-338.43) | 469.41<br>(411.80-523.53) | 55.01<br>(49.39-61.72) | 3637.25<br>(3214.20-4040.66) | 5826.86<br>(5172.23-6501.42) | 60.20<br>(54.26-66.45) | 126.33<br>(60.36-253.93) | 203.08<br>(97.57-407.99) | 60.75<br>(54.68-67.63) |
| Trinidad and Tobago | 4879.54<br>(4312.09-5457.58) | 10980.43<br>(9706.65-12243.36) | 125.03<br>(116.98-133.51) | 58782.23<br>(52329.44-65064.47) | 145745.99<br>(129485.65-161267.12) | 147.94<br>(140.87-154.94) | 2063.41<br>(981.40-4166.60) | 5133.24<br>(2478.02-10560.20) | 148.77<br>(140.58-157.57) |
| Tunisia | 23828.30<br>(21005.64-26755.88) | 69720.70<br>(61709.51-77559.69) | 192.60<br>(183.05-201.60) | 281097.11<br>(247994.98-314359.38) | 874118.18<br>(773391.35-972641.97) | 210.97<br>(200.70-219.91) | 9675.15<br>(4593.69-19642.87) | 30167.23<br>(14549.09-61097.78) | 211.80<br>(200.33-223.75) |
| Turkmenistan | 9069.78<br>(7961.31-10234.85) | 24003.35<br>(20721.06-27399.20) | 190.02<br>(179.99-199.97) | 112657.2<br>1(97731.31-128117.03) | 297030.64<br>(255487.29-341108.94) | 220.77<br>(209.91-231.69) | 3945.73<br>(1888.90-7987.89) | 10538.16<br>(4990.58-20938.36) | 224.68<br>(211.84-237.30) |
| Tuvalu | 35.95<br>(31.51-40.14) | 60.96<br>(53.46-68.28) | 164.65<br>(154.74-174.86) | 426.99<br>(378.06-476.47) | 768.14<br>(676.67-861.37) | 163.66<br>(154.50-173.16) | 14.80<br>(7.05-30.14) | 26.89<br>(12.73-54.97) | 167.08<br>(156.11-178.95) |

| | | | | | | | | | |
|---|---|---|---|---|---|---|---|---|---|
| Türkiye | 172034.55<br>(151349.54-191269.98) | 498940.01<br>(442251.12-557033.39) | 69.56<br>(64.11-75.32) | 1953405.25<br>(1735773.55-2176900.05) | 6265937.25<br>(5560635.54-6937782.92) | 79.90<br>(74.17-85.74) | 67006.79<br>(32365.54-134315.34) | 217554.77<br>(104524.79-438712.79) | 81.67<br>(74.08-88.33) |
| Uganda | 29417.51<br>(25827.76-32999.61) | 82029.48<br>(72113.17-91565.95) | 178.85<br>(169.01-189.07) | 319755.63<br>(282997.68-358252.92) | 846302.42<br>(748399.72-943276.88) | 164.67<br>(156.20-173.89) | 10808.16<br>(5230.93-21752.40) | 28900.96<br>(13959.74-58727.41) | 167.40<br>(156.98-178.92) |
| Ukraine | 356545.13<br>(312759.74-401589.88) | 386110.14<br>(341240.92-431370.24) | 8.29<br>(4.79-11.78) | 4936819.25<br>(4325172.58-5555751.54) | 5699623.83<br>(5010776.96-6405476.83) | 15.45<br>(12.32-18.74) | 173385.45<br>(84207.03-350221.67) | 202302.15<br>(97312.40-406819.99) | 16.68<br>(12.84-20.50) |
| United Arab Emirates | 4548.70<br>(3970.18-5115.42) | 63707.74<br>(55393.97-72162.23) | 1300.57<br>(1235.14-1367.01) | 35269.13<br>(30792.82-40075.35) | 503960.12<br>(441962.32-572572.31) | 1328.90<br>(1279.20-1386.76) | 1210.94<br>(573.69-2457.77) | 17355.80<br>(8125.42-34938.74) | 1333.25<br>(1269.76-1409.17) |
| United Kingdom | 416553.10<br>(371985.22-462864.40) | 610070.33<br>(544797.67-679814.06) | 46.46<br>(45.00-48.10) | 6004365.47<br>(5411317.39-6647048.93) | 8953548.47<br>(8063136.45-9891755.23) | 49.12<br>(48.13-50.13) | 215272.50<br>(104015.22-432200.57) | 322643.41<br>(156642.45-645022.18) | 49.88<br>(48.75-51.22) |
| United Republic of Tanzania | 53271.94<br>(46769.39-59336.83) | 145866.52<br>(129292.07-162102.24) | 173.81<br>(165.16-183.46) | 593225.19<br>(524534.28-660943.80) | 1545396.50<br>(1374111.04-1718529.93) | 160.51<br>(152.90-168.67) | 20257.16<br>(9681.26-40379.52) | 53090.81<br>(25271.55-107594.85) | 162.08<br>(153.93-172.21) |
| United States of America | 1770581.66<br>(1582305.58-1951370.12) | 3192701.53<br>(2830531.82-3554075.01) | 80.32<br>(76.28-84.38) | 25007735.07<br>(22549194.96-27580854.86) | 47581100.89<br>(42638857.15-52684415.20) | 90.27<br>(88.59-91.99) | 900534.50<br>(432395.48-1819200.41) | 1710760.12<br>(829259.16-3463603.19) | 89.97<br>(87.94-92.55) |
| United States Virgin Islands | 579.91<br>(510.16-647.66) | 889.40<br>(784.36-995.15) | 53.37<br>(44.06-63.39) | 6535.50<br>(5788.18-7239.81) | 13567.14<br>(12092.65-14990.09) | 107.59<br>(98.62-116.16) | 231.07<br>(110.26-463.72) | 483.12<br>(234.06-963.15) | 109.08<br>(99.28-118.70) |
| Uruguay | 19163.75<br>(17021.81-21377.55) | 26842.26<br>(23942.06-29965.55) | 40.07<br>(35.52-44.22) | 266618.11<br>(236979.95-296555.43) | 392380.61<br>(354172.81-436252.38) | 47.17<br>(42.91-51.02) | 9484.71<br>(4532.01-18884.85) | 14039.33<br>(6794.96-27922.33) | 48.02<br>(43.39-53.23) |
| Uzbekistan | 53136.92<br>(46259.39-59857.27) | 159659.3<br>2(137465.65-180474.79) | 200.47<br>(190.01-211.13) | 691164.84<br>(600001.03-784589.83) | 1921520.82<br>(1657298.21-2197996.32) | 178.01<br>(168.83-186.88) | 24208.24<br>(11686.02-48890.35) | 67713.83<br>(32353.59-135488.50) | 179.71<br>(169.26-190.17) |
| Vanuatu | 343.15<br>(299.16-381.29) | 1056.32<br>(930.56-1190.22) | 207.83<br>(197.39-219.03) | 3586.88<br>(3152.36-3991.89) | 11429.51<br>(10100.44-12778.89) | 218.65<br>(208.66-229.30) | 122.57<br>(59.64-248.52) | 392.31<br>(190.30-791.27) | 220.06<br>(209.04-232.36) |
| Venezuela (Bolivarian Republic of) | 59833.32<br>(53027.77-66428.49) | 180660.39<br>(159653.17-201258.61) | 201.94<br>(191.13-212.22) | 669767.47<br>(597380.95-742736.58) | 2255788.02<br>(1984530.74-2494580.22) | 236.80<br>(227.43-247.01) | 23413.93<br>(11252.96-46677.34) | 79429.29<br>(38221.46-160696.88) | 239.24<br>(227.47-251.71) |
| Viet Nam | 145579.11<br>(128053.35-162687.98) | 448882.04<br>(391922.63-502050.10) | 208.34<br>(194.63-221.30) | 1779684.81<br>(1572372.51-1987537.22) | 5415246.51<br>(4745047.23-6045659.66) | 204.28<br>(191.72-217.02) | 60615.15<br>(29204.81-120769.91) | 187214.70<br>(89043.91-373119.41) | 208.86<br>(194.03-224.80) |
| Yemen | 21538.37<br>(18960.91-23990.46) | 77884.98<br>(68746.92-86681.40) | 261.61<br>(247.20-275.23) | 227270.32<br>(201431.08-254487.85) | 804925.40<br>(712722.05-894558.83) | 254.17<br>(240.87-267.03) | 7635.07<br>(3681.79-15593.27) | 27348.24<br>(13108.70-55052.66) | 258.19<br>(242.61-274.47) |
| Zambia | 14991.90<br>(13119.26-16794.84) | 42992.71<br>(37783.47-48108.27) | 186.77<br>(176.34-197.10) | 159770.29<br>(141651.09-177776.16) | 429173.59<br>(380416.15-476903.19) | 168.62<br>(160.87-177.32) | 5468.47<br>(2599.24-11123.08) | 14661.58<br>(7052.48-29519.94) | 168.11<br>(158.38-178.98) |
| Zimbabwe | 20556.84<br>(18220.64-22841.63) | 39757.63<br>(34863.72-44366.30) | 93.40<br>(86.57-100.04) | 224498.10<br>(199246.66-250423.26) | 409820.92<br>(362489.52-457873.97) | 82.55<br>(76.69-88.27) | 7714.41<br>(3733.54-15456.25) | 13961.24<br>(6714.89-27908.04) | 80.98<br>(74.02-88.78) |

### Table S5. The ASRs and EAPCs of osteoarthritis among 204 countries and territories from 1990 to 2021.

| Country | ASIR (per 100,000 population) | | | ASPR (per 100,000 population) | | | ASDR (per 100,000 population) | | |
|---|---|---|---|---|---|---|---|---|---|
| | 1990 | 2021 | EAPC (95%CI) | 1990 | 2021 | EAPC (95%CI) | 1990 | 2021 | EAPC (95%CI) |
| | Both (95%UI) | Both (95%UI) | | Both (95%UI) | Both (95%UI) | | Both (95%UI) | Both (95%UI) | |
| Afghanistan | 369.68 (327.35-412.51) | 409.68 (362.26-460.31) | 0.40 (0.36-0.44) | 4551.49 (4037.46-5103.92) | 5109.54 (4529.06-5687.19) | 0.44 (0.40-0.49) | 151.94 (74.17-298.64) | 168.72 (82.61-342.54) | 0.42 (0.36-0.47) |
| Albania | 394.35 (350.82-437.54) | 450.79 (399.77-501.81) | 0.50 (0.48-0.53) | 5015.70 (4461.40-5552.49) | 5802.12 (5134.05-6443.77) | 0.56 (0.53-0.58) | 172.72 (84.18-345.16) | 202.48 (95.80-411.34) | 0.62 (0.59-0.65) |
| Algeria | 423.18 (375.95-471.98) | 497.51 (436.82-552.44) | 0.54 (0.53-0.55) | 5358.53 (4741.54-5992.18) | 6415.25 (5660.49-7119.58) | 0.59 (0.58-0.61) | 183.41 (87.54-373.15) | 221.98 (107.75-449.66) | 0.63 (0.62-0.65) |
| American Samoa | 547.33 (481.28-608.20) | 592.89 (525.32-658.09) | 0.22 (0.16-0.27) | 7179.89 (6341.41-8017.68) | 7748.26 (6881.09-8567.07) | 0.20 (0.14-0.25) | 251.69 (119.41-509.07) | 270.12 (129.45-551.87) | 0.18 (0.12-0.24) |
| Andorra | 511.64 (452.39-572.82) | 551.87 (489.74-614.22) | 0.23 (0.21-0.26) | 6593.29 (5912.65-7304.55) | 7051.36 (6341.99-7816.83) | 0.22 (0.20-0.24) | 233.96 (112.66-475.84) | 251.11 (120.46-499.59) | 0.24 (0.21-0.26) |
| Angola | 467.32 (411.19-518.94) | 512.67 (451.42-570.53) | 0.31 (0.30-0.32) | 6140.72 (5390.56-6872.11) | 6744.43 (5931.12-7515.87) | 0.31 (0.30-0.32) | 212.45 (101.54-425.96) | 234.86 (113.65-474.44) | 0.35 (0.33-0.36) |
| Antigua and Barbuda | 539.12 (479.32-598.36) | 578.69 (509.83-644.12) | 0.22 (0.21-0.23) | 6937.47 (6157.08-7659.58) | 7428.77 (6608.40-8288.91) | 0.21 (0.20-0.22) | 245.09 (115.82-494.51) | 261.86 (125.07-527.03) | 0.20 (0.19-0.22) |
| Argentina | 541.77 (479.37-603.57) | 594.76 (527.85-658.47) | 0.28 (0.25-0.31) | 7040.54 (6291.77-7820.67) | 7662.06 (6875.74-8441.60) | 0.27 (0.24-0.29) | 249.80 (118.69-501.09) | 273.79 (131.17-552.10) | 0.29 (0.26-0.32) |
| Armenia | 417.83 (366.13-467.52) | 498.36 (433.96-561.53) | 0.66 (0.62-0.70) | 5647.29 (4927.09-6375.58) | 6877.82 (5958.90-7835.83) | 0.75 (0.70-0.80) | 196.82 (94.89-394.86) | 243.48 (117.86-489.79) | 0.81 (0.76-0.87) |
| Australia | 554.29 (492.60-615.90) | 619.68 (549.23-687.01) | 0.34 (0.31-0.37) | 7193.32 (6457.14-7943.79) | 7930.63 (7103.71-8774.54) | 0.31 (0.29-0.34) | 254.52 (122.89-514.01) | 283.74 (140.00-579.91) | 0.34 (0.32-0.36) |
| Austria | 512.67 (455.67-567.05) | 547.99 (489.83-610.91) | 0.19 (0.18-0.20) | 6666.34 (5990.86-7355.99) | 7020.47 (6338.21-7745.78) | 0.15 (0.15-0.16) | 235.44 (112.91-471.62) | 250.18 (119.59-502.77) | 0.18 (0.17-0.19) |
| Azerbaijan | 456.98 (400.36-514.64) | 507.78 (443.37-571.04) | 0.42 (0.34-0.50) | 6391.43 (5543.13-7311.22) | 7036.93 (6082.81-7974.31) | 0.40 (0.31-0.50) | 226.04 (108.16-455.57) | 250.24 (121.34-498.42) | 0.43 (0.33-0.54) |
| Bahamas | 560.02 (495.79-620.75) | 593.64 (522.42-658.79) | 0.19 (0.18-0.21) | 7207.99 (6414.18-8012.83) | 7623.76 (6709.03-8430.50) | 0.18 (0.16-0.20) | 256.28 (122.45-515.73) | 270.48 (130.50-543.63) | 0.18 (0.16-0.20) |
| Bahrain | 474.08 (417.34-525.61) | 510.40 (450.11-570.18) | 0.23 (0.23-0.24) | 6245.17 (5519.86-6943.99) | 6726.28 (5940.43-7469.32) | 0.23 (0.23-0.24) | 216.41 (103.00-444.87) | 232.14 (111.32-470.08) | 0.22 (0.22-0.23) |
| Bangladesh | 402.60 (355.11-449.36) | 455.67 (405.41-506.44) | 0.44 (0.40-0.48) | 5088.75 (4524.31-5639.13) | 5823.51 (5178.29-6468.16) | 0.47 (0.43-0.51) | 172.10 (82.68-347.31) | 199.66 (96.24-406.20) | 0.52 (0.47-0.56) |
| Barbados | 555.94 (490.84-620.48) | 595.24 (525.11-661.83) | 0.23 (0.21-0.25) | 7124.99 (6331.32-7932.76) | 7638.19 (6759.38-8450.26) | 0.23 (0.21-0.25) | 252.98 (118.68-511.77) | 271.00 (128.56-550.66) | 0.23 (0.21-0.26) |
| Belarus | 507.03 (447.17-566.78) | 564.41 (497.20-630.19) | 0.40 (0.38-0.41) | 6906.92 (6100.22-7764.96) | 7779.43 (6836.89-8711.71) | 0.44 (0.42-0.46) | 243.01 (115.90-494.74) | 276.64 (131.02-552.93) | 0.49 (0.47-0.52) |
| Belgium | 512.44 (456.01-570.60) | 547.40 (487.41-610.97) | 0.18 (0.16-0.20) | 6669.13 (5978.04-7364.22) | 7020.14 (6333.51-7766.14) | 0.16 (0.14-0.17) | 235.88 (112.56-469.94) | 249.12 (120.13-503.67) | 0.16 (0.15-0.17) |
| Belize | 509.91 (450.86-565.06) | 579.85 (515.88-644.57) | 0.39 (0.35-0.44) | 6432.33 (5709.35-7146.34) | 7355.87 (6544.10-8170.19) | 0.40 (0.35-0.46) | 225.89 (109.24-451.93) | 259.76 (124.56-531.99) | 0.42 (0.36-0.48) |
| Benin | 427.31 (379.13-475.64) | 492.73 (436.01-545.17) | 0.46 (0.44-0.48) | 5318.89 (4714.43-5935.71) | 6253.80 (5563.65-6986.83) | 0.52 (0.50-0.54) | 180.26 (86.82-363.75) | 216.43 (103.74-435.14) | 0.59 (0.57-0.61) |

| | | | | | | | | | |
|---|---|---|---|---|---|---|---|---|---|
| Bermuda | 584.28 (515.14-649.11) | 610.02 (538.06-674.79) | 0.14 (0.12-0.15) | 7528.63 (6700.86-8313.17) | 7848.67 (6950.21-8646.83) | 0.13 (0.12-0.14) | 269.51 (127.75-547.59) | 280.77 (134.04-577.67) | 0.13 (0.12-0.15) |
| Bhutan | 409.45 (363.49-454.69) | 465.74 (411.24-516.82) | 0.43 (0.41-0.44) | 5197.25 (4635.95-5794.22) | 5962.90 (5303.80-6602.66) | 0.45 (0.44-0.47) | 176.72 (85.34-357.62) | 205.30 (99.36-416.97) | 0.50 (0.48-0.51) |
| Bolivia (Plurinational State of) | 491.71 (436.14-549.30) | 555.53 (491.78-616.88) | 0.40 (0.39-0.42) | 6196.19 (5522.68-6909.47) | 7051.95 (6265.19-7828.99) | 0.42 (0.40-0.44) | 214.72 (103.06-434.06) | 246.67 (118.88-499.94) | 0.46 (0.44-0.48) |
| Bosnia and Herzegovina | 425.94 (377.16-473.08) | 492.00 (431.49-548.58) | 0.53 (0.46-0.59) | 5527.35 (4885.85-6162.46) | 6533.06 (5777.64-7326.00) | 0.61 (0.54-0.69) | 190.82 (90.86-382.32) | 228.09 (108.19-460.57) | 0.66 (0.58-0.75) |
| Botswana | 456.76 (405.55-509.55) | 535.62 (473.79-595.78) | 0.49 (0.47-0.51) | 5822.96 (5193.81-6519.56) | 6937.39 (6147.07-7710.58) | 0.54 (0.52-0.56) | 201.46 (96.17-406.88) | 241.11 (116.35-485.70) | 0.56 (0.54-0.58) |
| Brazil | 527.73 (467.49-585.46) | 589.96 (522.15-651.51) | 0.38 (0.37-0.39) | 6602.20 (5862.60-7306.99) | 7433.17 (6589.59-8250.70) | 0.40 (0.39-0.41) | 228.00 (109.00-459.80) | 260.23 (124.97-525.16) | 0.45 (0.44-0.47) |
| Brunei Darussalam | 638.10 (565.55-711.02) | 686.67 (609.40-759.01) | 0.23 (0.21-0.26) | 8218.40 (7341.18-9058.99) | 8815.59 (7892.91-9708.06) | 0.23 (0.22-0.25) | 295.71 (141.47-598.41) | 319.00 (152.75-642.40) | 0.26 (0.24-0.28) |
| Bulgaria | 482.22 (426.11-535.83) | 520.68 (461.73-577.32) | 0.25 (0.24-0.26) | 6386.36 (5607.17-7153.15) | 7016.96 (6194.42-7863.49) | 0.32 (0.30-0.33) | 223.82 (106.25-449.81) | 247.88 (119.89-500.07) | 0.36 (0.34-0.37) |
| Burkina Faso | 399.70 (353.47-447.89) | 434.60 (382.77-485.58) | 0.27 (0.26-0.27) | 4913.09 (4360.56-5506.06) | 5373.97 (4778.24-6014.43) | 0.28 (0.28-0.29) | 165.59 (80.14-334.84) | 183.48 (89.33-369.00) | 0.34 (0.33-0.34) |
| Burundi | 399.82 (353.20-444.94) | 408.81 (361.23-455.05) | 0.09 (0.07-0.10) | 4971.21 (4410.04-5556.21) | 5090.05 (4515.49-5674.70) | 0.09 (0.08-0.11) | 169.22 (82.51-338.55) | 174.11 (84.35-350.30) | 0.12 (0.11-0.14) |
| Cabo Verde | 431.97 (381.53-485.99) | 499.28 (442.48-558.22) | 0.49 (0.49-0.50) | 5361.85 (4732.88-6021.12) | 6384.63 (5666.94-7115.52) | 0.60 (0.59-0.61) | 184.22 (89.06-371.37) | 222.28 (106.71-449.99) | 0.65 (0.64-0.66) |
| Cambodia | 349.23 (307.33-389.74) | 395.30 (350.24-440.74) | 0.44 (0.42-0.46) | 4390.22 (3900.72-4913.70) | 5073.17 (4526.94-5661.49) | 0.51 (0.49-0.53) | 147.32 (71.44-297.48) | 172.45 (82.59-347.10) | 0.56 (0.54-0.58) |
| Cameroon | 448.31 (396.44-501.41) | 497.96 (442.52-554.89) | 0.31 (0.29-0.33) | 5590.59 (4980.35-6247.19) | 6290.74 (5604.23-6947.52) | 0.35 (0.32-0.37) | 190.37 (92.14-383.34) | 217.40 (105.19-439.99) | 0.40 (0.37-0.43) |
| Canada | 416.01 (365.47-464.62) | 460.07 (405.52-513.81) | 0.21 (0.15-0.27) | 5726.13 (5028.54-6431.15) | 6228.33 (5509.32-6983.19) | 0.20 (0.16-0.24) | 200.58 (97.21-409.53) | 219.12 (105.39-441.98) | 0.20 (0.16-0.25) |
| Central African Republic | 414.92 (365.64-461.84) | 429.06 (378.82-478.07) | 0.10 (0.09-0.10) | 5234.28 (4629.39-5830.79) | 5416.43 (4810.45-6077.85) | 0.09 (0.08-0.10) | 176.32 (84.74-356.63) | 183.52 (89.17-370.77) | 0.12 (0.11-0.13) |
| Chad | 399.77 (352.55-447.53) | 420.66 (371.48-469.65) | 0.15 (0.14-0.16) | 4928.72 (4386.60-5501.04) | 5195.54 (4610.45-5833.55) | 0.15 (0.15-0.16) | 166.59 (80.97-337.15) | 176.36 (85.79-356.03) | 0.17 (0.16-0.18) |
| Chile | 535.97 (476.19-595.99) | 599.63 (536.35-665.62) | 0.33 (0.30-0.36) | 6879.89 (6154.64-7626.43) | 7686.10 (6940.84-8490.80) | 0.34 (0.31-0.37) | 242.02 (116.91-492.11) | 273.01 (132.17-543.92) | 0.37 (0.34-0.41) |
| China | 487.11 (428.13-543.75) | 554.61 (486.85-619.54) | 0.58 (0.51-0.66) | 6148.92 (5417.29-6855.85) | 7030.66 (6211.20-7831.69) | 0.61 (0.53-0.70) | 210.61 (101.91-423.86) | 244.79 (117.30-491.91) | 0.67 (0.58-0.76) |
| Colombia | 506.09 (447.92-565.04) | 565.49 (498.00-628.15) | 0.38 (0.37-0.39) | 6431.68 (5691.26-7130.27) | 7248.39 (6425.18-8035.66) | 0.41 (0.40-0.42) | 223.91 (106.60-450.01) | 255.85 (121.38-520.40) | 0.47 (0.45-0.48) |
| Comoros | 410.18 (362.05-460.02) | 453.26 (401.04-504.17) | 0.34 (0.33-0.36) | 5099.13 (4529.15-5712.76) | 5730.41 (5090.90-6425.52) | 0.40 (0.39-0.42) | 173.75 (83.81-351.33) | 197.51 (95.79-401.89) | 0.46 (0.43-0.48) |
| Congo | 465.42 (408.90-518.48) | 507.02 (449.56-563.60) | 0.27 (0.25-0.29) | 6008.24 (5308.97-6688.82) | 6579.57 (5822.29-7269.41) | 0.28 (0.26-0.30) | 207.24 (99.02-419.17) | 228.94 (108.50-462.26) | 0.31 (0.29-0.34) |
| Cook Islands | 524.97 (466.34-583.31) | 600.06 (530.08-669.97) | 0.40 (0.36-0.44) | 6847.30 (6086.24-7589.92) | 7820.68 (6948.49-8635.62) | 0.39 (0.35-0.44) | 239.21 (115.12-469.67) | 274.29 (131.43-553.03) | 0.41 (0.37-0.45) |
| Costa Rica | 515.07 (456.42-573.76) | 572.96 (507.23-639.61) | 0.35 (0.33-0.36) | 6530.05 (5782.33-7255.26) | 7319.23 (6495.11-8094.93) | 0.37 (0.35-0.38) | 228.10 (109.03-460.02) | 257.22 (124.02-519.16) | 0.39 (0.38-0.41) |
| Croatia | 475.78 (421.03-529.37) | 514.63 (454.92-573.20) | 0.31 (0.29-0.32) | 6364.06 (5617.23-7127.73) | 6889.04 (6077.98-7714.01) | 0.31 (0.29-0.33) | 222.49 (105.85-451.18) | 242.83 (116.19-490.50) | 0.35 (0.33-0.37) |

| | | | | | | | | | |
|---|---|---|---|---|---|---|---|---|---|
| Cuba | 511.30 (452.56-569.57) | 561.70 (497.53-626.00) | 0.33 (0.32-0.35) | 6460.64 (5711.96-7181.60) | 7149.55 (6341.91-7924.77) | 0.35 (0.34-0.36) | 226.65 (108.22-458.52) | 252.13 (119.70-508.83) | 0.38 (0.36-0.39) |
| Cyprus | 489.24 (435.53-543.47) | 540.01 (480.84-598.39) | 0.31 (0.29-0.33) | 6280.38 (5647.32-6960.81) | 6887.52 (6177.64-7614.12) | 0.31 (0.28-0.34) | 220.50 (106.03-442.22) | 244.83 (118.54-489.74) | 0.35 (0.32-0.38) |
| Czechia | 497.57 (437.85-559.98) | 530.32 (467.70-594.32) | 0.20 (0.19-0.22) | 6729.07 (5936.28-7547.49) | 7158.00 (6278.52-8069.96) | 0.19 (0.17-0.21) | 234.93 (113.65-476.23) | 252.38 (122.37-511.39) | 0.22 (0.20-0.24) |
| Côte d'Ivoire | 430.14 (380.01-479.29) | 473.79 (419.28-528.18) | 0.29 (0.27-0.30) | 5375.53 (4761.31-5987.44) | 5989.82 (5296.36-6648.85) | 0.32 (0.30-0.33) | 180.99 (87.66-366.64) | 206.55 (98.96-421.13) | 0.39 (0.37-0.41) |
| Democratic People's Republic of Korea | 472.99 (414.87-528.27) | 505.10 (444.93-562.60) | 0.23 (0.21-0.24) | 6082.82 (5405.92-6773.87) | 6522.42 (5756.49-7259.56) | 0.23 (0.21-0.25) | 209.30 (99.57-421.76) | 226.01 (109.70-457.60) | 0.26 (0.24-0.28) |
| Democratic Republic of the Congo | 435.39 (383.37-484.02) | 442.97 (392.55-492.31) | -0.00 (-0.07-0.06) | 5495.06 (4869.45-6140.49) | 5630.43 (4986.72-6263.44) | 0.00 (-0.07-0.07) | 186.03 (88.98-378.98) | 192.57 (91.26-388.56) | 0.04 (-0.05-0.12) |
| Denmark | 542.83 (481.96-601.04) | 539.24 (479.51-598.50) | 0.02 (-0.01-0.05) | 7157.23 (6383.36-7947.87) | 6913.85 (6189.62-7612.05) | -0.03 (-0.07-0.01) | 256.17 (125.23-525.24) | 246.85 (119.06-502.47) | -0.02 (-0.07-0.02) |
| Djibouti | 406.30 (358.06-454.02) | 475.16 (421.53-528.18) | 0.58 (0.54-0.62) | 5098.63 (4541.04-5685.89) | 6078.33 (5360.14-6782.81) | 0.65 (0.61-0.70) | 174.85 (84.17-352.08) | 211.85 (101.25-426.85) | 0.72 (0.67-0.77) |
| Dominica | 516.38 (456.62-573.82) | 560.33 (497.81-619.90) | 0.26 (0.24-0.28) | 6535.93 (5798.18-7263.44) | 7159.03 (6343.23-7959.96) | 0.29 (0.26-0.32) | 228.88 (109.84-461.68) | 251.66 (120.62-504.77) | 0.30 (0.27-0.34) |
| Dominican Republic | 513.72 (455.30-568.88) | 569.34 (503.36-635.73) | 0.36 (0.34-0.37) | 6527.81 (5770.04-7251.73) | 7275.90 (6455.96-8027.51) | 0.37 (0.36-0.38) | 229.61 (108.88-460.32) | 256.67 (123.39-517.29) | 0.38 (0.37-0.40) |
| Ecuador | 533.99 (474.80-593.78) | 590.72 (522.18-656.04) | 0.35 (0.34-0.37) | 6764.78 (6025.12-7470.54) | 7519.98 (6667.34-8311.41) | 0.36 (0.34-0.37) | 238.23 (113.46-473.99) | 266.02 (127.18-531.31) | 0.38 (0.36-0.40) |
| Egypt | 426.67 (376.10-479.06) | 485.92 (428.33-542.21) | 0.35 (0.31-0.38) | 5381.83 (4766.07-5997.61) | 6181.47 (5514.48-6898.66) | 0.36 (0.33-0.40) | 184.34 (88.76-376.30) | 212.88 (102.71-441.12) | 0.38 (0.34-0.41) |
| El Salvador | 506.49 (446.95-560.38) | 570.47 (505.56-631.65) | 0.40 (0.37-0.43) | 6409.22 (5654.70-7112.05) | 7260.22 (6468.35-8001.84) | 0.42 (0.39-0.45) | 222.83 (105.15-450.96) | 254.69 (121.81-512.17) | 0.45 (0.42-0.48) |
| Equatorial Guinea | 404.63 (357.48-451.47) | 525.10 (463.69-583.57) | 1.02 (0.95-1.08) | 5036.52 (4473.47-5638.07) | 6803.41 (6010.86-7551.62) | 1.16 (1.09-1.24) | 168.70 (82.70-341.89) | 236.45 (113.18-474.06) | 1.31 (1.23-1.40) |
| Eritrea | 387.17 (342.83-433.15) | 424.42 (373.65-470.44) | 0.30 (0.29-0.31) | 4815.44 (4277.17-5363.61) | 5346.09 (4733.62-5981.62) | 0.34 (0.33-0.36) | 162.80 (77.37-326.36) | 181.97 (87.13-363.62) | 0.37 (0.36-0.38) |
| Estonia | 526.34 (465.97-586.94) | 582.92 (509.57-651.05) | 0.35 (0.33-0.37) | 7234.71 (6372.16-8155.57) | 8078.01 (7084.89-9172.98) | 0.37 (0.34-0.40) | 255.17 (122.08-509.70) | 288.85 (137.70-578.21) | 0.43 (0.39-0.46) |
| Eswatini | 468.62 (414.64-521.03) | 533.06 (470.10-591.84) | 0.37 (0.32-0.42) | 5923.74 (5253.77-6600.06) | 6853.44 (6098.84-7609.43) | 0.43 (0.38-0.47) | 205.20 (97.27-414.23) | 236.74 (114.31-476.35) | 0.41 (0.36-0.47) |
| Ethiopia | 416.45 (368.14-462.50) | 497.67 (438.54-554.72) | 0.66 (0.63-0.69) | 5154.73 (4583.26-5784.53) | 6355.21 (5595.77-7115.11) | 0.78 (0.74-0.82) | 174.24 (83.89-354.23) | 221.13 (106.00-443.57) | 0.90 (0.86-0.94) |
| Fiji | 489.39 (434.75-547.82) | 561.84 (494.76-627.97) | 0.43 (0.39-0.46) | 6324.64 (5621.26-7061.38) | 7350.01 (6534.68-8144.31) | 0.46 (0.42-0.50) | 218.16 (103.81-436.67) | 255.44 (124.01-518.35) | 0.49 (0.45-0.53) |
| Finland | 512.46 (456.36-570.22) | 549.98 (493.38-610.30) | 0.22 (0.20-0.24) | 6653.50 (5991.12-7344.39) | 7049.95 (6353.47-7847.64) | 0.19 (0.17-0.21) | 234.36 (113.87-474.56) | 250.40 (120.14-504.88) | 0.22 (0.20-0.23) |
| France | 511.42 (453.51-571.99) | 547.52 (485.99-604.87) | 0.20 (0.17-0.23) | 6642.77 (5964.33-7337.27) | 7026.03 (6328.47-7763.58) | 0.18 (0.15-0.21) | 234.40 (112.98-472.93) | 249.80 (120.05-501.61) | 0.20 (0.17-0.23) |
| Gabon | 449.43 (394.82-501.74) | 521.52 (462.10-581.15) | 0.47 (0.45-0.48) | 5726.79 (5087.22-6371.20) | 6715.95 (6006.05-7495.81) | 0.49 (0.47-0.51) | 196.73 (94.44-402.34) | 233.49 (111.49-471.87) | 0.53 (0.51-0.56) |
| Gambia | 426.60 (376.61-478.64) | 491.19 (432.75-544.29) | 0.46 (0.45-0.47) | 5354.08 (4758.28-5999.44) | 6266.31 (5546.61-6941.78) | 0.51 (0.50-0.53) | 183.23 (88.19-367.49) | 216.97 (104.66-440.43) | 0.55 (0.54-0.57) |
| Georgia | 446.26 (394.53-501.00) | 479.34 (420.92-539.34) | 0.20 (0.15-0.25) | 6085.05 (5310.80-6875.33) | 6585.18 (5732.69-7446.44) | 0.21 (0.14-0.28) | 214.36 (103.98-429.72) | 231.67 (111.79-465.13) | 0.21 (0.13-0.28) |

| | | | | | | | | | |
|---|---|---|---|---|---|---|---|---|---|
| Germany | 519.99 (464.38-575.95) | 553.67 (490.76-616.73) | 0.20 (0.15-0.25) | 6788.09 (6108.46-7478.47) | 7101.48 (6375.14-7854.64) | 0.21 (0.14-0.28) | 240.34 (116.58-486.29) | 252.42 (124.23-508.18) | 0.21 (0.13-0.28) |
| Ghana | 483.22 (426.33-536.33) | 514.65 (452.38-573.11) | 0.15 (0.13-0.17) | 6293.92 (5570.49-7038.64) | 6596.62 (5839.40-7323.09) | 0.11 (0.09-0.13) | 219.37 (104.44-442.96) | 230.94 (110.66-463.47) | 0.11 (0.09-0.13) |
| Greece | 493.03 (440.17-550.49) | 538.05 (478.80-598.85) | 0.24 (0.16-0.33) | 6273.43 (5658.99-6962.72) | 6841.38 (6128.06-7558.30) | 0.21 (0.09-0.32) | 220.27 (106.68-442.47) | 242.69 (117.51-488.87) | 0.23 (0.10-0.35) |
| Greenland | 421.38 (372.78-472.42) | 471.16 (416.96-527.70) | 0.52 (0.41-0.63) | 5758.74 (5072.46-6443.67) | 6311.89 (5577.44-7018.56) | 0.60 (0.47-0.74) | 199.70 (96.66-407.62) | 221.68 (106.58-445.91) | 0.70 (0.54-0.85) |
| Grenada | 507.06 (448.94-563.76) | 559.45 (496.57-623.76) | 0.35 (0.33-0.37) | 6439.74 (5694.45-7173.18) | 7182.67 (6385.46-7940.49) | 0.30 (0.28-0.32) | 225.19 (108.01-454.13) | 252.03 (120.04-508.96) | 0.35 (0.32-0.38) |
| Guam | 526.14 (459.96-585.54) | 585.14 (516.21-653.49) | 0.31 (0.28-0.33) | 6899.48 (6103.13-7697.34) | 7662.81 (6785.00-8492.58) | 0.34 (0.31-0.38) | 243.66 (117.19-490.61) | 272.57 (130.83-543.30) | 0.35 (0.31-0.39) |
| Guatemala | 473.66 (418.41-526.38) | 526.84 (464.90-587.70) | 0.35 (0.32-0.37) | 5948.62 (5272.25-6601.35) | 6651.75 (5895.61-7375.18) | 0.34 (0.32-0.37) | 204.21 (97.43-411.01) | 229.35 (110.64-468.92) | 0.37 (0.34-0.40) |
| Guinea | 407.52 (360.24-458.15) | 441.35 (391.03-490.61) | 0.34 (0.33-0.35) | 5023.77 (4471.81-5601.18) | 5472.85 (4862.60-6116.27) | 0.35 (0.34-0.36) | 170.35 (82.60-346.01) | 187.16 (90.56-375.05) | 0.37 (0.36-0.37) |
| Guinea-Bissau | 415.91 (366.18-466.96) | 449.91 (396.64-504.30) | 0.23 (0.22-0.25) | 5149.53 (4564.94-5748.58) | 5632.25 (4996.83-6286.28) | 0.25 (0.24-0.26) | 174.40 (85.58-351.70) | 192.95 (92.62-388.98) | 0.28 (0.27-0.29) |
| Guyana | 502.74 (444.60-559.47) | 559.63 (492.67-621.86) | 0.23 (0.22-0.24) | 6351.55 (5626.34-7059.22) | 7142.27 (6320.33-7924.32) | 0.27 (0.25-0.28) | 218.60 (105.30-444.74) | 247.38 (118.93-502.57) | 0.30 (0.29-0.31) |
| Haiti | 442.36 (391.72-492.75) | 482.72 (427.11-537.44) | 0.35 (0.33-0.37) | 5470.04 (4873.16-6126.75) | 6017.15 (5337.40-6713.95) | 0.38 (0.36-0.40) | 185.89 (90.75-375.68) | 205.56 (99.61-414.73) | 0.41 (0.38-0.43) |
| Honduras | 483.21 (423.44-537.00) | 534.58 (472.02-594.80) | 0.31 (0.30-0.32) | 6085.80 (5355.05-6786.11) | 6762.81 (5990.99-7500.81) | 0.34( 0.33-0.35) | 210.73 (101.67-419.56) | 234.49 (113.62-471.52) | 0.36 (0.35-0.37) |
| Hungary | 500.34 (443.39-557.91) | 534.71 (469.04-597.53) | 0.33 (0.33-0.34) | 6709.10 (5925.46-7503.83) | 7214.44 (6368.95-8126.69) | 0.35 (0.34-0.35) | 233.81 (112.60-473.37) | 255.43 (121.98-513.41) | 0.36 (0.35-0.37) |
| Iceland | 549.74 (490.87-609.42) | 570.31 (509.81-631.84) | 0.19 (0.17-0.22) | 7152.25 (6408.10-7939.30) | 7403.16 (6672.74-8204.43) | 0.21 (0.17-0.24) | 257.13 (125.65-529.60) | 266.59 (129.22-534.84) | 0.26 (0.23-0.30) |
| India | 438.57 (389.25-485.34) | 505.00 (445.85-559.19) | 0.08 (0.03-0.13) | 5507.82 (4893.00-6104.55) | 6450.10 (5727.52-7137.44) | 0.11 (0.06-0.15) | 185.04 (89.55-374.84) | 221.21 (106.34-446.55) | 0.11 (0.05-0.16) |
| Indonesia | 383.12 (337.78-426.13) | 446.32 (393.96-496.65) | 0.47 (0.45-0.50) | 4870.66 (4310.22-5475.32) | 5753.51 (5067.26-6426.03) | 0.54(0.51-0.5 7) | 166.18 (79.41-335.23) | 199.35 (95.36-399.40) | 0.62 (0.59-0.64) |
| Iran (Islamic Republic of) | 431.91 (381.86-478.32) | 489.96 (433.93-541.89) | 0.51 (0.49-0.53) | 5447.47 (4832.39-6098.90) | 6230.63 (5520.28-6928.61) | 0.54 (0.52-0.56) | 186.61 (89.16-376.72) | 215.06 (102.51-433.50) | 0.60 (0.57-0.62) |
| Iraq | 448.04 (392.76-498.76) | 482.17 (424.64-537.96) | 0.42 (0.37-0.47) | 5726.27 (5061.17-6360.22) | 6212.41 (5512.95-6899.99) | 0.45 (0.36-0.54) | 196.15 (93.48-394.81) | 211.66 (102.78-427.04) | 0.48 (0.39-0.58) |
| Ireland | 509.37 (451.56-566.39) | 551.97 (492.92-614.47) | 0.21 (0.19-0.23) | 6567.15 (5836.02-7276.77) | 7038.71 (6311.33-7817.10) | 0.22 (0.20-0.25) | 232.12 (111.79-469.46) | 250.99 (121.51-505.28) | 0.21 (0.18-0.23) |
| Israel | 514.10 (457.68-573.02) | 550.06 (491.20-613.26) | 0.23 (0.22-0.25) | 6608.75 (5921.60-7309.56) | 7043.03 (6360.05-7771.73) | 0.22 (0.20-0.24) | 234.39 (111.80-470.27) | 251.49 (119.76-507.17) | 0.24 (0.22-0.27) |
| Italy | 528.28 (469.83-586.66) | 560.17 (498.16-622.36) | -0.19 (-0.37--0.02) | 6763.99 (6076.33-7492.00) | 7102.79 (6365.16-7858.68) | -0.25 (-0.44--0.05) | 238.92 (115.29-479.42) | 253.76 (122.37-511.53) | -0.30 (-0.53--0.08) |
| Jamaica | 512.63 (452.87-570.60) | 560.85 (496.04-622.78) | 0.24 (0.19-0.30) | 6489.84 (5736.57-7193.44) | 7129.51 (6347.43-7865.33) | 0.24 (0.17-0.30) | 228.40 (108.85-461.42) | 251.52 (120.26-507.66) | 0.29 (0.22-0.37) |
| Japan | 633.39 (560.47-699.89) | 671.40 (594.22-740.18) | 0.31 (0.29-0.33) | 7955.13 (7071.66-8796.20) | 8442.68 (7514.97-9306.51) | 0.32 (0.29-0.34) | 287.04 (137.89-578.09) | 309.47 (147.61-625.16) | 0.33 (0.30-0.36) |
| Jordan | 453.18 (403.15-504.64) | 512.39 (455.00-567.56) | 0.40 (0.22-0.57) | 5845.34 (5186.40-6512.19) | 6661.01 (5930.75-7365.31) | 0.45 (0.23-0.67) | 201.61 (97.21-405.82) | 230.62 (110.63-463.10) | 0.56 (0.29-0.83) |

| | | | | | | | | | |
|---|---|---|---|---|---|---|---|---|---|
| Kazakhstan | 468.31 (412.41-523.97) | 545.27 (477.21-610.87) | 0.41 (0.38-0.44) | 6534.70 (5661.37-7420.15) | 7753.16 (6780.81-8862.72) | 0.43 (0.40-0.46) | 229.68 (110.43-463.58) | 275.74 (132.53-557.97) | 0.45 (0.42-0.48) |
| Kenya | 447.59 (396.00-495.11) | 516.33 (457.17-572.70) | 0.52 (0.49-0.55) | 5591.60 (4952.58-6240.34) | 6578.32 (5819.56-7327.82) | 0.59 (0.55-0.63) | 192.09 (92.36-388.17) | 229.67 (109.42-462.80) | 0.64 (0.59-0.68) |
| Kiribati | 499.13 (440.05-557.54) | 542.96 (480.76-604.78) | 0.48 (0.45-0.51) | 6504.82 (5779.53-7252.47) | 7072.38 (6288.68-7879.49) | 0.54 (0.51-0.57) | 223.76 (107.40-446.50) | 245.16 (117.39-491.33) | 0.61 (0.57-0.65) |
| Kuwait | 472.49 (421.41-526.80) | 530.97 (470.38-590.98) | 0.22 (0.17-0.26) | 6195.32 (5519.63-6877.85) | 6867.24 (6095.67-7617.16) | 0.20 (0.15-0.25) | 215.93 (102.79-440.53) | 238.38 (113.72-478.00) | 0.22 (0.17-0.28) |
| Kyrgyzstan | 441.31 (385.52-493.85) | 490.97 (425.08-552.09) | 0.43 (0.41-0.45) | 6139.58 (5334.58-7001.17) | 6870.46 (5921.52-7865.82) | 0.40 (0.38-0.42) | 215.42 (103.79-431.66) | 243.57 (116.39-488.63) | 0.39 (0.37-0.41) |
| Lao People's Democratic Republic | 353.39 (312.05-393.84) | 402.53 (353.19-448.15) | 0.41 (0.35-0.47) | 4473.64 (3984.69-5028.53) | 5173.35 (4569.48-5731.86) | 0.46 (0.38-0.54) | 151.59 (72.58-306.98) | 177.54 (84.62-358.28) | 0.51 (0.42-0.59) |
| Latvia | 518.03 (458.86-574.95) | 574.13 (507.90-640.01) | 0.46 (0.44-0.49) | 7047.79 (6232.73-7902.98) | 7920.03 (6968.25-8955.27) | 0.51 (0.47-0.54) | 247.55 (117.55-502.41) | 282.11 (134.93-576.79) | 0.56 (0.52-0.59) |
| Lebanon | 435.82 (386.32-485.68) | 508.74 (448.00-564.39) | 0.37 (0.35-0.40) | 5584.99 (4968.90-6252.87) | 6601.22 (5883.40-7300.53) | 0.42 (0.39-0.45) | 191.32 (91.40-385.43) | 227.15 (109.45-453.89) | 0.49 (0.45-0.53) |
| Lesotho | 433.46 (383.82-484.21) | 504.29 (447.08-559.17) | 0.50 (0.46-0.53) | 5446.96 (4831.75-6051.35) | 6488.47 (5745.11-7198.35) | 0.54 (0.51-0.57) | 187.32 (90.63-373.79) | 222.40 (106.73-451.47) | 0.56 (0.53-0.59) |
| Liberia | 432.33 (384.14-485.35) | 483.81 (427.55-539.80) | 0.53 (0.51-0.55) | 5391.26 (4783.97-6010.30) | 6111.05 (5430.77-6772.88) | 0.62 (0.59-0.65) | 181.45 (87.44-362.61) | 208.11 (100.37-423.06) | 0.61 (0.58-0.65) |
| Libya | 453.84 (400.43-504.46) | 503.98 (443.81-558.83) | 0.45 (0.41-0.49) | 5860.10 (5206.23-6521.70) | 6530.77 (5787.05-7228.17) | 0.50 (0.45-0.55) | 202.92 (96.32-412.74) | 225.21 (107.82-453.27) | 0.56 (0.50-0.61) |
| Lithuania | 511.24 (452.64-572.02) | 568.44 (499.89-637.71) | 0.35 (0.32-0.38) | 6922.38 (6106.09-7812.01) | 7829.53 (6905.47-8826.83) | 0.36 (0.32-0.39) | 243.17 (116.35-491.92) | 278.30 (132.64-574.62) | 0.34 (0.31-0.38) |
| Luxembourg | 517.72 (462.11-579.82) | 549.70 (491.73-611.82) | 0.40 (0.37-0.42) | 6740.38 (6069.67-7446.97) | 7055.12 (6350.95-7786.37) | 0.46 (0.43-0.50) | 238.81 (115.50-483.36) | 251.93 (122.45-509.09) | 0.51 (0.48-0.55) |
| Madagascar | 387.82 (341.63-435.90) | 408.20 (360.87-454.33) | 0.17 (0.15-0.18) | 4777.37 (4264.66-5337.38) | 5050.98 (4487.25-5638.64) | 0.14 (0.12-0.15) | 161.83 (78.28-325.10) | 172.41 (82.83-346.84) | 0.16 (0.14-0.18) |
| Malawi | 401.64 (356.05-448.51) | 441.17 (389.27-493.50) | 0.18 (0.17-0.19) | 4979.87 (4428.52-5585.09) | 5536.18 (4902.49-6192.45) | 0.19 (0.18-0.21) | 168.71 (81.54-339.58) | 189.73 (91.76-382.09) | 0.23 (0.21-0.24) |
| Malaysia | 411.58 (363.02-457.65) | 473.66 (417.83-527.02) | 0.34 (0.33-0.35) | 5386.56 (4753.29-5990.01) | 6267.07 (5520.08-6965.71) | 0.38 (0.36-0.39) | 185.03 (87.55-370.73) | 217.70 (103.87-443.51) | 0.44 (0.42-0.46) |
| Maldives | 394.84 (347.18-439.80) | 456.17 (400.23-506.73) | 0.47 (0.46-0.49) | 5162.56 (4558.25-5779.79) | 6113.49 (5332.08-6842.20) | 0.49 (0.47-0.52) | 177.55 (84.70-359.45) | 213.79 (101.95-429.88) | 0.54 (0.52-0.56) |
| Mali | 404.78 (356.37-453.36) | 445.03 (394.43-496.85) | 0.49 (0.46-0.53) | 5003.74 (4436.31-5625.95) | 5557.47 (4934.97-6183.51) | 0.55 (0.53-0.58) | 168.89 (80.93-338.21) | 189.59 (91.88-381.30) | 0.61 (0.58-0.65) |
| Malta | 514.89 (455.64-572.87) | 551.31 (491.66-613.98) | 0.31 (0.30-0.31) | 6659.62 (5938.22-7376.78) | 7071.75 (6385.67-7831.83) | 0.34 (0.34-0.35) | 236.77 (115.32-472.40) | 252.86 (122.60-508.25) | 0.39 (0.38-0.39) |
| Marshall Islands | 483.80 (425.40-539.62) | 540.81 (474.60-600.25) | 0.18 (0.15-0.21) | 6338.84 (5581.77-7083.13) | 7072.47 (6217.97-7882.15) | 0.17 (0.14-0.20) | 219.31 (104.67-440.29) | 244.86 (116.04-495.27) | 0.19 (0.15-0.23) |
| Mauritania | 442.99 (390.85-495.64) | 500.54 (441.28-558.33) | 0.33 (0.31-0.36) | 5543.30 (4943.52-6184.44) | 6313.63 (5564.19-6966.39) | 0.32 (0.29-0.35) | 190.19 (92.23-388.04) | 220.12 (105.28-443.81) | 0.32 (0.29-0.35) |
| Mauritius | 426.21 (374.78-474.79) | 483.28 (426.38-540.07) | 0.37 (0.35-0.38) | 5628.46 (4946.82-6267.53) | 6427.34 (5638.19-7185.67) | 0.39 (0.37-0.41) | 193.69 (92.30-391.76) | 222.26 (106.14-445.17) | 0.45 (0.43-0.46) |
| Mexico | 549.16 (485.53-609.06) | 617.03 (544.47-681.41) | 0.43 (0.42-0.44) | 6890.62 (6125.73-7623.91) | 7826.53 (6918.85-8646.82) | 0.44 (0.43-0.46) | 239.45 (114.45-484.81) | 277.12 (132.54-558.87) | 0.46 (0.45-0.48) |
| Micronesia (Federated States of) | 489.52 (430.63-543.96) | 550.86 (485.32-611.78) | 0.41 (0.39-0.43) | 6346.09 (5630.55-7059.52) | 7191.63 (6345.37-7963.08) | 0.46 (0.43-0.49) | 219.37 (104.24-438.86) | 250.51 (119.72-500.88) | 0.52 (0.49-0.55) |

| | | | | | | | | | |
|---|---|---|---|---|---|---|---|---|---|
| Monaco | 536.83 (480.55-597.82) | 566.68 (505.30-629.25) | 0.35 (0.30-0.41) | 6940.07 (6234.74-7689.48) | 7238.13 (6514.11-7963.94) | 0.38 (0.32-0.43) | 248.76 (118.94-503.98) | 259.79 (124.70-520.48) | 0.40 (0.34-0.45) |
| Mongolia | 421.97 (371.58-472.44) | 517.10 (448.90-583.24) | 0.15 (0.14-0.16) | 5744.98 (5009.96-6500.22) | 7361.05 (6383.13-8508.66) | 0.13 (0.12-0.14) | 200.49 (96.11-401.54) | 260.86 (124.73-529.12) | 0.14 (0.12-0.15) |
| Montenegro | 490.10 (430.27-544.71) | 520.39 (455.72-581.45) | 0.69 (0.65-0.73) | 6534.03 (5736.13-7303.23) | 6982.04 (6128.84-7825.05) | 0.87 (0.82-0.92) | 230.26 (110.50-464.37) | 246.54 (117.94-504.22) | 0.93 (0.88-0.98) |
| Morocco | 430.06 (380.30-478.99) | 473.51 (419.47-527.28) | 0.24 (0.23-0.26) | 5503.53 (4899.07-6127.99) | 6061.39 (5386.56-6727.52) | 0.26 (0.25-0.28) | 189.02 (90.88-386.39) | 207.00 (99.31-422.84) | 0.28 (0.26-0.30) |
| Mozambique | 393.94 (348.00-439.98) | 432.72 (382.52-481.34) | 0.28 (0.25-0.31) | 4868.36 (4318.74-5423.74) | 5413.36 (4778.29-6077.61) | 0.27 (0.23-0.31) | 163.40 (78.32-330.18) | 183.15 (87.86-370.97) | 0.25 (0.20-0.29) |
| Myanmar | 358.73 (318.00-397.31) | 423.47 (373.22-470.01) | 0.32 (0.30-0.33) | 4569.29 (4058.26-5124.49) | 5509.96 (4883.59-6154.18) | 0.36 (0.34-0.37) | 154.51 (74.29-312.45) | 189.01 (90.46-376.10) | 0.40 (0.38-0.42) |
| Namibia | 435.73 (384.48-486.90) | 491.31 (434.64-546.38) | 0.61 (0.58-0.64) | 5491.05 (4865.00-6137.22) | 6282.58 (5591.32-6986.71) | 0.68 (0.64-0.71) | 189.21 (92.31-383.13) | 217.66 (104.04-438.91) | 0.74 (0.70-0.78) |
| Nauru | 498.59 (439.16-555.24) | 565.58 (496.77-627.82) | 0.35 (0.33-0.38) | 6469.66 (5732.81-7183.72) | 7401.39 (6526.73-8232.71) | 0.40 (0.37-0.43) | 224.67 (106.75-447.42) | 257.73 (123.00-512.70) | 0.42 (0.39-0.45) |
| Nepal | 387.57 (345.59-431.61) | 446.59 (398.83-496.31) | 0.38 (0.36-0.40) | 4848.20 (4313.74-5394.94) | 5637.45 (5020.51-6276.11) | 0.41 (0.38-0.43) | 161.56 (78.73-329.46) | 190.42 (92.16-385.86) | 0.42 (0.39-0.45) |
| Netherlands | 536.48 (482.43-594.24) | 562.31 (499.26-625.49) | 0.48 (0.45-0.51) | 7007.11 (6413.98-7691.19) | 7179.66 (6487.57-7948.13) | 0.51 (0.48-0.53) | 250.65 (121.91-504.45) | 256.35 (123.40-520.44) | 0.55 (0.52-0.58) |
| New Zealand | 563.05 (500.62-625.21) | 622.04 (553.44-687.72) | 0.06 (-0.01-0.14) | 7201.53 (6453.83-7965.55) | 7851.94 (7027.87-8662.21) | -0.01 (-0.09-0.08) | 254.13 (122.95-512.36) | 281.65 (134.65-568.23) | -0.03 (-0.12-0.07) |
| Nicaragua | 480.78 (422.73-533.68) | 542.53 (479.59-605.28) | 0.31 (0.29-0.33) | 6028.68 (5350.19-6702.72) | 6877.36 (6091.92-7616.93) | 0.28 (0.26-0.30) | 207.89 (100.06-419.25) | 238.86 (116.01-483.03) | 0.33 (0.30-0.35) |
| Niger | 399.91 (353.17-443.53) | 423.84 (375.53-472.84) | 0.40 (0.37-0.42) | 4942.51 (4402.73-5542.87) | 5232.06 (4650.16-5890.50) | 0.43 (0.41-0.45) | 167.32 (81.73-336.66) | 178.35 (86.39-355.00) | 0.45 (0.43-0.48) |
| Nigeria | 450.16 (398.00-500.71) | 498.72 (440.84-553.16) | 0.18 (0.18-0.19) | 5626.71 (4985.17-6276.04) | 6255.95 (5535.47-6973.39) | 0.18 (0.18-0.19) | 192.41 (92.65-388.43) | 217.13 (104.48-439.38) | 0.21 (0.21-0.22) |
| Niue | 520.49 (460.94-581.93) | 584.10 (517.25-647.85) | 0.35 (0.31-0.40) | 6805.20 (6053.21-7651.00) | 7654.17 (6773.95-8485.39) | 0.37 (0.33-0.42) | 237.44 (113.56-477.26) | 268.12 (129.45-536.33) | 0.43 (0.39-0.48) |
| North Macedonia | 453.81 (402.11-505.54) | 496.39 (437.42-552.48) | 0.36 (0.33-0.40) | 5939.21 (5255.63-6635.68) | 6595.38 (5821.31-7362.79) | 0.37 (0.33-0.41) | 206.54 (98.87-419.99) | 230.94 (111.13-468.16) | 0.39 (0.35-0.43) |
| Northern Mariana Islands | 525.90 (465.45-584.25) | 571.41 (503.61-639.83) | 0.33 (0.32-0.35) | 6928.34 (6124.78-7717.16) | 7473.14 (6649.55-8256.84) | 0.39 (0.37-0.41) | 244.05 (117.02-494.33) | 263.59 (124.96-531.34) | 0.43 (0.40-0.45) |
| Norway | 527.78 (468.90-586.59) | 566.43 (504.41-629.34) | 0.21 (0.16-0.26) | 6810.29 (6081.69-7529.30) | 7194.44 (6445.14-7978.71) | 0.19 (0.14-0.24) | 241.23 (116.18-483.89) | 257.93 (124.39-518.57) | 0.19 (0.14-0.24) |
| Oman | 423.69 (377.23-472.04) | 501.17 (444.48-559.73) | 0.28 (0.19-0.37) | 5553.44 (4912.60-6176.06) | 6610.79 (5877.94-7322.45) | 0.26 (0.15-0.37) | 190.09 (91.78-387.39) | 229.48 (109.88-462.56) | 0.32 (0.19-0.45) |
| Pakistan | 394.09 (346.61-441.36) | 457.98 (402.52-509.95) | 0.57 (0.54-0.60) | 4966.42 (4389.77-5580.03) | 5854.83 (5137.27-6550.19) | 0.58 (0.56-0.60) | 168.48 (79.89-341.15) | 200.60 (95.98-403.02) | 0.63 (0.61-0.66) |
| Palau | 523.52 (461.21-583.20) | 577.27 (510.57-644.04) | 0.52 (0.50-0.54) | 6836.86 (6071.43-7611.03) | 7584.40 (6727.15-8413.92) | 0.56 (0.54-0.58) | 238.52 (113.71-486.33) | 264.80 (126.34-530.31) | 0.59 (0.58-0.61) |
| Palestine | 431.54 (383.21-482.87) | 482.51 (423.88-538.85) | 0.29 (0.25-0.32) | 5473.46 (4856.53-6145.15) | 6252.59 (5540.47-6945.76) | 0.30 (0.26-0.34) | 187.46 (90.89-375.80) | 214.84 (102.84-438.13) | 0.30 (0.25-0.34) |
| Panama | 489.36 (432.95-543.53) | 557.46 (495.29-619.08) | 0.33 (0.31-0.36) | 6212.11 (5507.84-6917.34) | 7127.00 (6334.22-7844.65) | 0.40 (0.38-0.43) | 216.38 (103.08-435.86) | 250.60 (120.45-502.55) | 0.42 (0.40-0.44) |
| Papua New Guinea | 418.16 (367.93-466.12) | 458.49 (402.59-512.05) | 0.40 (0.39-0.41) | 5273.87 (4678.32-5859.78) | 5806.13 (5140.09-6474.72) | 0.42 (0.41-0.43) | 178.21 (85.94-357.89) | 197.86 (96.02-398.60) | 0.45 (0.43-0.46) |

| | | | | | | | | | |
|---|---|---|---|---|---|---|---|---|---|
| Paraguay | 523.21 (462.64-581.96) | 552.50 (490.16-612.86) | 0.27 (0.26-0.28) | 6693.27 (5932.58-7381.73) | 7045.67 (6223.98-7826.86) | 0.28 (0.26-0.29) | 234.44 (112.25-472.60) | 246.42 (116.75-501.02) | 0.31 (0.29-0.32) |
| Peru | 520.82 (462.16-579.79) | 578.82 (509.40-641.16) | 0.18 (0.14-0.22) | 6641.17 (5895.34-7382.66) | 7387.15 (6546.68-8191.56) | 0.16 (0.11-0.21) | 233.73 (112.96-473.00) | 262.45 (125.87-531.72) | 0.16 (0.10-0.22) |
| Philippines | 379.14 (333.45-423.79) | 436.09 (382.63-485.96) | 0.34 (0.32-0.36) | 4937.21 (4334.29-5512.24) | 5704.12 (4994.45-6360.81) | 0.33 (0.31-0.36) | 168.30 (79.99-338.38) | 197.17 (93.94-396.21) | 0.36 (0.33-0.39) |
| Poland | 485.83 (428.33-540.86) | 545.91 (480.16-607.04) | 0.42 (0.39-0.46) | 6352.60 (5622.61-7107.47) | 7209.37 (6359.02-8062.52) | 0.42 (0.38-0.45) | 220.78 (106.13-445.93) | 255.44 (122.70-514.99) | 0.46 (0.42-0.50) |
| Portugal | 499.38 (446.73-554.39) | 545.22 (488.28-604.98) | 0.40 (0.38-0.41) | 6419.41 (5732.16-7112.09) | 6967.51 (6290.48-7707.56) | 0.43 (0.41-0.46) | 225.43 (109.11-455.69) | 247.29 (118.83-498.32) | 0.51 (0.48-0.54) |
| Puerto Rico | 579.84 (514.61-646.56) | 625.47 (556.85-693.46) | 0.35 (0.27-0.43) | 7421.24 (6620.21-8213.12) | 8021.69 (7104.50-8862.45) | 0.39 (0.28-0.50) | 263.80 (126.22-528.30) | 285.66 (136.91-575.20) | 0.46 (0.33-0.58) |
| Qatar | 471.63 (416.68-524.49) | 518.18 (460.61-577.47) | 0.27 (0.25-0.29) | 6220.40 (5474.07-6930.89) | 6835.35 (6109.62-7568.07) | 0.28 (0.26-0.30) | 215.51 (103.38-433.91) | 236.31 (114.23-472.87) | 0.29 (0.27-0.31) |
| Republic of Korea | 665.91 (592.27-739.15) | 701.23 (625.36-776.78) | 0.25 (0.23-0.28) | 8639.54 (7715.08-9501.27) | 8997.39 (8082.99-9897.80) | 0.27 (0.24-0.29) | 310.35 (149.89-629.38) | 327.14 (157.44-662.81) | 0.26 (0.24-0.28) |
| Republic of Moldova | 466.87 (413.24-517.77) | 527.51 (467.44-589.45) | 0.26 (0.15-0.37) | 6181.68 (5461.83-6881.08) | 7085.48 (6248.88-7948.01) | 0.23 (0.12-0.34) | 215.04 (102.74-436.54) | 250.31 (119.74-501.88) | 0.28 (0.15-0.40) |
| Romania | 451.25 (400.32-499.77) | 492.13 (434.73-545.08) | 0.50 (0.43-0.56) | 5917.37 (5251.77-6600.81) | 6494.95 (5751.53-7224.19) | 0.56 (0.48-0.64) | 205.84 (98.80-413.85) | 229.11 (108.11-463.15) | 0.62 (0.54-0.71) |
| Russian Federation | 567.41 (498.64-635.22) | 595.22 (523.11-664.62) | 0.32 (0.31-0.34) | 7889.89 (6889.74-8937.70) | 8051.32 (7065.94-9048.10) | 0.35 (0.33-0.37) | 278.77 (132.59-565.82) | 286.24 (137.07-579.34) | 0.41 (0.38-0.43) |
| Rwanda | 393.20 (349.16-439.83) | 426.56 (375.22-474.36) | 0.27 (0.23-0.30) | 4847.03 (4312.01-5421.17) | 5300.37 (4691.85-5937.64) | 0.23 (0.18-0.29) | 164.19 (78.80-330.13) | 181.00 (87.52-364.52) | 0.28 (0.21-0.34) |
| Saint Kitts and Nevis | 542.16 (477.32-601.08) | 588.16 (521.19-651.58) | 0.29 (0.27-0.31) | 6945.60 (6130.20-7686.98) | 7571.27 (6700.23-8384.82) | 0.32 (0.30-0.34) | 244.01 (116.79-490.25) | 267.40 (127.24-537.19) | 0.36 (0.33-0.39) |
| Saint Lucia | 507.97 (448.15-566.44) | 567.98 (502.87-628.95) | 0.26 (0.25-0.28) | 6415.95 (5699.45-7143.84) | 7248.65 (6444.19-8008.88) | 0.28 (0.26-0.29) | 222.62 (107.61-452.77) | 254.17 (122.58-513.27) | 0.30 (0.28-0.31) |
| Saint Vincent and the Grenadines | 504.61 (449.56-563.01) | 558.35 (492.89-624.11) | 0.33 (0.30-0.37) | 6397.95 (5679.70-7065.58) | 7102.72 (6286.60-7848.03) | 0.36 (0.32-0.40) | 223.20 (106.16-449.77) | 248.54 (120.13-511.44) | 0.39 (0.35-0.44) |
| Samoa | 501.83 (442.89-558.99) | 554.50 (488.20-621.23) | 0.35 (0.34-0.37) | 6490.46 (5805.31-7196.06) | 7229.28 (6387.21-8051.03) | 0.36 (0.34-0.38) | 224.83 (107.81-446.44) | 252.00 (121.02-504.65) | 0.37 (0.35-0.39) |
| San Marino | 524.79 (466.35-586.08) | 561.86 (499.88-630.44) | 0.32 (0.30-0.33) | 6783.34 (6108.04-7523.27) | 7145.91 (6446.11-7953.04) | 0.35 (0.34-0.36) | 241.43 (118.01-484.73) | 255.32 (123.31-515.39) | 0.37 (0.36-0.39) |
| Sao Tome and Principe | 449.75 (398.28-501.90) | 524.21 (462.18-582.24) | 0.20 (0.18-0.21) | 5685.90 (5023.11-6379.01) | 6782.54 (5994.58-7537.66) | 0.16 (0.14-0.18) | 196.47 (94.67-403.55) | 238.24 (114.64-481.44) | 0.17 (0.15-0.19) |
| Saudi Arabia | 438.33 (387.05-487.74) | 519.02 (457.66-575.01) | 0.51 (0.49-0.53) | 5667.97 (5009.98-6297.62) | 6748.90 (5982.75-7462.24) | 0.60 (0.57-0.63) | 193.15 (93.26-388.67) | 232.61 (112.24-467.66) | 0.66 (0.63-0.69) |
| Senegal | 433.81 (381.84-484.04) | 475.47 (416.82-528.94) | 0.34 (0.26-0.43) | 5413.82 (4796.04-6047.40) | 6000.67 (5295.74-6683.86) | 0.26 (0.14-0.39) | 184.61 (88.68-373.17) | 206.53 (98.57-415.10) | 0.27 (0.14-0.41) |
| Serbia | 454.10 (402.28-504.77) | 501.27 (442.69-560.42) | 0.26 (0.25-0.28) | 5940.02 (5240.90-6624.06) | 6639.37 (5843.91-7416.34) | 0.29 (0.28-0.31) | 207.20 (98.31-417.56) | 233.45 (111.10-471.88) | 0.32 (0.31-0.34) |
| Seychelles | 425.83 (375.95-474.19) | 484.27 (424.30-538.98) | 0.37 (0.36-0.39) | 5597.53 (4954.42-6239.91) | 6468.72 (5702.10-7176.25) | 0.42 (0.40-0.44) | 194.22 (92.43-387.60) | 224.69 (107.34-452.80) | 0.46 (0.43-0.48) |
| Sierra Leone | 410.11 (361.85-459.47) | 456.66 (405.02-507.81) | 0.41 (0.38-0.45) | 5080.13 (4526.96-5700.05) | 5743.70 (5103.84-6394.00) | 0.45 (0.41-0.49) | 171.91 (83.49-344.78) | 197.17 (94.57-398.34) | 0.46 (0.41-0.50) |
| Singapore | 662.46 (583.53-735.49) | 685.67 (606.53-760.52) | 0.34 (0.32-0.37) | 8553.41 (7625.49-9400.16) | 8795.59 (7834.77-9662.12) | 0.39 (0.36-0.42) | 312.25 (148.94-621.24) | 323.81 (156.07-648.84) | 0.45 (0.41-0.48) |

| | | | | | | | | | |
|---|---|---|---|---|---|---|---|---|---|
| Slovakia | 493.73<br>(434.26-553.74) | 530.37<br>(465.34-589.97) | 0.07<br>(0.06-0.09) | 6633.92<br>(5808.75-7454.60) | 7172.31<br>(6328.32-8022.93) | 0.08<br>(0.07-0.09) | 231.89<br>(109.63-471.96) | 253.76<br>(121.46-514.80) | 0.11<br>(0.10-0.13) |
| Slovenia | 491.11<br>(431.90-546.56) | 525.36<br>(463.12-586.40) | 0.22<br>(0.20-0.24) | 6626.19<br>(5823.73-7430.59) | 7066.98<br>(6214.38-7901.68) | 0.24<br>(0.21-0.26) | 231.24<br>(109.64-465.72) | 249.66<br>(119.93-506.22) | 0.28<br>(0.25-0.30) |
| Solomon Islands | 444.64<br>(392.62-495.53) | 506.17<br>(444.44-563.70) | 0.22<br>(0.20-0.25) | 5668.76<br>(5030.93-6322.85) | 6521.52<br>(5770.18-7279.33) | 0.20<br>(0.18-0.23) | 194.58<br>(93.43-392.85) | 225.65<br>(107.13-452.46) | 0.24<br>(0.21-0.27) |
| Somalia | 397.57<br>(350.12-443.01) | 413.53<br>(365.08-463.06) | 0.40<br>(0.37-0.43) | 4936.42<br>(4380.82-5554.07) | 5154.48<br>(4571.28-5746.80) | 0.43<br>(0.40-0.46) | 167.54<br>(80.77-340.99) | 175.08<br>(84.25-352.26) | 0.45<br>(0.42-0.48) |
| South Africa | 538.52<br>(477.00-598.27) | 579.68<br>(513.41-643.73) | 0.16<br>(0.15-0.17) | 6897.67<br>(6089.39-7678.13) | 7437.78<br>(6576.71-8268.63) | 0.17<br>(0.16-0.19) | 241.82<br>(116.11-486.72) | 259.73<br>(125.61-521.32) | 0.19<br>(0.17-0.20) |
| South Sudan | 391.68<br>(345.93-437.05) | 419.30<br>(370.66-468.51) | 0.26<br>(0.25-0.27) | 4833.03<br>(4315.14-5389.30) | 5173.45<br>(4585.44-5772.62) | 0.28<br>(0.26-0.29) | 161.50<br>(78.48-322.42) | 173.75<br>(83.86-349.85) | 0.27<br>(0.25-0.29) |
| Spain | 509.74<br>(452.16-570.09) | 550.53<br>(490.54-613.91) | 0.24<br>(0.23-0.25) | 6585.06<br>(5902.27-7293.35) | 7067.43<br>(6346.65-7834.40) | 0.25<br>(0.23-0.26) | 233.09<br>(112.14-466.70) | 252.41<br>(121.52-509.81) | 0.27<br>(0.26-0.29) |
| Sri Lanka | 380.91<br>(336.17-423.87) | 438.19<br>(385.54-486.66) | 0.23<br>(0.20-0.26) | 4907.64<br>(4340.57-5473.98) | 5737.33<br>(5047.68-6405.69) | 0.25<br>(0.22-0.29) | 167.17<br>(79.94-337.48) | 196.48<br>(93.15-395.00) | 0.27<br>(0.23-0.31) |
| Sudan | 374.78<br>(330.89-417.35) | 451.05<br>(400.85-502.02) | 0.48<br>(0.46-0.49) | 4649.53<br>(4118.90-5216.84) | 5699.10<br>(5039.27-6361.30) | 0.52<br>(0.51-0.53) | 156.88<br>(75.28-315.18) | 195.16<br>(93.83-392.69) | 0.54<br>(0.52-0.56) |
| Suriname | 539.08<br>(474.72-599.35) | 583.07<br>(515.65-644.18) | 0.62<br>(0.58-0.67) | 6909.37<br>(6124.45-7648.52) | 7492.59<br>(6632.25-8310.76) | 0.69<br>(0.64-0.74) | 243.78<br>(115.72-497.39) | 263.42<br>(126.60-526.28) | 0.74<br>(0.69-0.80) |
| Sweden | 448.03<br>(393.92-504.34) | 490.50<br>(429.31-550.45) | 0.28<br>(0.27-0.29) | 5721.62<br>(5030.11-6419.35) | 6206.46<br>(5449.73-6981.99) | 0.29<br>(0.28-0.30) | 201.04<br>(94.46-412.76) | 219.94<br>(103.56-446.30) | 0.29<br>(0.27-0.30) |
| Switzerland | 512.78<br>(455.29-571.81) | 536.25<br>(478.09-595.83) | 0.38<br>(0.20-0.56) | 6659.33<br>(5952.03-7400.47) | 6869.39<br>(6203.94-7594.36) | 0.40<br>(0.19-0.61) | 234.87<br>(113.98-474.03) | 243.32<br>(117.04-483.61) | 0.44<br>(0.21-0.68) |
| Syrian Arab Republic | 421.30<br>(376.92-467.98) | 482.73<br>(425.82-541.86) | 0.13<br>(0.12-0.13) | 5315.64<br>(4725.16-5952.02) | 6178.65<br>(5461.63-6883.36) | 0.10<br>(0.10-0.10) | 181.91<br>(87.43-364.87) | 212.96<br>(102.28-423.55) | 0.12<br>(0.11-0.12) |
| Taiwan<br>(Province of China) | 519.32<br>(456.78-584.18) | 594.32<br>(526.06-663.52) | 0.46<br>(0.44-0.47) | 6717.37<br>(5931.78-7456.00) | 7761.15<br>(6899.98-8652.98) | 0.52<br>(0.50-0.54) | 234.43<br>(112.04-470.13) | 273.91<br>(129.60-554.09) | 0.55<br>(0.52-0.57) |
| Tajikistan | 398.52<br>(351.00-447.99) | 437.73<br>(382.62-489.19) | 0.50<br>(0.47-0.52) | 5395.27<br>(4710.30-6124.38) | 5919.48<br>(5131.91-6713.73) | 0.52<br>(0.50-0.54) | 188.20<br>(90.38-379.32) | 207.38<br>(99.15-415.74) | 0.56<br>(0.53-0.58) |
| Thailand | 383.46<br>(337.94-423.76) | 462.73<br>(408.91-515.69) | 0.34<br>(0.29-0.40) | 4903.26<br>(4351.89-5480.03) | 5999.99<br>(5325.54-6676.29) | 0.34<br>(0.27-0.41) | 166.64<br>(81.37-337.43) | 207.81<br>(100.02-418.95) | 0.36<br>(0.29-0.43) |
| Timor-Leste | 348.39<br>(306.28-389.21) | 400.38<br>(352.99-446.00) | 0.65<br>(0.63-0.66) | 4416.96<br>(3920.11-4966.90) | 5191.62<br>(4582.52-5781.30) | 0.69<br>(0.68-0.70) | 148.80<br>(71.51-300.75) | 176.95<br>(84.18-358.22) | 0.76<br>(0.74-0.77) |
| Togo | 421.50<br>(372.31-469.40) | 469.20<br>(415.21-524.26) | 0.52<br>(0.49-0.55) | 5239.90<br>(4654.07-5851.10) | 5942.02<br>(5283.59-6623.74) | 0.61<br>(0.57-0.65) | 178.46<br>(85.94-356.45) | 205.46<br>(97.97-413.44) | 0.67<br>(0.62-0.72) |
| Tokelau | 485.33<br>(427.99-541.92) | 561.14<br>(496.02-621.88) | 0.33<br>(0.31-0.35) | 6244.09<br>(5532.85-6958.76) | 7321.90<br>(6519.88-8126.66) | 0.39<br>(0.37-0.41) | 215.95<br>(104.32-435.22) | 255.45<br>(122.74-515.18) | 0.44<br>(0.42-0.47) |
| Tonga | 493.71<br>(435.22-550.03) | 548.75<br>(483.06-609.57) | 0.48<br>(0.45-0.51) | 6346.13<br>(5623.63-7059.81) | 7098.51<br>(6312.37-7929.74) | 0.52<br>(0.49-0.56) | 220.27<br>(105.41-444.73) | 247.51<br>(119.11-497.80) | 0.56<br>(0.53-0.60) |
| Trinidad and Tobago | 542.83<br>(477.85-606.65) | 584.43<br>(517.34-650.64) | 0.28<br>(0.23-0.33) | 6946.98<br>(6166.35-7707.36) | 7493.92<br>(6678.70-8293.18) | 0.29<br>(0.24-0.35) | 243.98<br>(116.21-492.29) | 263.61<br>(127.05-542.33) | 0.31<br>(0.25-0.37) |
| Tunisia | 429.98<br>(379.47-481.71) | 492.79<br>(438.30-545.86) | 0.28<br>(0.26-0.30) | 5476.94<br>(4853.16-6104.75) | 6374.66<br>(5640.98-7079.66) | 0.29<br>(0.27-0.31) | 188.28<br>(89.50-382.15) | 219.92<br>(106.32-446.51) | 0.30<br>(0.27-0.32) |
| Turkmenistan | 431.32<br>(381.15-485.90) | 504.12<br>(440.58-569.61) | 0.45<br>(0.44-0.46) | 5900.62<br>(5140.52-6667.35) | 7040.77<br>(6084.80-8034.71) | 0.50<br>(0.49-0.51) | 207.07<br>(99.30-418.51) | 250.85<br>(118.85-501.52) | 0.52<br>(0.50-0.54) |
| Tuvalu | 483.16<br>(425.55-536.43) | 547.04<br>(481.10-611.49) | 0.56<br>(0.52-0.60) | 6213.43<br>(5507.54-6931.80) | 7164.29<br>(6313.54-8004.47) | 0.63<br>(0.57-0.69) | 215.00<br>(102.66-438.95) | 250.57<br>(118.95-512.59) | 0.69<br>(0.62-0.76) |

| | | | | | | | | | |
|---|---|---|---|---|---|---|---|---|---|
| Türkiye | 432.28<br>(383.55-478.70) | 504.54<br>(447.64-564.47) | 0.37<br>(0.33-0.41) | 5451.00<br>(4840.24-6098.01) | 6494.84<br>(5774.23-7194.61) | 0.43<br>(0.39-0.47) | 186.92<br>(89.85-377.73) | 225.44<br>(108.35-455.74) | 0.48<br>(0.44-0.52) |
| Uganda | 393.38<br>(345.16-440.69) | 432.88<br>(383.31-481.65) | 0.51<br>(0.44-0.57) | 4869.53<br>(4324.75-5462.94) | 5411.13<br>(4799.77-6007.11) | 0.57<br>(0.49-0.65) | 164.44<br>(79.49-331.17) | 185.04<br>(89.61-375.17) | 0.61<br>(0.51-0.71) |
| Ukraine | 523.44<br>(460.03-582.69) | 563.85<br>(495.98-629.08) | 0.32<br>(0.31-0.32) | 6932.92<br>(6103.05-7792.59) | 7552.96<br>(6637.19-8518.02) | 0.35<br>(0.34-0.35) | 243.21<br>(118.00-492.05) | 267.24<br>(127.59-536.33) | 0.40<br>(0.39-0.41) |
| United Arab Emirates | 442.64<br>(390.98-493.56) | 492.29<br>(435.28-547.19) | 0.27<br>(0.25-0.29) | 5833.58<br>(5145.39-6506.84) | 6385.07<br>(5686.29-7057.91) | 0.30<br>(0.28-0.33) | 200.85<br>(95.71-407.28) | 220.60<br>(104.60-444.37) | 0.35<br>(0.33-0.38) |
| United Kingdom | 553.89<br>(494.51-614.65) | 599.09<br>(535.27-664.80) | 0.36<br>(0.34-0.38) | 7062.83<br>(6351.53-7819.47) | 7557.53<br>(6798.39-8354.38) | 0.30<br>(0.27-0.33) | 251.97<br>(121.31-505.51) | 271.03<br>(130.90-542.66) | 0.32<br>(0.28-0.35) |
| United Republic of Tanzania | 425.28<br>(376.47-472.64) | 453.74<br>(402.88-504.99) | 0.15<br>(0.03-0.26) | 5346.65<br>(4755.87-5958.26) | 5701.50<br>(5083.81-6350.48) | 0.11<br>(-0.02-0.24) | 182.42<br>(87.39-365.72) | 196.17<br>(93.77-396.95) | 0.11<br>(-0.04-0.26) |
| United States of America | 626.75<br>(554.47-694.08) | 668.49<br>(591.69-739.57) | 0.13<br>(0.09-0.16) | 8228.07<br>(7415.47-9087.48) | 8686.57<br>(7789.66-9568.34) | 0.10<br>(0.05-0.14) | 295.34<br>(141.51-594.21) | 310.78<br>(149.65-627.25) | 0.12<br>(0.07-0.17) |
| United States Virgin Islands | 569.16<br>(503.11-631.10) | 612.16<br>(539.67-680.71) | 0.06<br>(-0.08-0.21) | 7283.17<br>(6447.04-8083.47) | 7850.62<br>(6982.97-8658.84) | 0.09<br>(-0.05-0.24) | 258.46<br>(124.49-520.07) | 278.10<br>(133.64-553.80) | 0.07<br>(-0.09-0.23) |
| Uruguay | 540.15<br>(478.19-600.17) | 595.28<br>(530.77-661.96) | 0.24<br>(0.22-0.27) | 6998.28<br>(6221.71-7747.47) | 7656.56<br>(6885.19-8513.04) | 0.25<br>(0.22-0.27) | 248.30<br>(118.53-493.38) | 272.99<br>(131.65-540.80) | 0.24<br>(0.21-0.27) |
| Uzbekistan | 435.64<br>(382.79-491.51) | 498.85<br>(434.94-559.33) | 0.29<br>(0.27-0.31) | 6016.32<br>(5236.04-6831.58) | 6911.10<br>(5988.98-7871.82) | 0.29<br>(0.27-0.31) | 211.06<br>(102.14-426.42) | 244.62<br>(116.89-490.38) | 0.31<br>(0.29-0.33) |
| Vanuatu | 429.49<br>(376.82-474.39) | 479.58<br>(425.49-537.88) | 0.43<br>(0.40-0.47) | 5420.28<br>(4798.65-6034.68) | 6093.09<br>(5397.97-6783.92) | 0.43<br>(0.38-0.48) | 184.98<br>(89.11-372.82) | 209.10<br>(101.13-422.71) | 0.46<br>(0.41-0.51) |
| Venezuela (Bolivarian Republic of) | 525.72<br>(464.16-586.56) | 568.58<br>(504.58-632.00) | 0.34<br>(0.33-0.35) | 6720.09<br>(5958.04-7442.24) | 7290.87<br>(6435.35-8058.36) | 0.36<br>(0.35-0.37) | 235.34<br>(113.35-469.68) | 256.79<br>(123.56-519.59) | 0.38<br>(0.37-0.39) |
| Viet Nam | 351.54<br>(310.19-392.73) | 400.84<br>(350.75-445.43) | 0.16<br>(0.10-0.21) | 4453.32<br>(3942.04-4957.73) | 5186.58<br>(4567.32-5777.21) | 0.15<br>(0.09-0.21) | 151.39<br>(73.07-301.99) | 178.97<br>(85.09-356.75) | 0.15<br>(0.08-0.22) |
| Yemen | 363.38<br>(320.07-407.27) | 421.25<br>(371.11-470.28) | 0.47<br>(0.45-0.49) | 4516.35<br>(4006.31-5058.53) | 5330.19<br>(4703.28-5941.51) | 0.54<br>(0.51-0.56) | 151.46<br>(72.95-310.15) | 181.25<br>(87.44-367.28) | 0.59<br>(0.57-0.61) |
| Zambia | 430.51<br>(379.49-480.06) | 453.81<br>(400.90-504.97) | 0.56<br>(0.52-0.59) | 5409.13<br>(4790.74-6032.38) | 5732.05<br>(5074.71-6383.16) | 0.63<br>(0.59-0.67) | 185.17<br>(87.91-375.23) | 196.37<br>(94.63-394.72) | 0.69<br>(0.64-0.73) |
| Zimbabwe | 429.34<br>(381.87-477.08) | 447.86<br>(394.32-502.70) | 0.18<br>(0.14-0.23) | 5372.60<br>(4786.08-6015.11) | 5598.15<br>(4951.88-6252.71) | 0.21<br>(0.15-0.26) | 184.56<br>(89.04-371.24) | 190.98<br>(92.24-381.93) | 0.22<br>(0.16-0.29) |

**Table S6. The top three countries and territories with the burden of osteoarthritis in 2021**

| | Both | Number | Male | Number | Female | Number |
|---|---|---|---|---|---|---|
| Incident | China | 11652721.19 | China | 4671650.58 | China | 6981070.61 |
| | India | 6701764.08 | India | 2602746.96 | India | 4099017.11 |
| | United States of America | 3192701.53 | United States of America | 1326096.12 | United States of America | 1866605.41 |
| Prevalent | China | 152848105.90 | China | 60235814.14 | China | 92612291.79 |
| | India | 79214251.02 | India | 29658088.49 | India | 49556162.53 |
| | United States of America | 47581100.89 | United States of America | 19282206.37 | United States of America | 28298894.52 |
| DALYs | China | 5327389.98 | China | 2094854.20 | China | 3232535.78 |
| | India | 2718457.05 | India | 1004773.08 | India | 1713683.97 |
| | United States of America | 1710760.12 | United States of America | 680232.84 | United States of America | 1030527.28 |
| ASIR | Republic of Korea | 701.23 | United States of America | 577.60 | Republic of Korea | 859.15 |
| | Brunei Darussalam | 686.67 | South Africa | 560.57 | Singapore | 844.82 |
| | Singapore | 685.67 | Mexico | 560.42 | Brunei Darussalam | 842.11 |
| ASPR | Republic of Korea | 8997.39 | United States of America | 7579.65 | Republic of Korea | 10882.76 |
| | Brunei Darussalam | 8815.59 | Mexico | 7208.37 | Singapore | 10714.34 |
| | Singapore | 8795.59 | South Africa | 7124.31 | Brunei Darussalam | 10684.77 |
| ASDR | Republic of Korea | 327.14 | United States of America | 266.63 | Republic of Korea | 402.58 |
| | Singapore | 323.81 | Mexico | 254.51 | Singapore | 401.49 |
| | Brunei Darussalam | 319.00 | Russian Federation | 250.45 | Brunei Darussalam | 393.69 |

**Table S7. The DALYs and ASDR with their variations of osteoarthritis attributable to high BMI at the global, SDI quintile, and regional levels from 1990 to 2021**

| Characteristics | DALYs | | | ASDR (per 100,000 population) | | |
|---|---|---|---|---|---|---|
| | 1990 | 2021 | 1990-2021 | 1990 | 2021 | EAPC (95%CI) |
| | Both (95%UI) | Both (95%UI) | Change (%) | Both (95%UI) | Both (95%UI) | |
| **Global** | 1449681.33 (-127135.50-4113313.24) | 4422953.51 (-421237.19-12338124.92) | 205.10 (191.71-227.09) | 35.97 (-3.14-102.31) | 50.59 (-4.81-141.35) | 1.17 (1.15-1.20) |
| **Sex** | | | | | | |
| Male | 521229.26 (-46292.23-1480937.59) | 1644274.97 (-157014.66-4599839.39) | 215.46 (200.51-236.92) | 27.30 (-2.40-77.90) | 39.47 (-3.75-110.79) | 1.26 (1.24-1.28) |
| Female | 928452.07 (-80843.27-2633520.29) | 2778678.54 (-264222.53-7738285.53) | 199.28 (186.79-223.04) | 43.47 (-3.78-123.36) | 60.74 (-5.78-169.14) | 1.16 (1.13-1.19) |
| **Types** | | | | | | |
| Osteoarthritis knee | 1306589.34 (-114326.79-3742191.00) | 4019554.91 (-382558.22-11222317.96) | 207.64 (194.14-229.71) | 32.35 (-2.81-92.95) | 45.93 (-4.36-128.33) | 0.89 (0.86-0.91) |
| Osteoarthritis hip | 143091.99 (-12808.72-386693.93) | 403398.60 (-38678.96-1076491.22) | 181.92 (169.20-204.89) | 3.63 (-0.32-9.81) | 4.66 (-0.45-12.42) | 1.21 (1.18-1.23) |
| **SDI** | | | | | | |
| High | 569083.03 (-52686.32-1575580.47) | 1271056.92 (-127983.26-3441561.02) | 123.35 (112.98-144.93) | 52.63 (-4.89-145.38) | 66.70 (-6.82-179.94) | 0.71 (0.67-0.76) |
| High-middle | 393635.12 (-35314.98-1102209.19) | 1105143.77 (-108304.99-3016330.75) | 180.75 (168.42-201.99) | 39.00 (-3.48-109.56) | 55.85 (-5.46-152.49) | 1.26 (1.22-1.31) |
| Middle | 318093.44 (-26116.87-922529.50) | 1364967.96 (-126086.15-3863690.88) | 329.11 (302.47-367.69) | 28.73 (-2.33-83.68) | 48.16 (-4.41-137.15) | 1.86 (1.79-1.92) |
| Low-middle | 125910.19 (-10241.84-369632.67) | 527571.30 (-46776.09-1497615.07) | 319.01 (295.13-355.24) | 19.32 (-1.55-57.41) | 34.55 (-3.03-98.53) | 2.02 (1.98-2.06) |
| Low | 41133.19 (-3041.86-123324.51) | 150288.28 (-11967.92-440893.36) | 265.37 (247.47-286.50) | 16.81 (-1.23-51.23) | 26.73 (-2.09-79.82) | 1.55 (1.51-1.59) |
| **Regions** | | | | | | |
| Andean Latin America | 9894.27 (-919.01-27077.60) | 39752.31 (-4229.51-105662.70) | 301.77 (273.28-349.71) | 46.29 (-4.26-127.24) | 65.71 (-6.95-175.47) | 1.18 (1.14-1.21) |
| Australasia | 13064.09 (-1250.84-35716.43) | 39024.36 (-3963.41-105319.17) | 198.71 (172.77-254.30) | 56.54 (-5.43-154.09) | 79.24 (-8.19-213.40) | 1.10 (1.04-1.15) |
| Caribbean | 11257.85 (-978.89-31879.42) | 32228.65 (-3203.64-86894.80) | 186.28 (169.48-220.09) | 42.82 (-3.72-121.23) | 59.71 (-5.93-160.98) | 1.14 (1.10-1.18) |
| Central Asia | 16488.35 (-1541.00-45709.18) | 36183.43 (-3859.63-97115.19) | 119.45 (109.02-140.05) | 34.80 (-3.21-97.26) | 42.04 (-4.37-113.44) | 0.62 (0.61-0.63) |
| Central Europe | 63283.49 (-5875.68-172456.65) | 107282.01 (-11052.48-289260.70) | 69.53 (62.33-86.32) | 42.05 (-3.89-114.92) | 51.25 (-5.32-138.21) | 0.66 (0.64-0.67) |
| Central Latin America | 44279.16 (-4269.90-121092.83) | 174200.55 (-18965.68-458370.89) | 293.41 (269.72-341.58) | 50.85 (-4.85-139.84) | 67.69 (-7.32-178.75) | 0.91 (0.90-0.93) |
| Central Sub-Saharan Africa | 4802.48 (-338.39-14209.67) | 21121.49 (-1648.84-62061.11) | 339.80 (294.33-391.70) | 19.60 (-1.37-58.77) | 34.18 (-2.59-101.87) | 1.78 (1.73-1.83) |

| | | | | | | |
|---|---|---|---|---|---|---|
| East Asia | 256084.43 (-20939.60-755795.37) | 1201663.79 (-108094.93-3403964.66) | 369.25 (326.96-420.35) | 27.46 (-2.23-81.48) | 52.85 (-4.73-150.44) | 2.47 (2.33-2.60) |
| Eastern Europe | 120261.52 (-11249.12-330991.37) | 189331.51 (-19437.22-497720.31) | 57.43 (48.11-78.67) | 42.78 (-3.96-118.19) | 54.95 (-5.69-144.57) | 0.88 (0.85-0.90) |
| Eastern Sub-Saharan Africa | 14443.59 (-1065.74-43728.94) | 54046.78 (-4255.87-157360.22) | 274.19 (254.09-300.65) | 17.90 (-1.31-54.97) | 28.76 (-2.22-84.57) | 1.56 (1.53-1.58) |
| High-income Asia Pacific | 82638.80 (-6623.01-243996.36) | 201743.24 (-15797.04-603491.75) | 144.13 (127.78-163.48) | 39.93 (-3.20-117.82) | 51.50 (-4.14-153.08) | 0.86 (0.84-0.87) |
| High-income North America | 223661.45 (-22013.42-599742.69) | 501346.75 (-55304.04-1308805.36) | 124.15 (108.50-158.65) | 67.01 (-6.64-178.76) | 82.03 (-9.14-215.01) | 0.47 (0.31-0.62) |
| North Africa and Middle East | 72032.85 (-6988.65-198284.45) | 296602.76 (-34260.25-774588.43) | 311.76 (278.37-390.37) | 40.23 (-3.83-111.53) | 59.76 (-6.78-155.92) | 1.28 (1.27-1.29) |
| Oceania | 1315.02 (-122.57-3651.54) | 4313.51 (-449.91-11532.70) | 228.02 (212.02-256.19) | 38.94 (-3.57-109.06) | 48.86 (-4.96-132.98) | 1.28 (1.27-1.29) |
| South Asia | 84239.63 (-6104.07-248489.16) | 439406.19 (-35032.98-1289375.79) | 421.61 (371.26-486.22) | 13.18 (-0.94-39.29) | 27.81 (-2.20-81.78) | 0.69 (0.63-0.76) |
| Southeast Asia | 41752.59 (-3098.97-125493.00) | 201324.62 (-15981.22-582465.03) | 382.18 (352.39-424.84) | 14.76 (-1.09-44.76) | 27.87 (-2.18-80.99) | 2.65 (2.58-2.73) |
| Southern Latin America | 26719.70 (-2619.98-71378.73) | 64937.71 (-7109.23-167296.89) | 143.03 (127.97-174.15) | 57.23 (-5.61-153.03) | 76.31 (-8.39-196.56) | 2.65 (2.58-2.73) |
| Southern Sub-Saharan Africa | 12078.61 (-1082.54-33236.08) | 36137.02 (-3684.36-95528.35) | 199.18 (181.15-238.51) | 43.15 (-3.83-120.08) | 59.34 (-5.99-158.72) | 2.19 (2.13-2.26) |
| Tropical Latin America | 43719.28 (-3904.63-120749.45) | 165101.25 (-16732.60-441786.37) | 277.64 (254.99-318.36) | 45.65 (-4.02-126.66) | 62.95 (-6.35-168.57) | 0.94 (0.88-1.00) |
| Western Europe | 282127.82 (-25876.85-786737.66) | 525194.69 (-51633.50-1415589.91) | 86.15 (78.25-101.81) | 50.71 (-4.69-140.96) | 62.95 (-6.27-168.56) | 1.05 (1.03-1.06) |
| Western Sub-Saharan Africa | 25536.36 (-2047.47-75155.69) | 92010.89 (-8076.14-256558.55) | 260.31 (238.07-296.41) | 27.47 (-2.18-81.74) | 42.20 (-3.60-119.63) | 1.07 (1.04-1.09) |

### Table S8. The DALYs and ASDR with their variations of osteoarthritis attributable to high BMI at the national level from 1990 to 2021

| Characteristics | DALYs | | | ASDR (per 100,000 population) | | |
|---|---|---|---|---|---|---|
| | 1990 | 2021 | 1990-2021 | 1990 | 2021 | EAPC (95%CI) |
| | Both (95%UI) | Both (95%UI) | Change (%) | Both (95%UI) | Both (95%UI) | |
| Afghanistan | 2130.97 (-175.10-6095.98) | 4455.34 (-394.96-12279.37) | 109.08 (91.98-135.95) | 28.75 (-2.32-82.59) | 39.15 (-3.34-111.00) | 1.10 (1.03-1.16) |
| Albania | 816.02 (-79.36-2138.48) | 2048.70 (-217.09-5398.69) | 151.06 (133.22-181.64) | 38.43 (-3.73-101.45) | 47.70 (-5.08-125.80) | 0.73 (0.72-0.74) |
| Algeria | 4566.00 (-412.15-13037.46) | 21403.58 (-2342.76-56691.44) | 368.76 (323.85-468.89) | 35.21 (-3.15-100.90) | 55.71 (-6.01-148.01) | 1.52 (1.49-1.54) |
| American Samoa | 18.09 (-2.16-45.73) | 44.49 (-5.56-110.14) | 145.97 (130.84-181.13) | 70.29 (-8.10-180.51) | 83.99 (-10.25-210.46) | 0.51 (0.42-0.60) |
| Andorra | 28.74 (-2.79-79.59) | 92.04 (-8.51-248.05) | 220.25 (199.62-252.18) | 49.02 (-4.76-135.75) | 60.38 (-5.57-162.63) | 0.70 (0.69-0.71) |
| Angola | 800.45 (-61.27-2336.73) | 4461.97 (-350.93-12872.60) | 457.44 (400.01-540.63) | 18.04 (-1.36-53.49) | 32.43 (-2.46-95.66) | 1.89 (1.85-1.94) |
| Antigua and Barbuda | 23.72 (-2.25-67.41) | 71.60 (-7.78-189.02) | 201.83 (174.34-252.96) | 47.37 (-4.60-132.73) | 63.63 (-6.83-168.87) | 0.97 (0.96-0.98) |
| Argentina | 18350.55 (-1765.46-49359.32) | 41045.05 (-4484.65-106300.00) | 123.67 (107.90-157.17) | 56.27 (-5.42-151.52) | 75.07 (-8.23-193.87) | 0.95 (0.90-1.01) |
| Armenia | 987.65 (-96.70-2701.69) | 1847.18 (-195.30-4961.90) | 87.03 (73.54-107.20) | 35.19 (-3.39-96.97) | 42.95 (-4.55-115.55) | 0.68 (0.66-0.70) |
| Australia | 10901.18 (-1045.32-29843.17) | 33151.02 (-3319.87-88946.40) | 204.10 (175.56-267.04) | 56.56 (-5.43-154.34) | 80.28 (-8.21-214.46) | 1.13 (1.07-1.19) |
| Austria | 5648.57 (-489.96-15739.02) | 10064.51 (-919.72-27954.21) | 78.18 (66.71-97.29) | 49.91 (-4.39-137.30) | 60.54 (-5.61-166.71) | 0.60 (0.59-0.61) |
| Azerbaijan | 1793.79 (-161.98-4903.66) | 4876.06 (-515.87-12868.82) | 171.83 (153.12-209.77) | 34.99 (-3.08-95.83) | 43.98 (-4.48-117.43) | 0.80 (0.77-0.82) |
| Bahamas | 87.62 (-8.05-240.61) | 308.57 (-34.42-798.63) | 252.16 (220.90-312.95) | 54.78 (-4.98-151.63) | 70.88 (-7.75-185.39) | 0.84 (0.80-0.87) |
| Bahrain | 99.48 (-9.83-267.65) | 823.85 (-103.79-2093.89) | 728.12 (644.93-939.90) | 46.57 (-4.48-126.42) | 66.16 (-8.00-168.90) | 1.14 (1.13-1.15) |
| Bangladesh | 5110.29 (-377.68-14944.75) | 34855.05 (-2721.73-102652.61) | 582.06 (483.93-700.58) | 10.05 (-0.74-29.48) | 23.64 (-1.84-70.03) | 3.11 (2.99-3.23) |
| Barbados | 146.62 (-13.90-412.30) | 359.60 (-36.11-935.65) | 145.26 (125.12-184.24) | 55.40 (-5.48-153.48) | 71.79 (-7.23-187.12) | 0.82 (0.79-0.86) |
| Belarus | 5541.31 (-519.62-15528.75) | 8698.02 (-958.86-22790.91) | 56.97 (46.01-80.96) | 42.48 (-3.98-119.17) | 55.08 (-6.08-144.34) | 0.89 (0.87-0.90) |
| Belgium | 6772.31 (-568.01-19070.58) | 12498.05 (-1195.18-34371.14) | 84.55 (72.63-108.06) | 45.89 (-3.83-128.60) | 59.61 (-5.75-162.85) | 0.81 (0.77-0.85) |
| Belize | 51.77 (-4.93-142.85) | 232.80 (-24.29-615.75) | 349.69 (314.22-417.13) | 55.50 (-5.28-152.92) | 72.78 (-7.52-194.91) | 0.86 (0.77-0.95) |
| Benin | 636.98 (-52.15-1817.05) | 2538.22 (-211.39-7154.53) | 298.48 (263.34-341.87) | 30.78 (-2.51-88.62) | 44.09 (-3.59-126.02) | 1.14 (1.11-1.16) |
| Bermuda | 39.55 (-3.73-106.09) | 99.49 (-11.61-260.34) | 151.55 (130.12-197.76) | 62.21 (-5.84-167.13) | 78.82 (-9.26-205.74) | 0.77 (0.73-0.80) |

| | | | | | | |
|---|---|---|---|---|---|---|
| Bhutan | 84.96<br>(-7.58-235.01) | 286.03<br>(-26.00-786.12) | 236.66<br>(208.18-282.80) | 30.31<br>(-2.69-85.60) | 44.53<br>(-4.03-122.85) | 1.34<br>(1.29-1.38) |
| Bolivia<br>(Plurinational State of) | 1392.69<br>(-120.79-3877.32) | 5819.50<br>(-593.58-15670.71) | 317.86<br>(282.04-378.71) | 40.96<br>(-3.51-114.61) | 60.37<br>(-6.07-164.22) | 1.27<br>(1.23-1.32) |
| Bosnia and Herzegovina | 1619.57<br>(-156.23-4438.67) | 2850.34<br>(-302.98-7721.51) | 75.99<br>(64.30-99.02) | 37.26<br>(-3.46-103.19) | 47.54<br>(-5.03-128.52) | 0.79<br>(0.73-0.86) |
| Botswana | 173.61<br>(-13.93-467.33) | 832.07<br>(-69.66-2388.38) | 379.26<br>(329.42-470.16) | 29.44<br>(-2.33-79.57) | 51.80<br>(-4.23-148.70) | 1.82<br>(1.80-1.84) |
| Brazil | 42617.54<br>(-3797.36-117709.26) | 161305.28<br>(-16311.35-431405.06) | 278.50<br>(255.70-319.57) | 45.58<br>(-4.00-126.48) | 62.95<br>(-6.34-168.46) | 1.07<br>(1.05-1.09) |
| Brunei Darussalam | 47.79<br>(-3.86-140.91) | 274.28<br>(-25.24-773.32) | 473.95<br>(410.19-562.16) | 40.74<br>(-3.23-120.36) | 66.30<br>(-5.99-188.80) | 1.64<br>(1.53-1.75) |
| Bulgaria | 5463.24<br>(-533.00-15202.45) | 6689.54<br>(-666.85-18390.73) | 22.45<br>(14.52-36.43) | 43.71<br>(-4.26-121.77) | 49.99<br>(-4.98-136.50) | 0.43<br>(0.41-0.44) |
| Burkina Faso | 766.06<br>(-65.66-2259.30) | 2453.58<br>(-197.56-7176.78) | 220.28<br>(182.25-271.56) | 16.33<br>(-1.38-48.33) | 23.64<br>(-1.89-70.56) | 1.22<br>(1.18-1.26) |
| Burundi | 324.65<br>(-22.88-1011.01) | 1005.15<br>(-74.02-3093.63) | 209.61<br>(167.62-254.19) | 13.51<br>(-0.94-42.35) | 18.51<br>(-1.33-56.98) | 0.97<br>(0.90-1.05) |
| Cabo Verde | 66.37<br>(-5.29-199.93) | 230.89<br>(-21.53-647.25) | 247.87<br>(215.88-306.99) | 30.94<br>(-2.49-92.56) | 48.91<br>(-4.45-138.09) | 1.49<br>(1.48-1.50) |
| Cambodia | 622.91<br>(-48.01-1834.85) | 2703.40<br>(-205.30-8050.53) | 334.00<br>(278.58-410.88) | 12.38<br>(-0.95-37.11) | 19.50<br>(-1.47-58.49) | 1.55<br>(1.51-1.58) |
| Cameroon | 2035.92<br>(-175.95-5695.76) | 8155.30<br>(-870.37-21657.45) | 300.57<br>(268.91-355.68) | 42.23<br>(-3.60-119.26) | 58.54<br>(-6.13-157.26) | 1.03<br>(1.02-1.04) |
| Canada | 11096.33<br>(-1066.23-30208.48) | 30261.11<br>(-2877.28-81528.45) | 172.71<br>(154.86-203.77) | 34.76<br>(-3.34-94.52) | 45.56<br>(-4.41-122.58) | 0.69<br>(0.59-0.78) |
| Central African Republic | 209.55<br>(-15.02-644.48) | 719.38<br>(-55.25-2019.96) | 243.30<br>(202.22-303.30) | 16.30<br>(-1.15-50.78) | 27.43<br>(-2.02-78.12) | 1.73<br>(1.69-1.77) |
| Chad | 622.88<br>(-47.20-1939.39) | 1786.01<br>(-139.38-5064.02) | 186.73<br>(153.29-226.08) | 21.71<br>(-1.63-67.40) | 28.09<br>(-2.17-81.05) | 0.79<br>(0.76-0.82) |
| Chile | 6252.51<br>(-653.28-16742.02) | 20214.92<br>(-2263.82-52058.97) | 223.31<br>(201.85-266.44) | 60.83<br>(-6.32-163.66) | 79.70<br>(-8.95-205.68) | 0.85<br>(0.79-0.91) |
| China | 246468.73<br>(-20130.69-727780.59) | 1164587.77<br>(-104718.16-3297087.97) | 372.51<br>(329.77-424.77) | 27.43<br>(-2.22-81.50) | 52.98<br>(-4.73-150.74) | 2.48<br>(2.35-2.62) |
| Colombia | 8087.35<br>(-713.27-22436.22) | 34990.95<br>(-3654.66-94028.77) | 332.66<br>(295.84-397.15) | 43.80<br>(-3.85-123.62) | 63.05<br>(-6.56-169.44) | 1.20<br>(1.17-1.23) |
| Comoros | 43.97<br>(-3.33-135.14) | 195.33<br>(-15.89-555.05) | 344.25<br>(300.44-407.23) | 20.83<br>(-1.57-63.68) | 37.00<br>(-2.97-106.18) | 1.92<br>(1.91-1.93) |
| Congo | 297.20<br>(-24.33-847.02) | 1315.71<br>(-120.01-3632.51) | 342.71<br>(294.79-411.34) | 25.50<br>(-2.10-73.22) | 41.40<br>(-3.63-117.31) | 1.55<br>(1.51-1.59) |
| Cook Islands | 9.07<br>(-1.02-23.54) | 21.71<br>(-2.62-55.96) | 139.41<br>(124.31-169.36) | 66.95<br>(-7.44-175.27) | 85.01<br>(-10.32-217.46) | 0.74<br>(0.68-0.79) |
| Costa Rica | 898.23<br>(-91.24-2438.20) | 3684.06<br>(-400.15-9925.72) | 310.15<br>(280.43-368.33) | 50.40<br>(-5.08-137.17) | 66.54<br>(-7.21-179.25) | 0.90<br>(0.88-0.92) |
| Croatia | 2624.61<br>(-257.68-7262.21) | 4287.85<br>(-448.43-11668.27) | 63.37<br>(50.90-85.22) | 41.81<br>(-4.04-116.41) | 52.32<br>(-5.61-140.02) | 0.77<br>(0.74-0.81) |
| Cuba | 4340.16<br>(-355.98-12451.86) | 11857.98<br>(-1168.99-31969.75) | 173.22<br>(149.27-221.29) | 42.50<br>(-3.49-121.84) | 62.33<br>(-6.20-167.36) | 1.30<br>(1.27-1.33) |

| | | | | | | |
|---|---|---|---|---|---|---|
| Cyprus | 345.15 (-28.33-994.21) | 1206.38 (-120.98-3228.09) | 249.52 (216.51-306.23) | 41.27 (-3.39-118.88) | 59.55 (-6.00-158.63) | 1.21 (1.15-1.28) |
| Czechia | 6166.32 (-624.68-16537.97) | 10509.36 (-1162.29-27925.26) | 70.43 (60.63-85.13) | 45.53 (-4.59-122.42) | 52.78 (-5.86-138.17) | 0.48 (0.48-0.49) |
| Côte d'Ivoire | 1403.96 (-112.47-4093.61) | 5695.70 (-486.80-15685.65) | 305.69 (266.84-366.83) | 30.17 (-2.34-88.29) | 43.97 (-3.65-124.77) | 1.19 (1.17-1.21) |
| Democratic People's Republic of Korea | 3598.64 (-327.75-10376.05) | 11443.01 (-969.26-33123.88) | 217.98 (180.40-275.07) | 21.00 (-1.89-60.84) | 34.10 (-2.87-97.96) | 1.57 (1.45-1.68) |
| Democratic Republic of the Congo | 3224.38 (-217.75-9509.15) | 13675.70 (-1036.38-40873.89) | 324.13 (265.67-390.96) | 19.07 (-1.28-55.84) | 33.68 (-2.48-101.31) | 1.81 (1.75-1.87) |
| Denmark | 3551.56 (-313.91-10223.25) | 6158.58 (-587.10-16717.91) | 73.41 (62.07-92.69) | 46.68 (-4.19-132.67) | 57.36 (-5.63-155.08) | 0.66 (0.61-0.71) |
| Djibouti | 22.41 (-1.81-66.16) | 174.40 (-13.07-495.40) | 678.19 (576.17-789.97) | 14.02 (-1.10-42.21) | 22.69 (-1.68-66.74) | 1.58 (1.53-1.62) |
| Dominica | 33.44 (-3.27-90.04) | 62.40 (-7.17-162.58) | 86.61 (73.54-117.24) | 58.34 (-5.75-156.47) | 72.75 (-8.32-189.65) | 0.70 (0.66-0.73) |
| Dominican Republic | 1501.28 (-127.48-4154.82) | 5943.22 (-590.64-16273.07) | 295.88 (258.50-352.97) | 38.44 (-3.21-107.11) | 58.17 (-5.73-160.05) | 1.39 (1.34-1.43) |
| Ecuador | 2804.11 (-261.19-7576.53) | 12239.85 (-1272.66-32470.72) | 336.50 (291.30-414.93) | 50.72 (-4.67-138.79) | 73.47 (-7.62-195.72) | 1.28 (1.24-1.33) |
| Egypt | 13974.36 (-1419.18-37971.71) | 47765.56 (-6009.90-121024.94) | 241.81 (208.43-332.62) | 47.10 (-4.62-129.32) | 67.35 (-8.29-172.27) | 1.11 (1.07-1.14) |
| El Salvador | 1577.01 (-162.71-4342.47) | 4239.49 (-459.75-10952.02) | 168.83 (148.79-208.36) | 52.26 (-5.36-145.11) | 69.84 (-7.60-180.14) | 0.93 (0.88-0.99) |
| Equatorial Guinea | 54.96 (-4.16-160.43) | 289.56 (-26.75-789.83) | 426.90 (363.01-543.43) | 25.75 (-1.96-75.18) | 50.28 (-4.48-139.69) | 2.31 (2.24-2.37) |
| Eritrea | 141.38 (-11.49-412.64) | 599.43 (-43.89-1798.42) | 323.97 (253.39-416.04) | 10.79 (-0.84-31.51) | 18.76 (-1.35-56.47) | 1.78 (1.75-1.81) |
| Estonia | 941.92 (-84.66-2526.74) | 1356.72 (-142.89-3600.66) | 44.04 (34.25-60.70) | 46.05 (-4.16-123.42) | 55.89 (-6.02-147.38) | 0.72 (0.69-0.75) |
| Eswatini | 140.45 (-14.24-380.49) | 388.11 (-40.20-1008.45) | 176.32 (153.44-218.23) | 46.48 (-4.69-127.67) | 65.28 (-6.68-169.74) | 1.04 (0.95-1.14) |
| Ethiopia | 3241.69 (-238.30-9621.20) | 9987.09 (-774.84-29511.69) | 208.08 (178.00-245.08) | 14.94 (-1.11-45.08) | 21.15 (-1.61-63.32) | 1.13 (1.10-1.16) |
| Fiji | 223.06 (-21.77-596.75) | 629.68 (-74.45-1584.39) | 182.29 (155.48-236.40) | 54.46 (-5.26-148.90) | 74.50 (-8.65-189.48) | 0.96 (0.90-1.01) |
| Finland | 3566.32 (-332.00-9885.91) | 7066.30 (-724.97-19261.68) | 98.14 (84.77-123.35) | 51.25 (-4.82-141.66) | 63.59 (-6.57-172.60) | 0.68 (0.64-0.73) |
| France | 34060.87 (-3002.03-98797.02) | 73067.40 (-6486.90-204628.11) | 114.52 (96.80-143.16) | 43.19 (-3.86-125.30) | 59.21 (-5.26-165.04) | 1.02(0.92-1.11) |
| Gabon | 215.95 (-19.32-604.40) | 659.17 (-68.30-1786.17) | 205.24 (175.69-259.09) | 36.79 (-3.26-103.69) | 56.90 (-5.79-154.64) | 1.38 (1.35-1.41) |
| Gambia | 116.93 (-9.61-336.43) | 460.72 (-41.76-1280.20) | 294.02 (259.64-358.91) | 30.59 (-2.49-89.09) | 43.38 (-3.82-122.30) | 1.12 (1.09-1.15) |
| Georgia | 2314.33 (-212.96-6369.97) | 2353.55 (-233.46-6385.25) | 1.69 (-4.23-12.39) | 36.60 (-3.34-101.44) | 40.88 (-4.10-110.15) | 0.38 (0.38-0.39) |
| Germany | 67186.38 (-6235.22-183691.32) | 109573.52 (-10844.68-294559.90) | 63.09 (53.86-80.06) | 54.97 (-5.11-149.70) | 63.62 (-6.44-168.74) | 0.45 (0.43-0.47) |

| | | | | | | |
|---|---|---|---|---|---|---|
| Ghana | 1727.63<br>(-133.60-5091.80) | 8475.92<br>(-714.44-23455.97) | 390.61<br>(342.70-468.71) | 24.91<br>(-1.89-75.52) | 45.57<br>(-3.71-130.41) | 1.94<br>(1.92-1.96) |
| Greece | 7469.44<br>(-702.05-20511.07) | 13489.85<br>(-1356.99-36878.30) | 80.60<br>(67.08-104.25) | 49.86<br>(-4.70-136.70) | 65.08<br>(-6.58-177.53) | 0.93<br>(0.86-1.00) |
| Greenland | 14.46<br>(-1.43-39.24) | 35.41<br>(-3.57-94.08) | 144.95<br>(127.74-173.86) | 39.05<br>(-3.78-105.95) | 47.94<br>(-4.75-129.31) | 0.68<br>(0.65-0.71) |
| Grenada | 28.35<br>(-2.50-81.21) | 72.85<br>(-7.60-197.96) | 156.97<br>(134.24-201.28) | 43.27<br>(-3.90-122.69) | 60.40<br>(-6.19-165.16) | 1.09<br>(1.05-1.13) |
| Guam | 50.52<br>(-5.29-134.04) | 151.86<br>(-16.80-403.30) | 200.57<br>(179.75-234.57) | 57.94<br>(-5.96-156.71) | 72.22<br>(-7.98-191.31) | 0.71<br>(0.68-0.74) |
| Guatemala | 1680.72<br>(-148.07-4697.67) | 6912.30<br>(-693.55-18517.00) | 311.27<br>(278.78-372.87) | 45.02<br>(-3.92-126.72) | 60.69<br>(-6.05-162.31) | 0.98<br>(0.97-0.99) |
| Guinea | 840.87<br>(-66.52-2483.45) | 2072.23<br>(-166.92-5992.87) | 146.44<br>(120.22-175.79) | 24.48<br>(-1.92-72.81) | 34.20<br>(-2.73-99.54) | 1.04<br>(1.01-1.08) |
| Guinea-Bissau | 107.69<br>(-8.54-307.15) | 301.38<br>(-25.19-864.50) | 179.86<br>(154.38-215.89) | 25.23<br>(-2.03-73.51) | 36.40<br>(-2.97-105.53) | 1.15<br>(1.14-1.17) |
| Guyana | 169.08<br>(-14.70-474.46) | 393.33<br>(-38.34-1082.97) | 132.63<br>(112.36-167.00) | 42.33<br>(-3.66-120.27) | 57.37<br>(-5.49-159.18) | 1.00<br>(0.98-1.03) |
| Haiti | 670.34<br>(-49.89-2045.14) | 2574.99<br>(-205.87-7301.77) | 284.13<br>(242.94-338.77) | 19.11<br>(-1.42-58.16) | 31.49<br>(-2.49-91.11) | 1.74<br>(1.70-1.78) |
| Honduras | 940.91<br>(-84.10-2576.88) | 3940.95<br>(-430.07-10313.57) | 318.84<br>(287.87-369.48) | 44.28<br>(-3.94-122.24) | 58.89<br>(-6.42-155.85) | 0.93<br>(0.90-0.96) |
| Hungary | 6691.85<br>(-597.38-18415.16) | 9919.59<br>(-982.85-26877.07) | 48.23<br>(38.68-67.13) | 46.03<br>(-4.11-126.47) | 54.72<br>(-5.54-147.36) | 0.57<br>(0.56-0.59) |
| Iceland | 157.59<br>(-15.34-427.11) | 371.74<br>(-36.76-994.88) | 135.89<br>(121.34-162.63) | 57.55<br>(-5.65-155.35) | 68.50<br>(-6.85-182.32) | 0.56<br>(0.54-0.57) |
| India | 66788.91<br>(-4738.25-196448.68) | 351585.14<br>(-27811.55-1028334.05) | 426.41<br>(370.55-507.24) | 12.90<br>(-0.90-38.29) | 27.77<br>(-2.18-81.20) | 2.70<br>(2.63-2.78) |
| Indonesia | 14453.03<br>(-1078.20-42333.63) | 71651.20<br>(-5645.01-207717.66) | 395.75<br>(347.08-465.66) | 12.88<br>(-0.95-38.26) | 25.45<br>(-1.98-74.36) | 2.39<br>(2.30-2.47) |
| Iran<br>(Islamic Republic of) | 9277.80<br>(-800.77-26415.59) | 44994.39<br>(-4873.25-118696.41) | 384.97<br>(332.74-515.68) | 32.77<br>(-2.78-93.99) | 54.30<br>(-5.79-143.61) | 1.61<br>(1.58-1.63) |
| Iraq | 4026.63<br>(-390.44-10600.59) | 15479.54<br>(-1696.58-40324.74) | 284.43<br>(253.46-357.97) | 49.30<br>(-4.72-131.08) | 58.31<br>(-6.36-152.34) | 0.57<br>(0.55-0.59) |
| Ireland | 1989.02<br>(-178.37-5553.40) | 4873.71<br>(-495.85-12916.40) | 145.03<br>(127.14-176.14) | 50.23<br>(-4.61-139.22) | 64.47<br>(-6.64-169.89) | 0.81<br>(0.77-0.85) |
| Israel | 2467.49<br>(-237.83-6737.57) | 7376.17<br>(-704.26-20371.93) | 198.93<br>(180.13-232.19) | 51.92<br>(-5.06-141.36) | 63.08<br>(-6.06-173.03) | 0.58<br>(0.54-0.62) |
| Italy | 39993.62<br>(-3385.64-113457.83) | 74183.83<br>(-7012.06-204643.56) | 85.49<br>(75.43-102.82) | 46.14<br>(-3.94-129.82) | 57.51<br>(-5.50-157.29) | 0.66<br>(0.64-0.69) |
| Jamaica | 806.90<br>(-76.24-2310.07) | 2039.12<br>(-210.50-5382.15) | 152.71<br>(131.72-196.70) | 47.40<br>(-4.51-135.47) | 65.95<br>(-6.80-174.11) | 1.14<br>(1.09-1.20) |
| Japan | 69247.95<br>(-5508.99-205780.42) | 140243.81<br>(-10911.39-420874.53) | 102.52<br>(88.69-119.69) | 40.07<br>(-3.19-119.22) | 48.13<br>(-3.85-141.87) | 0.56<br>(0.53-0.58) |
| Jordan | 737.58<br>(-77.15-1914.16) | 5870.27<br>(-742.00-14855.33) | 695.89<br>(628.63-849.22) | 50.17<br>(-5.08-132.35) | 68.73<br>(-8.54-176.06) | 1.04<br>(1.01-1.06) |
| Kazakhstan | 4613.43<br>(-419.80-12614.50) | 8279.27<br>(-844.80-22480.99) | 79.46<br>(67.58-103.60) | 36.00<br>(-3.26-99.29) | 44.44<br>(-4.48-120.90) | 0.67<br>(0.66-0.69) |

| | | | | | | |
|---|---|---|---|---|---|---|
| Kenya | 1848.34<br>(-144.22-5289.00) | 9189.21<br>(-766.43-26159.07) | 397.16<br>(360.50-459.01) | 20.91<br>(-1.61-60.86) | 35.74<br>(-2.93-102.81) | 1.75<br>(1.70-1.80) |
| Kiribati | 21.29<br>(-2.22-57.53) | 56.77<br>(-6.95-145.11) | 166.60<br>(146.65-208.32) | 51.89<br>(-5.28-142.78) | 68.47<br>(-8.05-177.75) | 0.84<br>(0.76-0.93) |
| Kuwait | 386.13<br>(-42.73-989.86) | 2830.36<br>(-371.53-6847.76) | 633.01<br>(556.27-810.07) | 52.24<br>(-5.67-137.15) | 74.24<br>(-9.46-184.77) | 1.20<br>(1.18-1.22) |
| Kyrgyzstan | 1025.41<br>(-92.43-2812.92) | 2123.72<br>(-227.50-5639.90) | 107.11<br>(92.49-135.41) | 34.23<br>(-3.04-93.87) | 41.48<br>(-4.31-110.75) | 0.61<br>(0.59-0.63) |
| Lao People's Democratic Republic | 299.64<br>(-21.14-902.22) | 1286.48<br>(-89.08-3745.31) | 329.35<br>(263.48-399.69) | 13.38<br>(-0.94-40.79) | 24.23<br>(-1.67-71.27) | 2.16<br>(2.07-2.26) |
| Latvia | 1694.32<br>(-176.99-4549.63) | 2044.63<br>(-226.70-5545.45) | 20.68<br>(13.81-34.19) | 47.38<br>(-4.94-126.80) | 56.27<br>(-6.27-152.31) | 0.59<br>(0.57-0.61) |
| Lebanon | 1003.44<br>(-99.03-2727.85) | 3585.39<br>(-413.19-9199.43) | 257.31<br>(230.78-327.76) | 43.92<br>(-4.30-120.46) | 61.49<br>(-7.18-157.29) | 1.07<br>(0.99-1.14) |
| Lesotho | 319.31<br>(-29.23-857.18) | 571.07<br>(-56.50-1523.43) | 78.84<br>(64.51-104.41) | 36.72<br>(-3.32-99.58) | 50.60<br>(-4.95-136.85) | 1.03<br>(1.01-1.05) |
| Liberia | 441.58<br>(-37.04-1249.78) | 1291.51<br>(-126.82-3539.51) | 192.48<br>(165.26-237.53) | 36.97<br>(-3.09-103.82) | 52.04<br>(-4.97-144.81) | 1.19<br>(1.14-1.23) |
| Libya | 885.89<br>(-85.07-2422.97) | 3897.32<br>(-487.31-9846.32) | 339.93<br>(289.47-462.04) | 44.49<br>(-4.18-123.61) | 65.47<br>(-8.01-166.44) | 1.27<br>(1.24-1.30) |
| Lithuania | 1996.11<br>(-188.78-5503.65) | 2924.34<br>(-299.73-7660.30) | 46.50<br>(36.44-64.55) | 44.16<br>(-4.14-121.81) | 54.96<br>(-5.79-142.71) | 0.72<br>(0.69-0.76) |
| Luxembourg | 269.80<br>(-22.63-758.30) | 638.55<br>(-59.49-1734.40) | 136.68<br>(121.23-165.62) | 50.36<br>(-4.27-141.41) | 62.63<br>(-5.85-169.86) | 0.69<br>(0.66-0.72) |
| Madagascar | 787.55<br>(-52.91-2468.68) | 3127.41<br>(-230.36-8970.78) | 297.11<br>(242.29-371.16) | 14.89<br>(-0.99-46.37) | 24.76<br>(-1.78-70.40) | 1.66<br>(1.60-1.72) |
| Malawi | 742.51<br>(-61.28-2195.06) | 2451.18<br>(-195.22-7036.38) | 230.12<br>(190.13-282.09) | 18.09<br>(-1.49-53.90) | 29.87<br>(-2.34-87.70) | 1.66<br>(1.63-1.69) |
| Malaysia | 2799.27<br>(-231.42-7931.38) | 12581.44<br>(-1160.81-34309.88) | 349.45<br>(309.38-421.94) | 27.15<br>(-2.20-78.47) | 41.60<br>(-3.79-113.86) | 1.42<br>(1.37-1.48) |
| Maldives | 21.79<br>(-1.73-63.65) | 156.89<br>(-14.81-421.95) | 619.88<br>(535.93-775.40) | 19.65<br>(-1.55-58.20) | 36.93<br>(-3.36-102.45) | 2.25<br>(2.16-2.34) |
| Mali | 941.78<br>(-74.07-2748.73) | 2880.60<br>(-222.12-8331.50) | 205.87<br>(171.89-251.45) | 21.82<br>(-1.66-64.42) | 29.32<br>(-2.21-85.50) | 0.96<br>(0.90-1.01) |
| Malta | 192.01<br>(-16.40-551.83) | 543.51<br>(-52.03-1478.28) | 183.06<br>(160.40-228.99) | 44.75<br>(-3.84-128.65) | 62.03<br>(-6.06-169.02) | 1.03<br>(0.92-1.13) |
| Marshall Islands | 9.21<br>(-1.01-24.56) | 27.35<br>(-3.33-69.65) | 196.95<br>(176.16-241.26) | 52.86<br>(-5.69-143.21) | 66.41<br>(-7.90-173.31) | 0.68<br>(0.65-0.71) |
| Mauritania | 413.67<br>(-35.36-1159.30) | 1279.02<br>(-127.19-3448.80) | 209.19<br>(185.28-253.65) | 40.29<br>(-3.40-112.76) | 55.95<br>(-5.46-151.97) | 1.03<br>(1.00-1.06) |
| Mauritius | 225.45<br>(-17.74-639.95) | 838.78<br>(-78.05-2236.74) | 272.05<br>(233.46-344.82) | 29.04<br>(-2.27-82.50) | 44.52<br>(-4.11-119.00) | 1.42<br>(1.37-1.48) |
| Mexico | 24139.82<br>(-2393.80-64553.11) | 92604.38<br>(-10331.83-242413.59) | 283.62<br>(260.37-333.81) | 53.89<br>(-5.26-144.93) | 70.41<br>(-7.79-185.21) | 0.83<br>(0.81-0.85) |
| Micronesia (Federated States of) | 28.57<br>(-3.07-74.39) | 61.76<br>(-7.28-153.57) | 116.16<br>(98.59-154.99) | 56.47<br>(-6.01-148.12) | 72.49<br>(-8.25-184.70) | 0.75<br>(0.67-0.83) |
| Monaco | 36.72<br>(-3.28-100.75) | 59.30<br>(-5.81-158.69) | 61.50<br>(51.21-80.33) | 58.65<br>(-5.28-159.47) | 68.97<br>(-6.96-181.55) | 0.51<br>(0.49-0.53) |

| | | | | | | |
|---|---|---|---|---|---|---|
| Mongolia | 319.43<br>(-29.24-898.16) | 910.89<br>(-93.15-2441.78) | 185.16<br>(166.18-217.14) | 30.02<br>(-2.70-85.09) | 35.99<br>(-3.54-97.65) | 0.49<br>(0.42-0.56) |
| Montenegro | 292.20<br>(-28.33-784.19) | 531.67<br>(-56.64-1391.66) | 81.95<br>(69.95-109.50) | 45.91<br>(-4.45-123.54) | 54.77<br>(-5.85-144.06) | 0.61<br>(0.60-0.63) |
| Morocco | 5039.69<br>(-512.38-13769.44) | 18399.78<br>(-1958.28-47936.73) | 265.10<br>(239.12-306.49) | 34.30<br>(-3.44-94.14) | 50.55<br>(-5.32-132.63) | 1.28<br>(1.28-1.29) |
| Mozambique | 1182.11<br>(-84.34-3494.09) | 3763.49<br>(-278.25-10778.75) | 218.37<br>(177.50-268.24) | 17.85<br>(-1.26-54.04) | 29.62<br>(-2.14-85.52) | 1.69<br>(1.64-1.75) |
| Myanmar | 3891.67<br>(-287.30-11575.36) | 13098.03<br>(-1048.53-37651.60) | 236.57<br>(199.92-288.38) | 15.33<br>(-1.12-46.08) | 24.23<br>(-1.89-69.44) | 1.56<br>(1.53-1.59) |
| Namibia | 203.18<br>(-15.27-621.69) | 718.05<br>(-58.50-2003.04) | 253.40<br>(211.82-325.50) | 29.98<br>(-2.25-91.76) | 48.73<br>(-3.90-137.61) | 1.58<br>(1.52-1.63) |
| Nauru | 3.26<br>(-0.38-8.28) | 5.13<br>(-0.69-12.63) | 57.51<br>(46.55-81.40) | 60.52<br>(-6.86-158.09) | 78.18<br>(-10.12-196.07) | 0.76<br>(0.74-0.79) |
| Nepal | 1417.14<br>(-98.56-4300.57) | 6716.84<br>(-519.61-19291.67) | 373.97<br>(305.71-477.61) | 12.91<br>(-0.89-39.29) | 26.48<br>(-2.02-76.00) | 2.54<br>(2.46-2.62) |
| Netherlands | 9817.22<br>(-941.33-26899.55) | 20176.66<br>(-2035.50-55122.77) | 105.52<br>(91.77-125.84) | 50.71<br>(-4.89-138.00) | 62.38<br>(-6.38-169.82) | 0.67<br>(0.66-0.69) |
| New Zealand | 2162.90<br>(-207.01-5892.24) | 5873.33<br>(-643.53-15756.39) | 171.55<br>(151.77-206.37) | 56.43<br>(-5.42-152.81) | 73.84<br>(-8.13-198.75) | 0.90<br>(0.87-0.94) |
| Nicaragua | 815.19<br>(-80.46-2206.15) | 3406.83<br>(-391.14-9003.38) | 317.92<br>(284.70-383.16) | 50.88<br>(-4.95-138.18) | 66.65<br>(-7.64-175.91) | 0.88<br>(0.84-0.91) |
| Niger | 669.05<br>(-51.27-1930.05) | 2550.96<br>(-190.66-7493.79) | 281.28<br>(237.03-339.62) | 21.54<br>(-1.62-63.80) | 27.69<br>(-2.04-81.14) | 0.80<br>(0.79-0.81) |
| Nigeria | 12836.06<br>(-1032.21-37755.23) | 45180.88<br>(-4030.18-125193.03) | 251.98<br>(222.51-300.65) | 27.83<br>(-2.21-81.46) | 44.76<br>(-3.90-125.39) | 1.52<br>(1.42-1.61) |
| Niue | 1.25<br>(-0.13-3.30) | 1.73<br>(-0.19-4.42) | 38.44<br>(27.06-62.01) | 59.07<br>(-6.06-156.26) | 77.94<br>(-8.65-200.36) | 0.88<br>(0.82-0.94) |
| North Macedonia | 803.25<br>(-79.30-2138.56) | 1684.74<br>(-173.28-4593.61) | 109.74<br>(96.08-137.34) | 41.54<br>(-4.05-111.25) | 50.06<br>(-5.12-136.36) | 0.64<br>(0.63-0.65) |
| Northern Mariana Islands | 14.62<br>(-1.62-38.17) | 48.41<br>(-5.89-120.81) | 231.07<br>(203.17-288.74) | 62.79<br>(-6.89-168.27) | 79.13<br>(-9.30-200.44) | 0.68<br>(0.60-0.77) |
| Norway | 2845.12<br>(-241.23-8088.05) | 5117.09<br>(-471.49-14106.03) | 79.86<br>(71.78-95.79) | 45.65<br>(-3.92-127.42) | 55.68<br>(-5.16-152.58) | 0.62<br>(0.61-0.64) |
| Oman | 271.16<br>(-24.91-754.87) | 1703.50<br>(-200.10-4273.71) | 528.23<br>(436.04-747.66) | 34.56<br>(-3.08-97.50) | 64.59<br>(-7.44-164.98) | 2.17<br>(2.12-2.21) |
| Pakistan | 10838.33<br>(-884.91-31199.65) | 45963.13<br>(-3977.31-131671.44) | 324.08<br>(279.15-382.65) | 18.08<br>(-1.46-52.29) | 33.22<br>(-2.80-97.90) | 2.29<br>(2.16-2.42) |
| Palau | 6.41<br>(-0.67-16.87) | 20.27<br>(-2.46-51.33) | 216.24<br>(194.01-266.41) | 62.08<br>(-6.43-164.77) | 77.45<br>(-9.15-197.81) | 0.65<br>(0.59-0.71) |
| Palestine | 421.98<br>(-44.56-1128.89) | 1730.44<br>(-214.73-4315.89) | 310.07<br>(275.25-378.32) | 47.38<br>(-4.92-127.20) | 60.89<br>(-7.39-153.56) | 0.78<br>(0.76-0.80) |
| Panama | 714.39<br>(-63.77-1974.79) | 2968.89<br>(-314.70-7934.28) | 315.58<br>(278.47-385.84) | 47.79<br>(-4.28-132.27) | 67.16<br>(-7.13-179.64) | 1.09<br>(1.06-1.11) |
| Papua New Guinea | 653.67<br>(-56.35-1851.87) | 2531.08<br>(-245.05-6938.27) | 287.21<br>(257.65-331.98) | 30.75<br>(-2.61-87.80) | 40.11<br>(-3.73-111.39) | 0.81<br>(0.76-0.87) |
| Paraguay | 1101.74<br>(-107.26-3044.14) | 3795.98<br>(-421.24-10158.97) | 244.54<br>(221.25-294.14) | 48.45<br>(-4.68-134.06) | 63.13<br>(-6.95-169.30) | 0.89<br>(0.87-0.90) |

| | | | | | | |
|---|---|---|---|---|---|---|
| Peru | 5697.47<br>(-541.75-15578.31) | 21692.96<br>(-2363.27-58152.69) | 280.75<br>(254.73-324.35) | 45.76<br>(-4.31-125.96) | 63.39<br>(-6.87-170.68) | 1.08<br>(1.05-1.11) |
| Philippines | 5555.88<br>(-405.72-16620.35) | 24951.04<br>(-1881.59-72911.26) | 349.09<br>(313.32-393.15) | 16.78<br>(-1.21-51.03) | 27.60<br>(-2.05-80.96) | 1.60<br>(1.51-1.70) |
| Poland | 17928.88<br>(-1668.40-48391.68) | 34301.95<br>(-3582.63-92178.82) | 91.32<br>(82.62-109.79) | 41.03<br>(-3.83-110.90) | 50.33<br>(-5.29-135.38) | 0.67<br>(0.65-0.69) |
| Portugal | 6494.46<br>(-596.40-18164.12) | 13691.06<br>(-1407.54-36078.99) | 110.81<br>(95.90-136.33) | 47.07<br>(-4.34-131.62) | 62.63<br>(-6.52-163.43) | 0.83<br>(0.76-0.90) |
| Puerto Rico | 2292.96<br>(-226.41-6108.45) | 5016.58<br>(-546.27-13383.98) | 118.78<br>(102.79-145.39) | 63.69<br>(-6.31-169.65) | 80.42<br>(-8.98-211.60) | 0.79<br>(0.74-0.84) |
| Qatar | 89.18<br>(-10.09-232.69) | 1203.76<br>(-154.75-2986.09) | 1249.86<br>(1112.50-1605.70) | 52.04<br>(-5.60-140.26) | 72.47<br>(-8.93-186.97) | 1.03<br>(1.01-1.06) |
| Republic of Korea | 12437.25<br>(-1119.89-35321.23) | 55348.93<br>(-4640.22-164506.70) | 345.03<br>(283.70-426.03) | 39.32<br>(-3.52-111.86) | 58.71<br>(-4.93-174.25) | 1.51<br>(1.36-1.65) |
| Republic of Moldova | 1945.57<br>(-185.45-5300.20) | 3346.23<br>(-358.54-8865.00) | 71.99<br>(58.60-98.51) | 43.47<br>(-4.09-119.38) | 56.97<br>(-6.12-150.11) | 0.94<br>(0.92-0.97) |
| Romania | 11260.33<br>(-955.76-30938.42) | 17568.05<br>(-1706.78-47655.83) | 56.02<br>(45.26-77.98) | 39.71<br>(-3.36-109.58) | 50.22<br>(-4.92-136.17) | 0.78<br>(0.76-0.81) |
| Russian Federation | 76961.79<br>(-7251.91-211642.71) | 130796.75<br>(-13323.59-344632.60) | 69.95<br>(59.22-94.91) | 42.33<br>(-3.95-116.94) | 55.47<br>(-5.68-146.27) | 0.94<br>(0.92-0.97) |
| Rwanda | 512.22<br>(-39.33-1483.40) | 1877.73<br>(-157.86-5585.08) | 266.58<br>(222.10-316.57) | 16.98<br>(-1.29-50.03) | 26.71<br>(-2.20-81.65) | 1.47<br>(1.42-1.53) |
| Saint Kitts and Nevis | 17.16<br>(-1.53-49.05) | 52.04<br>(-5.67-135.21) | 203.21<br>(173.70-260.77) | 49.46<br>(-4.48-138.73) | 68.31<br>(-7.26-179.06) | 1.06<br>(1.03-1.09) |
| Saint Lucia | 38.01<br>(-3.46-105.89) | 151.18<br>(-15.01-419.76) | 297.72<br>(264.10-359.11) | 43.80<br>(-4.03-121.70) | 61.37<br>(-6.04-170.54) | 1.09<br>(1.06-1.12) |
| Saint Vincent and the Grenadines | 26.45<br>(-2.15-77.73) | 80.37<br>(-7.50-222.86) | 203.81<br>(176.38-248.15) | 37.95<br>(-3.08-111.04) | 54.96<br>(-5.09-152.96) | 1.26<br>(1.23-1.28) |
| Samoa | 58.83<br>(-7.01-146.56) | 119.68<br>(-15.39-300.90) | 103.44<br>(91.53-130.78) | 64.44<br>(-7.55-162.54) | 76.89<br>(-9.70-194.25) | 0.50<br>(0.45-0.56) |
| San Marino | 18.27<br>(-1.64-49.83) | 43.64<br>(-4.43-117.23) | 138.82<br>(125.29-167.39) | 54.48<br>(-4.93-148.01) | 66.08<br>(-6.86-175.43) | 0.61<br>(0.58-0.65) |
| Sao Tome and Principe | 22.96<br>(-2.02-65.56) | 62.54<br>(-6.12-170.31) | 172.40<br>(151.12-211.78) | 34.79<br>(-3.03-99.69) | 50.28<br>(-4.81-139.87) | 1.20<br>(1.18-1.22) |
| Saudi Arabia | 2885.24<br>(-293.92-7701.86) | 17739.35<br>(-2154.16-43804.17) | 514.83<br>(436.44-704.53) | 44.23<br>(-4.49-120.77) | 69.65<br>(-8.25-175.77) | 1.46<br>(1.40-1.52) |
| Senegal | 1016.22<br>(-83.44-2916.14) | 3402.04<br>(-289.61-9534.57) | 234.77<br>(207.33-275.15) | 29.66<br>(-2.40-85.75) | 40.50<br>(-3.39-114.50) | 0.96<br>(0.93-0.99) |
| Serbia | 4795.66<br>(-467.58-12888.04) | 8235.64<br>(-877.79-22029.75) | 71.73<br>(57.78-97.88) | 40.65<br>(-3.92-110.37) | 52.89<br>(-5.67-141.55) | 0.90<br>(0.88-0.91) |
| Seychelles | 20.06<br>(-1.88-55.81) | 66.20<br>(-7.70-174.93) | 230.01<br>(195.66-302.31) | 36.43<br>(-3.45-100.76) | 51.75<br>(-5.87-137.53) | 1.12<br>(1.07-1.17) |
| Sierra Leone | 512.88<br>(-38.55-1540.81) | 1450.69<br>(-116.67-4151.87) | 182.85<br>(154.22-217.02) | 24.52<br>(-1.83-73.88) | 35.29<br>(-2.79-103.30) | 1.14<br>(1.09-1.20) |
| Singapore | 905.81<br>(-71.00-2631.36) | 5876.21<br>(-470.88-16837.44) | 548.72<br>(457.29-655.11) | 37.74<br>(-2.93-109.44) | 66.50<br>(-5.32-190.31) | 1.78<br>(1.69-1.87) |
| Slovakia | 2730.36<br>(-274.89-7384.77) | 4952.39<br>(-499.21-13168.75) | 81.38<br>(71.44-102.11) | 46.03<br>(-4.62-124.56) | 53.52<br>(-5.43-142.65) | 0.46<br>(0.44-0.48) |

| | | | | | | |
|---|---|---|---|---|---|---|
| Slovenia | 1079.19 (-104.30-2935.84) | 2140.66 (-214.78-5687.59) | 98.36 (85.05-125.60) | 43.85 (-4.23-119.22) | 52.71 (-5.39-140.53) | 0.62 (0.61-0.64) |
| Solomon Islands | 61.99 (-5.22-175.17) | 215.98 (-22.38-585.94) | 248.40 (220.97-293.37) | 38.94 (-3.25-111.10) | 52.86 (-5.34-145.63) | 0.94 (0.87-1.00) |
| Somalia | 522.88 (-38.50-1451.84) | 1861.66 (-143.48-5165.20) | 256.04 (216.18-306.86) | 17.80 (-1.29-51.75) | 25.26 (-1.94-70.83) | 1.16 (1.12-1.19) |
| South Africa | 10066.71 (-923.95-27484.45) | 30343.35 (-3174.26-79888.13) | 201.42 (182.52-243.93) | 47.36 (-4.31-130.59) | 62.36 (-6.47-165.52) | 0.90 (0.89-0.91) |
| South Sudan | 354.52 (-27.80-1069.01) | 856.48 (-65.04-2507.67) | 141.59 (107.16-183.32) | 13.34 (-1.04-40.31) | 18.78 (-1.40-56.15) | 1.12 (1.07-1.17) |
| Spain | 29108.32 (-2901.55-80742.64) | 60552.08 (-6314.33-163332.96) | 108.02 (93.23-132.67) | 54.54 (-5.45-150.74) | 68.45 (-7.21-184.80) | 0.64 (0.57-0.71) |
| Sri Lanka | 2277.36 (-161.50-6696.64) | 8698.18 (-651.54-25624.59) | 281.94 (239.83-342.98) | 19.21 (-1.35-56.59) | 31.42 (-2.34-92.02) | 1.61 (1.56-1.66) |
| Sudan | 3118.83 (-261.89-8799.56) | 11290.05 (-1192.96-29891.89) | 262.00 (223.11-337.28) | 32.17 (-2.67-91.48) | 52.16 (-5.43-140.93) | 1.60 (1.56-1.64) |
| Suriname | 103.85 (-9.19-293.69) | 367.00 (-34.50-999.62) | 253.39 (224.68-303.19) | 38.69 (-3.36-110.06) | 55.59 (-5.17-152.23) | 1.20 (1.18-1.21) |
| Sweden | 5338.42 (-490.40-15425.50) | 9375.08 (-917.41-26347.43) | 75.62 (64.76-93.46) | 38.54 (-3.65-111.04) | 49.64 (-4.97-136.35) | 0.76 (0.70-0.83) |
| Switzerland | 4568.32 (-384.43-12923.63) | 8870.31 (-759.91-24454.31) | 94.17 (83.87-112.17) | 46.25 (-3.91-130.37) | 53.93 (-4.70-146.77) | 0.52 (0.49-0.54) |
| Syrian Arab Republic | 2408.11 (-239.57-6466.60) | 9377.63 (-1179.31-24199.07) | 289.42 (246.72-393.25) | 43.54 (-4.30-118.04) | 64.06 (-7.84-166.35) | 1.25 (1.21-1.28) |
| Taiwan (Province of China) | 6017.06 (-515.35-17446.13) | 25633.00 (-2512.24-73337.89) | 326.01 (282.09-393.17) | 35.72 (-3.03-104.29) | 62.39 (-6.13-177.95) | 1.89 (1.85-1.93) |
| Tajikistan | 855.99 (-78.65-2398.17) | 2293.66 (-245.97-6111.68) | 167.95 (148.17-204.48) | 31.01 (-2.79-87.30) | 36.04 (-3.72-97.33) | 0.48 (0.46-0.49) |
| Thailand | 8183.62 (-655.53-23833.08) | 45842.93 (-3995.38-130751.28) | 460.18 (396.66-567.62) | 20.34 (-1.61-59.37) | 41.57 (-3.64-118.83) | 2.44 (2.36-2.52) |
| Timor-Leste | 27.22 (-2.33-83.03) | 122.83 (-8.07-375.94) | 351.21 (251.06-464.21) | 7.80 (-0.68-23.49) | 13.66 (-0.90-42.16) | 1.94 (1.86-2.02) |
| Togo | 356.05 (-27.10-1013.54) | 1741.64 (-148.13-4916.40) | 389.16 (340.29-462.00) | 26.36 (-1.99-75.83) | 40.25 (-3.36-115.80) | 1.34 (1.32-1.36) |
| Tokelau | 0.71 (-0.07-1.88) | 1.08 (-0.12-2.73) | 51.31 (37.72-76.84) | 53.91 (-5.37-141.80) | 73.19 (-8.46-185.55) | 0.97 (0.91-1.03) |
| Tonga | 39.08 (-4.24-101.12) | 68.16 (-8.68-171.84) | 74.44 (62.47-100.54) | 65.41 (-6.98-170.61) | 81.98 (-10.33-208.12) | 0.64 (0.55-0.74) |
| Trinidad and Tobago | 440.50 (-39.27-1203.77) | 1322.86 (-147.25-3505.19) | 200.31 (175.06-246.23) | 51.49 (-4.58-141.32) | 67.75 (-7.51-179.68) | 0.94 (0.92-0.96) |
| Tunisia | 1890.37 (-180.83-5379.22) | 7661.54 (-870.99-20154.94) | 305.29 (265.99-385.29) | 35.81 (-3.37-102.52) | 55.37 (-6.23-146.02) | 1.44 (1.43-1.45) |
| Turkmenistan | 659.87 (-64.69-1777.85) | 1735.18 (-188.26-4615.29) | 272.74 (238.94-342.74) | 33.99 (-3.29-93.44) | 40.74 (-4.28-109.40) | 0.60 (0.59-0.62) |
| Tuvalu | 3.74 (-0.37-10.06) | 7.69 (-0.90-19.50) | 162.96 (147.60-193.64) | 51.39 (-5.01-139.06) | 69.88 (-8.08-177.63) | 0.97 (0.93-1.02) |
| Türkiye | 17314.65 (-1803.14-46614.56) | 64538.49 (-7666.24-166167.72) | 105.66 (87.51-148.18) | 47.35 (-4.84-128.41) | 66.37 (-7.81-170.88) | 1.08 (1.06-1.09) |

| | | | | | | |
|---|---|---|---|---|---|---|
| Uganda | 1257.62<br>(-91.53-3838.50) | 4932.15<br>(-392.63-14125.16) | 292.18<br>(250.26-343.53) | 18.11<br>(-1.32-55.56) | 29.49<br>(-2.32-86.13) | 1.64<br>(1.61-1.68) |
| Ukraine | 31180.49<br>(-2841.72-86080.00) | 40164.82<br>(-4198.73-106209.06) | 28.81<br>(19.00-49.87) | 43.49<br>(-3.90-120.17) | 53.06<br>(-5.59-139.96) | 0.70<br>(0.68-0.72) |
| United Arab Emirates | 280.90<br>(-26.74-750.80) | 5646.91<br>(-703.29-13808.40) | 1910.27<br>(1631.65-2609.34) | 42.55<br>(-3.87-118.44) | 67.40<br>(-7.98-172.85) | 1.54<br>(1.51-1.58) |
| United Kingdom | 49970.25<br>(-4784.47-134657.20) | 85642.93<br>(-9075.72-227898.56) | 71.39<br>(63.69-86.33) | 58.71<br>(-5.70-157.33) | 72.52<br>(-7.78-191.99) | 0.73<br>(0.64-0.81) |
| United Republic of Tanzania | 2802.12<br>(-215.41-8539.46) | 11035.99<br>(-904.55-30481.53) | 293.84<br>(257.31-352.22) | 24.31<br>(-1.85-73.48) | 39.54<br>(-3.17-109.84) | 1.62<br>(1.59-1.65) |
| United States of America | 212545.53<br>(-20945.25-569481.52) | 471042.37<br>(-52422.32-1227457.48) | 121.62<br>(105.93-156.48) | 70.46<br>(-7.00-187.75) | 86.42<br>(-9.71-226.10) | 0.47<br>(0.32-0.63) |
| United States Virgin Islands | 58.87<br>(-6.53-158.75) | 132.02<br>(-13.69-354.31) | 124.25<br>(108.77-154.48) | 64.63<br>(-7.05-175.28) | 76.88<br>(-8.10-201.79) | 0.56<br>(0.53-0.59) |
| Uruguay | 2115.37<br>(-201.11-5758.92) | 3674.16<br>(-361.92-9833.51) | 73.69<br>(62.25-97.47) | 55.42<br>(-5.29-150.96) | 72.47<br>(-7.23-194.18) | 0.87<br>(0.81-0.92) |
| Uzbekistan | 3918.45<br>(-386.39-10850.32) | 11763.93<br>(-1315.39-31271.47) | 200.22<br>(179.88-234.59) | 33.99<br>(-3.30-94.60) | 41.95<br>(-4.53-112.07) | 0.70<br>(0.69-0.72) |
| Vanuatu | 27.75<br>(-2.48-75.37) | 107.02<br>(-10.94-281.03) | 285.59<br>(256.54-335.60) | 39.09<br>(-3.42-107.76) | 53.64<br>(-5.36-143.10) | 1.01<br>(0.98-1.05) |
| Venezuela (Bolivarian Republic of) | 5425.54<br>(-536.71-15006.16) | 21452.69<br>(-2289.82-56063.77) | 295.40<br>(264.63-359.88) | 53.79<br>(-5.27-149.63) | 68.93<br>(-7.30-180.75) | 0.82<br>(0.81-0.83) |
| Viet Nam | 3314.29<br>(-260.62-10259.13) | 19046.42<br>(-1336.42-57845.54) | 474.68<br>(381.05-578.56) | 8.15<br>(-0.64-25.23) | 17.40<br>(-1.21-53.04) | 2.82<br>(2.66-2.98) |
| Yemen | 1185.05<br>(-86.16-3462.16) | 5929.07<br>(-498.51-16797.74) | 400.32<br>(350.10-481.46) | 22.38<br>(-1.60-66.24) | 38.06<br>(-3.10-108.51) | 1.80<br>(1.74-1.87) |
| Zambia | 649.28<br>(-48.86-1891.26) | 2943.04<br>(-260.27-8120.46) | 353.28<br>(301.77-417.41) | 20.79<br>(-1.57-61.10) | 37.25<br>(-3.19-106.22) | 1.92<br>(1.90-1.93) |
| Zimbabwe | 1175.33<br>(-88.97-3526.78) | 3284.37<br>(-285.25-8733.57) | 179.44<br>(153.85-228.82) | 26.85<br>(-2.01-80.93) | 43.19<br>(-3.67-117.94) | 1.49<br>(1.43-1.55) |
